# Genomic dimensions deconstruct the clinical heterogeneity of bipolar disorder

**DOI:** 10.1101/2025.06.23.25330155

**Authors:** Tracey van der Veen, Markos Tesfaye, Jessica Mei Kay Yang, Toni Boltz, Friederike S. David, Shane Crinion, Maria Koromina, Till F. M. Andlauer, Tim B. Bigdeli, Brandon J. Coombes, Tiffany A. Greenwood, Georgia Panagiotaropoulou, Nadine Parker, Heejong Sung, Nicholas Bass, Jonathan R. I. Coleman, José Guzman-Parra, Janos L. Kalman, Caroline C. McGrouther, Brittany L. Mitchell, Aaditya V. Rangan, Katie Scott, Alexey Shadrin, Daniel J. Smith, Annabel Vreeker, Kristina Adorjan, Diego Albani, Silvia Alemany, Ney Alliey-Rodriguez, Anastasia Antoniou, Michael Bauer, Eva C. Beins, Marco P. Boks, Rosa Bosch, Ben M. Brumpton, Nathalie Brunkhorst-Kanaan, Monika Budde, William Byerley, Judit Cabana-Domínguez, Murray J. Cairns, Bernardo Carpiniello, Miquel Casas, Pablo Cervantes, Chris Chatzinakos, Toni-Kim Clarke, Isabelle Claus, Cristiana Cruceanu, Alfredo Cuellar-Barboza, Piotr M. Czerski, Konstantinos Dafnas, Anders M. Dale, Nina Dalkner, J. Raymond DePaulo, Franziska Degenhardt, Srdjan Djurovic, Valentina Escott-Price, Ayman H. Fanous, Frederike T. Fellendorf, I. Nicol Ferrier, Liz Forty, Josef Frank, Oleksandr Frei, Nelson B. Freimer, Julie Garnham, Ian R. Gizer, Scott D. Gordon, Katherine Gordon-Smith, Tim Hahn, Marian L. Hamshere, Arvid Harder, Martin Hautzinger, Urs Heilbronner, Dennis Hellgren, Stefan Herms, Ian B. Hickie, Per Hoffmann, Peter A. Holmans, Stéphane Jamain, Lina Jonsson, James L. Kennedy, Sarah Kittel-Schneider, James A. Knowles, Elise Koch, Manolis Kogevinas, Thorsten M. Kranz, Steven A. Kushner, Catharina Lavebratt, Jacob Lawrence, Markus Leber, Penelope A. Lind, Susanne Lucae, Martin Lundberg, Donald J. MacIntyre, Wolfgang Maier, Adam X. Maihofer, Dolores Malaspina, Mirko Manchia, Eirini Maratou, Lina Martinsson, Melvin G. McInnis, James D. McKay, Helena Medeiros, Andreas Meyer-Lindenberg, Vincent Millischer, Derek W. Morris, Paraskevi Moutsatsou, Thomas W. Mühleisen, Claire O’Donovan, Catherine M. Olsen, Sergi Papiol, Antonio F. Pardiñas, Amy Perry, Andrea Pfennig, Claudia Pisanu, James B. Potash, Digby Quested, Mark H. Rapaport, Eline J. Regeer, John P. Rice, Margarita Rivera, Eva C. Schulte, Fanny Senner, Paul D. Shilling, Lisa Sindermann, Lea Sirignano, Dan Siskind, Claire Slaney, Olav B. Smeland, Janet L. Sobell, Maria Soler Artigas, Dan J. Stein, Frederike Stein, Beata Swiatkowska, Jackson G. Thorp, Claudio Toma, Leonardo Tondo, Paul A. Tooney, Marquis P. Vawter, Helmut Vedder, James T. R. Walters, Stephanie H. Witt, Allan H. Young, Peter P. Zandi, Lea Zillich, Estonian Biobank research team, Genomic Psychiatry Cohort (GPC) Investigators, HUNT All-In Psychiatry, Rolf Adolfsson, Lars Alfredsson, Lena Backlund, Bernhard T. Baune, Frank Bellivier, Susanne Bengesser, Wade H. Berrettini, Joanna M. Biernacka, Douglas Blackwood, Michael Boehnke, Gerome Breen, Vaughan J. Carr, Stanley Catts, Sven Cichon, Aiden Corvin, Nicholas Craddock, Udo Dannlowski, Dimitris Dikeos, Tõnu Esko, Bruno Etain, Panagiotis Ferentinos, Mark Frye, Janice M. Fullerton, Micha Gawlik, Elliot S. Gershon, Fernando S. Goes, Melissa J. Green, Joanna Hauser, Frans A. Henskens, Jens Hjerling-Leffler, Ian Jones, Lisa A. Jones, René S. Kahn, John R. Kelsoe, Tilo Kircher, George Kirov, Nene Kobayashi, Mikael Landén, Marion Leboyer, Melanie Lenger, Qingqin S. Li, Jolanta Lissowska, Carmel Loughland, Jurjen J. Luykx, Nicholas G. Martin, Carol A. Mathews, Fermin Mayoral, Susan L. McElroy, Andrew M. McIntosh, Sarah E. Medland, Ingrid Melle, Philip B. Mitchell, Gunnar Morken, Richard M. Myers, Chiara Möser, Bertram Müller-Myhsok, Benjamin M. Neale, Caroline M. Nievergelt, John I. Nurnberger, Markus M. Nöthen, Michael C. O’Donovan, Ketil J. Oedegaard, Tomas Olsson, Michael J. Owen, Sara A. Paciga, Christos Pantelis, Carlos N. Pato, Michele T. Pato, George P. Patrinos, Joanna M. Pawlak, Roy Perlis, Josep Antoni Ramos-Quiroga, Andreas Reif, Eva Z. Reininghaus, Marta Ribasés, Marcella Rietschel, Stephan Ripke, Guy A. Rouleau, Ulrich Schall, Martin Schalling, Peter R. Schofield, Thomas G. Schulze, Laura J. Scott, Rodney J. Scott, Alessandro Serretti, Jordan W. Smoller, Alessio Squassina, Eli A. Stahl, Eystein Stordal, Fabian Streit, Patrick F. Sullivan, Gustavo Turecki, Arne E. Vaaler, Eduard Vieta, John B. Vincent, Irwin D. Waldman, Cynthia S. Weickert, Thomas W. Weickert, David C. Whiteman, Martin Alda, Roel A. Ophoff, Kevin S. O’Connell, Niamh Mullins, Andreas J. Forstner, Maria Grigoroiu-Serbanescu, Howard J. Edenberg, Francis J. McMahon, Ole A. Andreassen, Arianna Di Florio, Andrew McQuillin, the Bipolar Disorder Working Group of the Psychiatric Genomics Consortium

**Affiliations:** Division of Psychiatry, University College London, London, UK; Human Genetics Branch, Intramural Research Program, National Institute of Mental Health, NIH, US Dept of HHS, Bethesda MD USA; Department of Psychiatry and Behavioral Sciences, SUNY Downstate Health Sciences University, Brooklyn, NY, USA; Institute for Genomics in Health, SUNY Downstate Health Sciences University, Brooklyn, NY, USA; NORMENT Centre, Institute of Clinical Medicine, University of Oslo and Division of Mental Health and Addiction, Oslo University Hospital, Oslo, Norway; Center for Precision Psychiatry, University of Oslo, Oslo, Norway; Department of Clinical Science, University of Bergen, Bergen, Norway; Centre for Neuropsychiatric Genetics and Genomics, Division of Psychological Medicine and Clinical Neurosciences, Cardiff University, Cardiff, UK; Stanley Center for Psychiatric Research, Broad Institute of MIT and Harvard, Cambridge, MA, USA; Analytic and Translational Genetics Unit, Department of Medicine, Massachusetts General Hospital, Boston, MA, USA; Department of Human Genetics, David Geffen School of Medicine, University of California Los Angeles, Los Angeles, CA, USA; Institute of Human Genetics, University of Bonn, School of Medicine and University Hospital Bonn, Bonn, Germany; Department of Psychiatry and Psychotherapy, University of Marburg, Marburg, Germany; Centre for Neuroimaging, Cognition and Genomics, School of Biological and Chemical Sciences and School of Psychology, University of Galway, Galway, Ireland; Department of Psychiatry, Icahn School of Medicine at Mount Sinai, New York, NY, USA; Charles Bronfman Institute for Personalized Medicine, Icahn School of Medicine at Mount Sinai, New York, NY, USA; Department of Genetics and Genomic Sciences, Icahn School of Medicine at Mount Sinai, New York, NY, USA; Global Computational Biology and Data Sciences, Boehringer Ingelheim Pharma GmbH and Co. KG, Biberach an der Riß, Germany; Department of Neurology, Klinikum rechts der Isar, School of Medicine, Technical University of Munich, Munich, Germany; Max Planck Institute of Psychiatry, Munich, Germany; VA NY Harbor Healthcare System, Brooklyn, NY, USA; Department of Epidemiology and Biostatistics, School of Public Health, SUNY Downstate Health Sciences University, Brooklyn, NY, USA; Department of Quantitative Health Sciences, Mayo Clinic, Rochester, MN, USA; Department of Psychiatry, University of California San Diego, La Jolla, CA, USA; Department of Psychiatry and Psychotherapy, Charité - Universitätsmedizin, Berlin, Germany; Social, Genetic and Developmental Psychiatry Centre, King’s College London, London, UK; NIHR Maudsley BRC, King’s College London, London, UK; Mental Health Department, University Regional Hospital, Biomedicine Institute (IBIMA), Málaga, Spain; Institute of Psychiatric Phenomics and Genomics (IPPG), LMU University Hospital, LMU Munich, Munich, Germany; Department of Psychiatry and Psychotherapy, University Hospital, LMU Munich, Munich, Germany; New York University, New York, NY, USA; Brain and Mental Health, QIMR Berghofer Medical Research Institute, Brisbane, QLD, Australia; School of Biomedical Sciences, Faculty of Medicine, The University of Queensland, Brisbane, QLD, Australia; Flatiron Institute, New York, NY, USA; Department of Psychiatry, Dalhousie University, Halifax, NS, Canada; KG Jebsen Centre for Neurodevelopmental disorders, University of Oslo, Oslo, Norway; Division of Psychiatry, Centre for Clinical Brain Sciences, University of Edinburgh, Edinburgh, UK; Department of Child and Adolescent Psychiatry/Psychology, Erasmus MC Sophia Children Hospital, Erasmus University, Rotterdam, The Netherlands; Psychiatry, Brain Center UMC Utrecht, Utrecht, The Netherlands; Department of Psychology Education and Child Studies, Erasmus School of Social and Behavioral Sciences, Erasmus University Rotterdam, The Netherlands; University Hospital of Psychiatry and Psychotherapy, University of Bern, Switzerland; Department of Neuroscience, Istituto Di Ricerche Farmacologiche Mario Negri IRCCS, Milano, Italy; Instituto de Salud Carlos III, Biomedical Network Research Centre on Mental Health (CIBERSAM), Madrid, Spain; Department of Psychiatry, Hospital Universitari Vall d’Hebron, Barcelona, Spain; Psychiatric Genetics Unit, Group of Psychiatry Mental Health and Addictions, Vall d’Hebron Research Institut (VHIR), Universitat Autònoma de Barcelona, Barcelona, Spain; Department of Psychiatry and Behavioral Neuroscience, University of Chicago, Chicago, IL, USA; Northwestern University, Chicago, IL, USA; National and Kapodistrian University of Athens, 2nd Department of Psychiatry, Attikon General Hospital, Athens, Greece; Department of Psychiatry and Psychotherapy, University Hospital Carl Gustav Carus, Technische Universität Dresden, Dresden, Germany; Programa SJD MIND Escoles, Hospital Sant Joan de Déu, Institut de Recerca Sant Joan de Déu, Esplugues de Llobregat, Spain; K. G. Jebsen Center for Genetic Epidemiology, Department of Public Health and Nursing, Faculty of Medicine and Health Sciences, Norwegian University of Science and Technology, Trondheim, Norway; Department of Psychiatry, Psychosomatic Medicine and Psychotherapy, University Hospital Frankfurt, Frankfurt am Main, Germany; Psychiatry, University of California San Francisco, San Francisco, CA, USA; School of Biomedical Sciences and Pharmacy, The University of Newcastle, Callaghan, NSW, Australia; Precision Medicine Research Program, Hunter Medical Research Institute, New Lambton, NSW, Australia; Section of Psychiatry, Department of Medical Sciences and Public Health, University of Cagliari, Italy; Department of Psychiatry and Forensic Medicine, Universitat Autònoma de Barcelona, Barcelona, Spain; Fundació Privada d’Investigació Sant Pau (FISP), Barcelona, Spain; Department of Psychiatry, Mood Disorders Program, McGill University Health Center, Montreal, QC, Canada; Division of Psychiatry, University of Edinburgh, Edinburgh, UK; Department of Physiology and Pharmacology, Karolinska Institutet, Stockholm, Sweden; Department of Psychiatry, Universidad Autonoma de Nuevo Leon, Monterrey, Mexico; Department of Psychiatry & Psychology, Mayo Clinic, Rochester, MN, USA; Department of Psychiatry, Laboratory of Psychiatric Genetics, Poznan University of Medical Sciences, Poznan, Poland; Center for Multimodal Imaging and Genetics, Departments of Neurosciences, Radiology, and Psychiatry, University of California, San Diego, CA, USA; Medical University of Graz, Division of Psychiatry and Psychotherapeutic Medicine, Graz, Austria; Department of Psychiatry and Behavioral Sciences, Johns Hopkins University School of Medicine, Baltimore, MD, USA; Department of Child and Adolescent Psychiatry, Psychosomatics and Psychotherapy, University Hospital Essen, University of Duisburg-Essen, Duisburg, Germany; Department of Medical Genetics, Oslo University Hospital Ullevål, Oslo, Norway; Department of Psychiatry, University of Arizona College of Medicine-Phoenix, Phoenix, AZ, USA; Carl T. Hayden Veterans Affairs Medical Center, Phoenix, AZ, USA; Banner-University Medical Center, Phoenix, AZ, USA; Academic Psychiatry, Newcastle University, Newcastle upon Tyne, UK; Department of Genetic Epidemiology in Psychiatry, Central Institute of Mental Health, Medical Faculty Mannheim, Heidelberg University, Mannheim, Germany; Institute of Clinical Medicine, University of Oslo, Oslo, Norway; Center for Neurobehavioral Genetics, Semel Institute for Neuroscience and Human Behavior, Los Angeles, CA, USA; Department of Psychiatry and Biobehavioral Science, Semel Institute, David Geffen School of Medicine, University of California, Los Angeles, Los Angeles, CA, USA; Department of Psychological Sciences, University of Missouri, Columbia, MO, USA; Genetics and Computational Biology, QIMR Berghofer Medical Research Institute, Brisbane, QLD, Australia; Psychological Medicine, University of Worcester, Worcester, UK; Institute for Translational Psychiatry, University of Münster, Münster, Germany; Department of Medical Epidemiology and Biostatistics, Karolinska Institutet, Stockholm, Sweden; Department of Psychology, Eberhard Karls Universität Tübingen, Tubingen, Germany; Department of Biomedicine, University of Basel, Basel, Switzerland; Institute of Medical Genetics and Pathology, University Hospital Basel, Basel, Switzerland; Brain and Mind Centre, The University of Sydney, Sydney, NSW, Australia; Univ Paris Est Créteil, INSERM, IMRB, Translational Neuropsychiatry, Créteil, France; Institute of Neuroscience and Physiology, University of Gothenburg, Gothenburg, Sweden; Campbell Family Mental Health Research Institute, Centre for Addiction and Mental Health, Toronto, ON, Canada; Neurogenetics Section, Centre for Addiction and Mental Health, Toronto, ON, Canada; Department of Psychiatry, University of Toronto, Toronto, ON, Canada; Institute of Medical Sciences, University of Toronto, Toronto, ON, Canada; Department of Psychiatry and Neurobehavioral Science, University College Cork, Cork, Ireland; Department of Psychiatry, Psychosomatics and Psychotherapy, Center of Mental Health, University Hospital Würzburg, Würzburg, Germany; Human Genetics Institute of New Jersey, Rutgers University, Piscataway, NJ, USA; ISGlobal, Barcelona, Spain; Department of Psychiatry, Erasmus MC, University Medical Center Rotterdam, Rotterdam, The Netherlands; Translational Psychiatry, Department of Molecular Medicine and Surgery, Karolinska Institutet, Stockholm, Sweden; Center for Molecular Medicine, Karolinska University Hospital, Stockholm, Sweden; Psychiatry, Northeast London NHS Foundation Trust, Ilford, UK; Clinic for Psychiatry and Psychotherapy, University Hospital Cologne, Cologne, Germany; School of Biomedical Sciences, Queensland University of Technology, Brisbane, QLD, Australia; Department of Translational Research in Psychiatry, Max Planck Institute of Psychiatry, Munich, Germany; Division of Psychiatry, Centre for Clinical Brain Sciences, The University of Edinburgh, Edinburgh, UK; Department of Psychiatry and Psychotherapy, University of Bonn, School of Medicine and University Hospital Bonn, Bonn, Germany; Research/Psychiatry, Veterans Affairs San Diego Healthcare System, San Diego, CA, USA; Unit of Clinical Psychiatry, University Hospital Agency of Cagliari, Cagliari, Italy; Department of Pharmacology, Dalhousie University, Halifax, Nova Scotia, Canada; National and Kapodistrian University of Athens, Medical School, Clinical Biochemistry Laboratory, Attikon General Hospital, Athens, Greece; Department of Clinical Neuroscience, Karolinska Institutet, Stockholm, Sweden; Centre for Psychiatry Research, SLSO Region Stockholm, Sweden; Department of Psychiatry, University of Michigan, Ann Arbor, MI, USA; Genetic Cancer Susceptibility Group, International Agency for Research on Cancer, Lyon, France; Institute for Genomic Health, SUNY Downstate Medical Center College of Medicine, Brooklyn, NY, USA; Department of Psychiatry and Psychotherapy, Central Institute of Mental Health, Medical Faculty Mannheim, University of Heidelberg, Mannheim, Germany; German Centre for Mental Health (DZPG), Germany; Department of Psychiatry and Psychotherapy, Clinical Division of General Psychiatry, Medical University of Vienna, Austria; Comprehensive Center for Clinical Neurosciences and Mental Health, Medical University of Vienna, Vienna, Austria; Centre for Neuroimaging and Cognitive Genomics (NICOG), School of Biological and Chemical Sciences, University of Galway, Galway, Ireland; Institute of Neuroscience and Medicine (INM-1), Research Centre Jülich, Jülich, Germany; Population Health, QIMR Berghofer Medical Research Institute, Brisbane, QLD, Australia; Department of Biomedical Sciences, University of Cagliari, Italy; Oxford Health NHS Foundation Trust, Warneford Hospital, Oxford, UK; Department of Psychiatry, University of Oxford, Warneford Hospital, Oxford, UK; Department of Psychiatry and Behavioral Sciences, Emory University School of Medicine, Atlanta, GA, USA; Outpatient Clinic for Bipolar Disorder, Altrecht, Utrecht, The Netherlands; Department of Psychiatry, Washington University in Saint Louis, Saint Louis, MO, USA; Department of Biochemistry and Molecular Biology II, Faculty of Pharmacy, University of Granada, Granada, Spain; Institute of Neurosciences ‘Federico Olóriz’, Biomedical Research Center (CIBM), University of Granada, Granada, Spain; Instituto de Investigación Biosanitaria ibs.GRANADA, Granada, Spain; German Center for Mental Health (DZPG), partner site Munich/Augsburg, 80336, Munich, Germany; Faculty of Medicine, University of Queensland, Brisbane, QLD, Australia; Psychiatry and the Behavioral Sciences, University of Southern California, Los Angeles, CA, USA; Department of Genetics, Microbiology, and Statistics, Faculty of Biology, Universitat de Barcelona, Barcelona, Spain; SAMRC Unit on Risk and Resilience in Mental Disorders, Dept of Psychiatry and Neuroscience Institute, University of Cape Town, Cape Town, South Africa; Department of Environmental Epidemiology, Nofer Institute of Occupational Medicine, Lodz, Poland; Neuroscience Research Australia, Sydney, NSW, Australia; Discipline of Psychiatry and Mental Health, School of Clinical Medicine, Faculty of Medicine and Health, University of New South Wales, Sydney, NSW, Australia; Centro de Biología Molecular Severo Ochoa, Universidad Autónoma de Madrid and CSIC, Madrid, Spain; Department of Psychiatry, Harvard Medical School, Boston, MA, USA; School of Biomedical Science and Pharmacy, University of Newcastle, Newcastle, NSW, Australia; Department of Psychiatry and Human Behavior, School of Medicine, University of California, Irvine, CA, USA; Psychiatry, Psychiatrisches Zentrum Nordbaden, Wiesloch, Germany; Department of Psychological Medicine, Institute of Psychiatry, Psychology and Neuroscience, King’s College London, London, UK; South London and Maudsley NHS Foundation Trust, Bethlem Royal Hospital, Monks Orchard Road, Beckenham, Kent, UK; Department of Clinical Sciences, Psychiatry, Umeå University Medical Faculty, Umeå, Sweden; Institute of Environmental Medicine, Karolinska Institutet, Stockholm, Sweden; Department of Psychiatry, University of Münster, Münster, Germany; Department of Psychiatry, Melbourne Medical School, The University of Melbourne, Melbourne, VIC, Australia; The Florey Institute of Neuroscience and Mental Health, The University of Melbourne, Parkville, VIC, Australia; Université Paris Cité, INSERM, Optimisation Thérapeutique en Neuropsychopharmacologie, UMRS-1144, Paris, France; APHP Nord, DMU Neurosciences, GHU Saint Louis-Lariboisière-Fernand Widal, Département de Psychiatrie et de Médecine Addictologique, Paris, France; Psychiatry, University of Pennsylvania, Philadelphia, PA, USA; Center for Statistical Genetics and Department of Biostatistics, University of Michigan, Ann Arbor, MI, USA; University of Queensland, Brisbane, QLD, Australia; Neuropsychiatric Genetics Research Group, Dept of Psychiatry and Trinity Translational Medicine Institute, Trinity College Dublin, Dublin, Ireland; National and Kapodistrian University of Athens, 1st Department of Psychiatry, Eginition Hospital, Athens, Greece; Estonian Genome Centre, Institute of Genomics, University of Tartu, Tartu, Estonia; Neuroscience Therapeutic Area, Janssen Research and Development, LLC, Titusville, NJ, USA; School of Biomedical Sciences, Faculty of Medicine and Health, University of New South Wales, Sydney, NSW, Australia; Department of Human Genetics, University of Chicago, Chicago, IL, USA; Department of Psychiatry, Department of Psychiatric Genetics, Poznan University of Medical Sciences, Poznan, Poland; School of Medicine and Public Health, University of Newcastle, Newcastle, NSW, Australia; Department of Medical Biochemistry and Biophysics, Karolinska Institutet, Stockholm, Sweden; JRD Data Science, Janssen Research and Development, LLC, Titusville, NJ, USA; Cancer Epidemiology and Prevention, M. Sklodowska-Curie National Research Institute of Oncology, Warsaw, Poland; University of Newcastle, Newcastle, NSW, Australia; Department of Psychiatry, Amsterdam University Medical Center, Amsterdam, The Netherlands; Department of Psychiatry and Neuropsychology, School for Mental Health and Neuroscience, Maastricht University Medical Center, Maastricht, The Netherlands; School of Psychology, The University of Queensland, Brisbane, QLD, Australia; Department of Psychiatry and Genetics Institute, University of Florida, Gainesville, FL, USA; Research Institute, Lindner Center of HOPE, Mason, OH, USA; School of Psychology and Faculty of Medicine, The University of Queensland, Brisbane, QLD, Australia; School of Psychology and Counselling, Queensland University of Technology, Brisbane, QLD, Australia; Division of Mental Health and Addiction, University of Oslo, Institute of Clinical Medicine, Oslo, Norway; Department of Mental Health, Faculty of Medicine and Health Sciences, Norwegian University of Science and Technology (NTNU), Trondheim, Norway; Psychiatry, St Olavs University Hospital, Trondheim, Norway; HudsonAlpha Institute for Biotechnology, Huntsville, AL, USA; Munich Cluster for Systems Neurology (SyNergy), Munich, Germany; University of Liverpool, Liverpool, UK; Medical and Population Genetics, Broad Institute, Cambridge, MA, USA; Stanley Center for Psychiatric Research, Broad Institute, Cambridge, MA, USA; Analytic and Translational Genetics Unit, Massachusetts General Hospital, Boston, MA, USA; Psychiatry, Indiana University School of Medicine, Indianapolis, IN, USA; Division of Psychiatry, Haukeland Universitetssjukehus, Bergen, Norway; Faculty of Medicine and Dentistry, University of Bergen, Bergen, Norway; Department of Clinical Neuroscience and Center for Molecular Medicine, Karolinska Institutet at Karolinska University Hospital, Solna, Sweden; Human Genetics and Computational Biomedicine, Pfizer Global Research and Development, Groton, CT, USA; Melbourne Neuropsychiatry Centre, Department of Psychiatry, The University of Melbourne, VIC, Australia; Monash Institute of Pharmaceutical Sciences (MIPS), Monash University, Parkville, VIC, Australia; Rutgers Health, Rutgers University, Piscataway, New Jersey, USA; University of Patras, School of Health Sciences, Department of Pharmacy, Laboratory of Pharmacogenomics and Individualized Therapy, Patras, Greece; United Arab Emirates University, College of Medicine and Health Sciences, Department of Genetics and Genomics, Al-Ain, United Arab Emirates; United Arab Emirates University, Zayed Center for Health Sciences, Al-Ain, United Arab Emirates; Erasmus University Medical Center Rotterdam, Faculty of Medicine and Health Sciences, Department of Pathology, Clinical Bioinformatics Unit, Rotterdam, The Netherlands; Center for Quantitative Health and Department of Psychiatry, Massachusetts General Hospital, Boston, MA, USA; Department of Neurology and Neurosurgery, McGill University, Faculty of Medicine, Montreal, QC, Canada; Montreal Neurological Institute and Hospital, McGill University, Montréal, QC, Canada; Centre for Brain and Mental Health Research, The University of Newcastle, Newcastle, NSW, Australia; Hunter Medical Research Institute, New Lambton Heights, NSW, Australia; Department of Psychiatry and Psychotherapy, University Medical Center Göttingen, Göttingen, Germany; Department of Psychiatry and Behavioral Sciences, SUNY Upstate Medical University, Syracuse, NY, USA; Pritzker Neuropsychiatric Disorders Research Consortium, University of Michigan, USA; The School of Biomedical Sciences and Pharmacy, Faculty of Medicine, Health and Wellbeing, University of Newcastle, Newcastle, NSW, Australia; Cancer Detection and Therapies Program, Hunter Medical Research Institute, University of Newcastle, Newcastle, NSW, Australia; Department of Medicine and Surgery, Kore University of Enna, Enna, Italy; Oasi Research Institute-IRCCS, Troina, Italy; Department of Psychiatry, Massachusetts General Hospital, Boston, MA, USA; Psychiatric and Neurodevelopmental Genetics Unit (PNGU), Massachusetts General Hospital, Boston, MA, USA; Department of Psychiatry, Hospital Namsos, Namsos, Norway; Department of Neuroscience, Norges Teknisk Naturvitenskapelige Universitet Fakultet for naturvitenskap og teknologi, Trondheim, Norway; Hector Institute for Artificial Intelligence in Psychiatry, Central Institute of Mental Health, Medical Faculty Mannheim, Heidelberg University, Mannheim, Germany; Department of Genetics, University of North Carolina at Chapel Hill, Chapel Hill, NC, USA; Department of Psychiatry, University of North Carolina at Chapel Hill, Chapel Hill, NC, USA; Department of Psychiatry, McGill University, Montreal, QC, Canada; Dept of Psychiatry, Sankt Olavs Hospital Universitetssykehuset i Trondheim, Trondheim, Norway; Clinical Institute of Neuroscience, Hospital Clinic, University of Barcelona, IDIBAPS, CIBERSAM, Barcelona, Spain; Department of Psychology, Emory University, Atlanta, GA, USA; Department of Neuroscience, SUNY Upstate Medical University, Syracuse, NY, USA; National Institute of Mental Health, Klecany, Czech Republic; Centre for Human Genetics, University of Marburg, Marburg, Germany; Biometric Psychiatric Genetics Research Unit, Alexandru Obregia Clinical Psychiatric Hospital, Bucharest, Romania; Department of Biochemistry, Molecular Biology and Pharmacology, Indiana University School of Medicine, Indianapolis, IN, USA; Department of Medical and Molecular Genetics, Indiana University, Indianapolis, IN, USA

**Author notes:** Corresponding Author: Tracey van der Veen, University College London, Gower Street, London, WC1E 6BT, UK. A list of members and affiliations appears in the Supplement. These authors contributed as first authors: Tracey van der Veen, Markos Tesfaye. These authors contributed as second authors: Jessica Mei Kay Yang, Toni Boltz, Friederike S. David, Shane Crinion, Maria Koromina. These authors jointly supervised this work: Martin Alda, Roel A. Ophoff, Kevin S. O’Connell, Niamh Mullins, Andreas J. Forstner, Maria Grigoroiu-Serbanescu, Howard J. Edenberg, Francis J. McMahon, Ole A. Andreassen, Arianna Di Florio, Andrew McQuillin.

## Abstract

Bipolar disorder’s (BD) clinical heterogeneity has an unresolved genetic basis. We meta-analyzed genome-wide association studies (GWAS) of 16 BD subphenotypes in 226,032 individuals from 57 cohorts (38,022 cases); 10 advanced to multivariate and multi-trait analyses. Four factors (compulsive, psychotic, dysregulated, internalizing) explained 82.8% of shared genetic variance. BD1 and BD2 loaded on distinct factors despite a high genetic correlation; 87.0% of common-factor loci were significant in neither subtype. Unipolar mania aligned with psychosis over internalizing, and was distinguishable from BD1, and rapid cycling showed heritable cross-domain liability. We identified 356 risk loci, 158 novel, including the first univariate-GWAS associations for psychosis, unipolar mania, rapid cycling and schizoaffective disorder—and 249 credible genes (89 high-confidence), 12 with approved-drug or clinical-phase annotations. Cell-type association showed a midbrain dopaminergic–GABAergic gradient along the psychotic factor. **BD’s genetic architecture appears hierarchical—a general liability resolving into dimensions of course and comorbidity beyond diagnostic subtypes.**

## INTRODUCTION

**Bipolar disorder (BD) is among the most complex and devastating psychiatric illnesses, yet the features that most shape its course, comorbidity and treatment response—psychosis, mania, rapid cycling, substance use and suicidality—vary so widely between patients that collapsing them into one or two diagnoses averages away the very dimensions on which prognosis and treatment hinge.** BD has been characterized genetically at the diagnostic-subtype level (bipolar disorder I, BD1; bipolar disorder II, BD2)^1–3^ and at the cross-disorder level^4–8^, yet the BD liability explained remains insufficient for diagnostic prediction in the general population^3^. BD’s heritability may concentrate within subphenotypes—narrower features (course specifiers) with more homogeneous signals^9,10^—that are familial, associating differentially with suicidality^11^, lithium response^12^, and age at onset^13^. Pooling these subphenotypes within BD1, BD2, or across BD averages their variant effects below detectio

BD is genetically complex by construction. Three architectures compose this heterogeneity: shared common-variant risk across diagnostic subtypes^1,3,14^, subphenotype-specific liability beneath them^9^, and shared common-variant risk with other psychiatric disorders^4–6,8^. A single-phenotype design captures one architecture and forfeits the other two. Mapping BD’s biology requires resolving all three jointly.

BD1 is more strongly genetically correlated with schizophrenia (SCZ), whereas BD2 is more strongly correlated with major depressive disorder (MDD), potentially indicating different proportions of shared liability rather than distinct disorders^1,3,14,15^. Large-scale clinical data show BD2 as more complex than BD1 in clinical course and comorbidity^16^.

However, clinical specifiers carry distinct polygenic signatures: psychosis is associated with elevated SCZ polygenic risk score (PRS), mania with elevated BD PRS, and depression with higher MDD PRS^17–19^, with rapid cycling indexing cross-diagnostic liability^20^. Yet genomic studies have resolved risk mostly at the level of disorders, subtypes^2^ or cross-disorder analyses^5–8^, leaving clinical heterogeneity within BD genetically unmapped.

Here we resolve BD below the diagnostic subtype, establishing, to our knowledge, the first univariate genome-wide-significant loci for psychosis, unipolar mania, rapid cycling and schizoaffective disorder.

Ten subphenotypes with sufficient power advanced to multivariate factor analysis, common-factor GWAS, and cross-disorder analyses, spanning three diagnostic subtypes (BD1; BD2; SZA, schizoaffective disorder bipolar type), three course specifiers (PSY, psychosis; UM, unipolar mania; RC, rapid cycling), and four psychiatric comorbidities (PAN, panic disorder; OCD, obsessive-compulsive disorder; SUD, alcohol or substance use disorder; and SA, suicide attempt).

This study resolves BD heterogeneity into a hierarchical genetic architecture—a general liability resolving into dimensions of course and comorbidity beyond subtype categories, informing stratification, nosology, and drug-targeting.

## RESULTS

### Subphenotype heritability and structured shared liability

We performed separate GWAS meta-analyses for the 16 BD subphenotypes across 57 of 79^3^ European ancestry cohorts, with deep-phenotyped data (38,022 cases; 188,010 healthy controls) using the standard Psychiatric Genomics Consortium (PGC) pipeline^21^. **Extended Data Fig. 1** summarizes the analytic workflow. Contributing cohorts obtained informed consent and local ethics approval (**Supplementary Table 1**). Subphenotype definitions and harmonization, prevalences, missingness, inclusions/exclusions, covariates, GWAS quality-control (QC) and cohort descriptions, are detailed in **Supplementary Tables 1-7**; **Supplementary Notes 1–2** and **Methods**.

The ten univariate subphenotype GWAS were carried forward to multivariate genomic structural equation modeling (Genomic SEM)^6^ and Multi-trait analysis of GWAS (MTAG)^22^; six subphenotypes were excluded due to low heritability and multicollinearity (**Supplementary Note 1.4**). Linkage Disequilibrium Score Regression (LDSC)^23^ intercepts (1.002–1.047; s.e.=0.007– 0.015) indicated predominantly polygenic signal (**Supplementary Table 2; Supplementary Note 2.3**). The 10 retained subphenotypes showed significant single-nucleotide polymorphism (SNP)heritability (h²SNP, OCD=0.102 to SZA=0.266; s.e.=0.007–0.048), on the liability scale (**Fig. 1**).

**Fig. 1.**
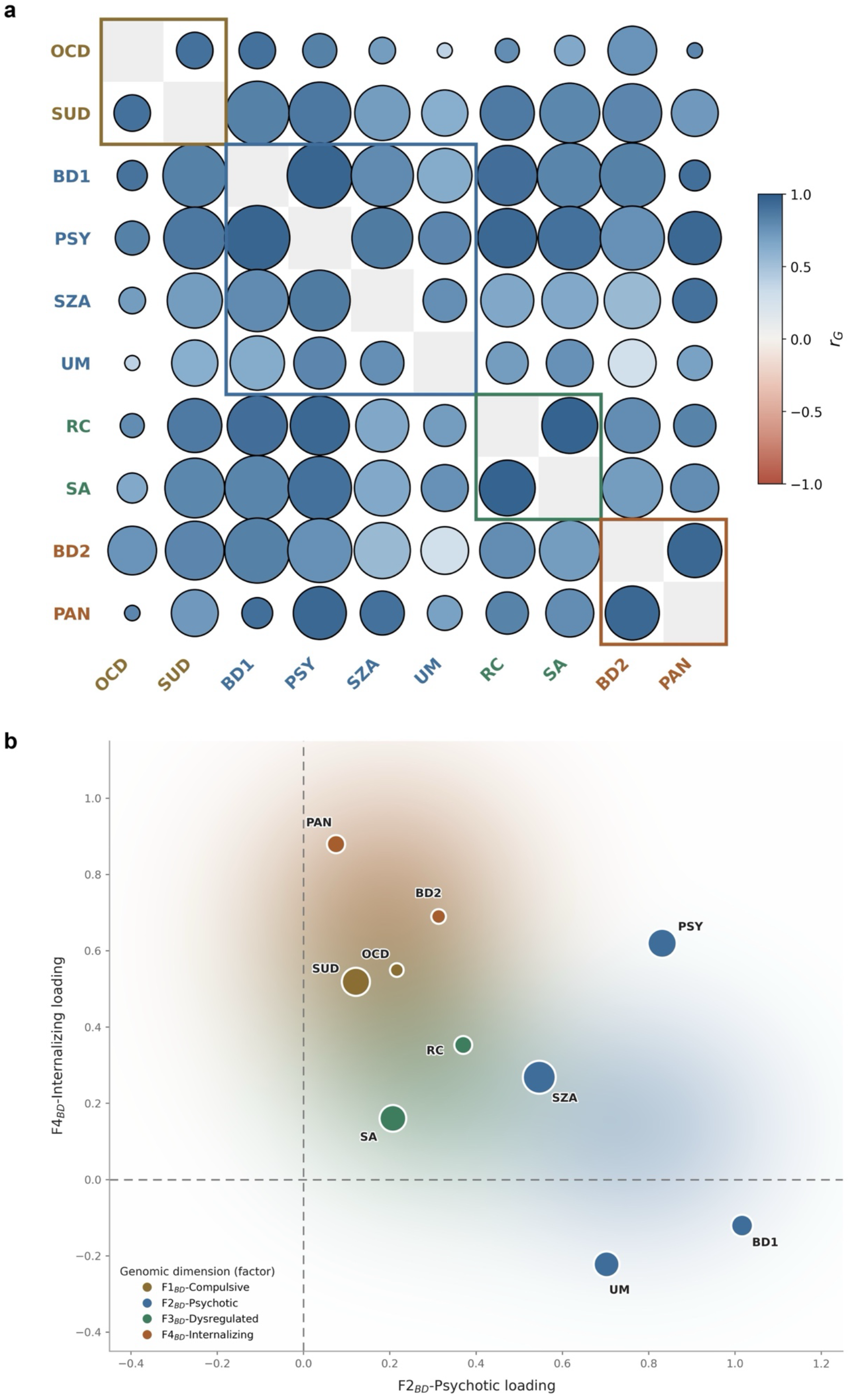
Genomic dimensions of the bipolar spectrum. **a.** Pairwise genetic correlations (*r*_G_) among the 10 subphenotypes (**Supplementary Table 8**), ordered and outlined by their assigned genomic dimension (colored blocks) resolved in Fig. 1b; axis labels are colored also by dimensions. Circle color denotes *r*_G_ direction and circle size denotes precision (larger circles indicate smaller standard error (s.e.) of *r*_G_); the diagonal is omitted. All pairwise *r*_G_ are Bonferroni-significant (*P* < 1.11 × 10⁻³). **b.** Standardized factor loadings from the exploratory factor analysis (EFA) of 10 bipolar disorder subphenotypes, positioning each subphenotype by its loading on the F2_BD_-Psychotic dimension (x-axis) and the F4_BD_-Internalizing dimension (y-axis) (**Supplementary Table 11**). Points are colored by their assigned genomic dimension from the four-factor model—F1_BD_- Compulsive, F2_BD_-Psychotic, F3_BD_-Dysregulated, F4_BD_-Internalizing—and sized by liability-scale SNP-based heritability (**Supplementary Table 2**). The shaded background blends the four-dimension colors to indicate their approximate regions; dashed lines mark zero loadings. The full four-factor structure and inter-factor correlations are shown in Fig. 3. *n* = 226,032 individuals (38,022 cases, 188,010 healthy controls).

Pairwise bivariate LDSC showed strong, positive genetic correlations among the 10 subphenotypes (*r*_G_ 0.280–0.966; Bonferroni-corrected, *P* < 1.11 × 10⁻³; **Fig. 1a, Supplementary Table 8**) — shared structure that motivated dimensionality reduction. Exploratory factor analysis (EFA) of this correlation structure resolved the subphenotypes into four genomic dimensions (**Fig. 1b**), and the correlations followed these dimensional boundaries. BD1 correlated most strongly with PSY (*r*_G_=0.956, s.e.=0.010) and BD2 with PAN (*r*_G_=0.940, s.e.=0.041)—each exceeding the BD1– BD2 correlation (*r*_G_=0.829, s.e.=0.010). The psychotic features were strongly correlated with one another (PSY–SZA, *r*_G_ = 0.859, s.e. = 0.022) and with BD1, and UM aligned more with PSY (*r*_G_ = 0.809, s.e. = 0.047) than BD1 (*r*_G_ = 0.630, s.e. = 0.040). The course and comorbidity pairs correlated. RC with SA was the strongest correlation (*r*_G_=0.966, s.e.=0.036), and SUD with OCD (*r*_G_=0.905, s.e.=0.090) exceeding either’s correlation with both subtypes.

Phenome-wide genetic architecture (*r*_G_) was shared across the 10 subphenotypes and 1,409 external traits, with BD suicide ideation (SI) included as a positive control (which, as expected, correlates with the subphenotypes, especially suicide attempt). Each of the four factors showed a distinguishable *r*_G_ signature (**Supplementary Table 9**). Along the psychotic–internalizing axis, internalizing-trait overlap—anxiety (ANX), MDD, post-traumatic stress disorder (PTSD) and borderline personality disorder (BPD)—declined from PAN and BD2 through BD1 to SZA and PSY, and near-zero at UM (*r*_G_ 0.51–0.60 at BD2, −0.12 to 0.03 at UM), while SCZ overlap was broadly shared (*r*_G_ =0.475–0.725, s.e.=0.018–0.093; *P* < 2.63 × 10⁻⁴) (**Supplementary Fig. 1**).

Family-history and cognition correlations crossed factor boundaries: positive with severe depression in first-degree relatives, and inverse with cognitive performance. BD1 showed a positive association with educational attainment, consistent with BD’s positive correlation^1,3^. Together, the within-BD and phenome-wide correlations resolve four genetically distinguishable dimensions arrayed along a psychotic–internalizing gradient (**Fig. 2; Supplementary Table 10)**.

**Fig. 2.**
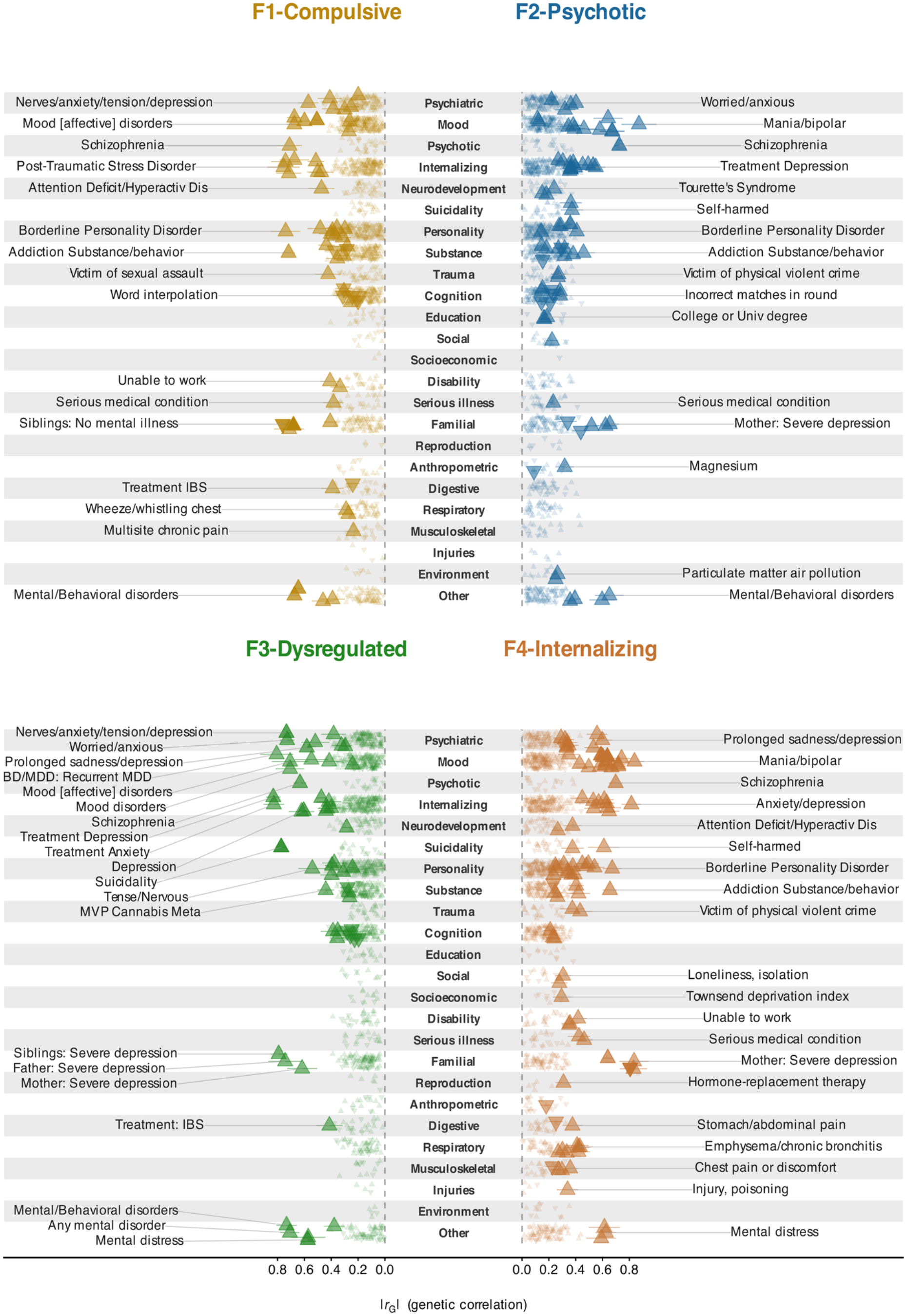
Phenome-wide genetic correlations between bipolar disorder dimension and external traits. LDSC genetic correlations (*r*_G_) between each of the 10 BD subphenotypes and 1,409 curated external traits (**Supplementary Table 10**). Subphenotypes are grouped into their four factor dimensions for convenience, defined by the factor model (Fig. 1b, Fig. 3). For each external trait within a panel, the plotted triangle shows the strongest |*r*_G_| among that factor’s subphenotypes. The central spine lists phenotype categories; horizontal distance from the spine encodes |*r*_G_|, the most strongly correlated traits lie farthest out. Upward triangles (▴) mark positive *r*_G_, downward (▾) negative *r*_G_; fill opacity scales with significance (−log₁₀(*P*)); Bonferroni-significant correlations (*P* < 3.55 × 10⁻⁵). Labeled traits are the strongest-|*r*_G_| trait per phenotype category. Horizontal whiskers span ±1 standard error. Seven cognitive traits are oriented so higher values indicate higher cognitive performance (Matrix = Matrix Pattern Completion task; Memory = Memory – Pairs Matching Test; RT = Reaction Time; Symbol Digit = Symbol Digit Substitution Task; Trails-B = Trail Making Test – B; Tower = Tower Rearranging Task; VNR = Verbal Numerical Reasoning Test)^92^ (**Supplementary Table 10)**. *n* = 226,032 individuals (38,022 cases, 188,010 healthy controls).

### Genomic SEM resolves the bipolar spectrum into four factors

The four-factor latent structure provided the best fit among the models evaluated using Genomic SEM. EFA (**Fig. 1b**) supported four factors over two- to five-factor, bifactor, and common-factor alternatives; only standardized loadings > 0.30 were carried into confirmatory factor analysis (CFA). The four-factor CFA solution was well-fitted with Comparative Fit Index (CFI)=0.976 and Standardized Root Mean Square Residual (SRMR)=0.071; SRMR < 0.10 is considered an acceptable fit^6^. This explained 82.8% of shared genetic variance (**Supplementary Tables 11-12**).

The four factors were (1) F1_BD_-Compulsive (OCD, SUD), (2) F2_BD_-Psychotic (BD1, PSY, SZA, UM), (3) F3_BD_-Dysregulated (RC, SA), and (4) F4_BD_-Internalizing (PAN, BD2) (**Fig. 3**). These factor labels were grounded in psychopathology taxonomy (**Supplementary Note 3.1**). Sensitivity analyses establishing factor number, separability, and stability are detailed in **Supplementary Note 3.2**.

**Fig. 3.**
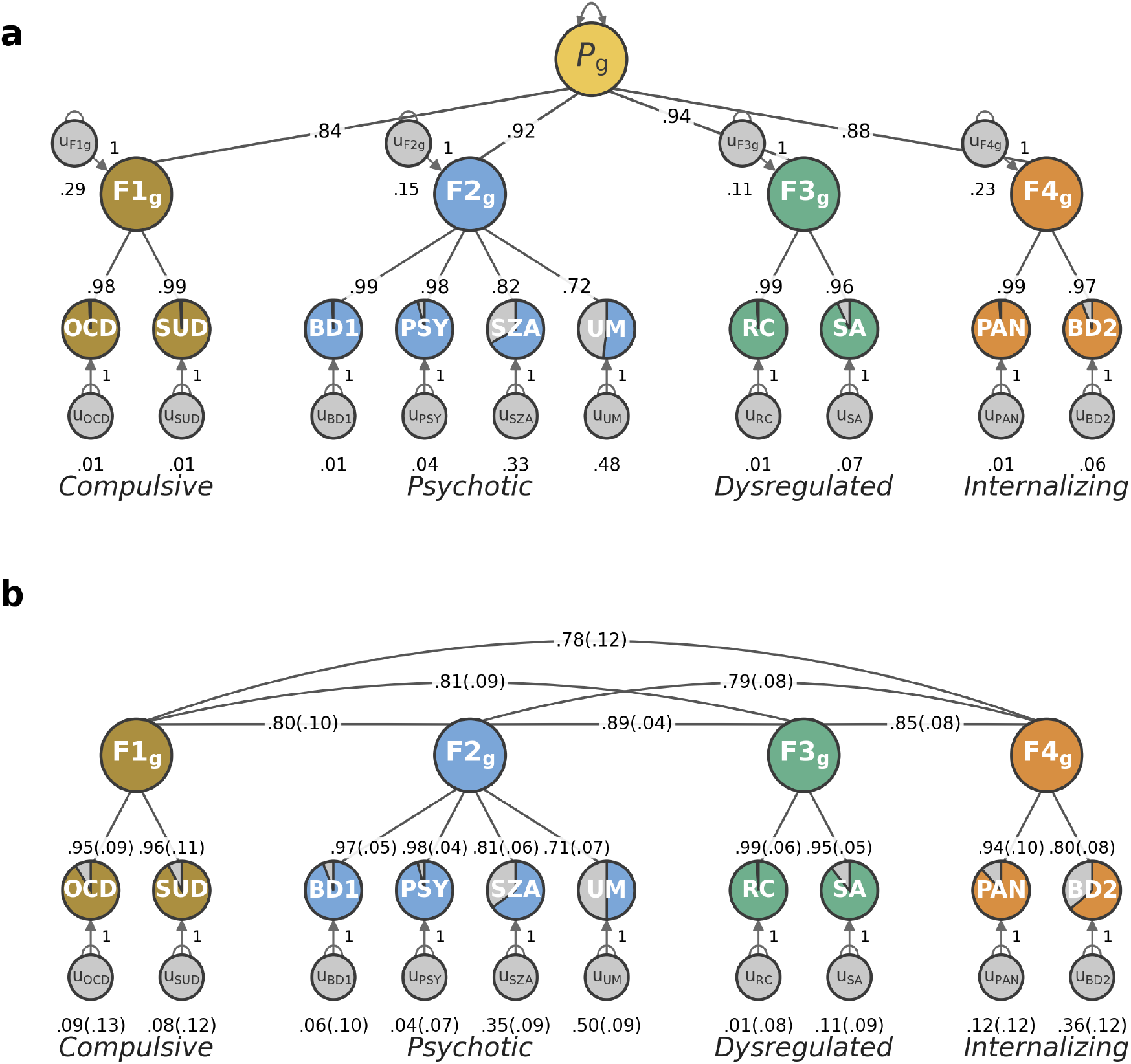
Modeling bipolar disorder heterogeneity with 10 subphenotypes. **a.** Standardized factor loadings from the higher-order *p*-factor hierarchical model. The common-factor (*P*g; yellow) represents the general bipolar spectrum liability shared across the four first-order factors (F1–F4): F1_BD_-Compulsive (gold), F2_BD_-Psychotic (blue), F3_BD_-Dysregulated (green), F4_BD_-Internalizing (orange). Path coefficients are standardized factor loadings; all estimates are standardized relative to SNP-based heritability. Residual variance (U, gray circles) reflects variance not explained by the factor solution. **b**. Inter-factor genetic correlations from the correlated four-factor model (standard errors in parentheses), ranging from 0.778 to 0.892 within the BD spectrum. Factor composition: F1_BD_-Compulsive (OCD, SUD), F2_BD_-Psychotic (BD1, PSY, SZA, UM), F3_BD_-Dysregulated (RC, SA), and F4_BD_-Internalizing (PAN, BD2). The four-factor model explained 82.8% of shared genetic variance. Full model fit indices and parameter estimates are in **Supplementary Tables 11-12**. *n* = 226,032 individuals (38,022 cases, 188,010 healthy controls).

The factor structure followed the psychotic–internalizing axis established in the genetic correlations above: BD1, PSY, SZA, and UM co-loaded on F2_BD_-Psychotic, while PAN and BD2 jointly loaded on F4_BD_-Internalizing—the subtypes loading separately despite their strong pairwise BD1–BD2 *r*_G_. Inter-factor correlations were high (0.778–0.892). As these factors partition a single disorder rather than distinct disorders, they share more common bipolar liability and correlate more strongly than cross-disorder factors^24^; separability rests on bifactor rejection, distinct loadings, and SNP-level heterogeneity. RC loaded broadly across all four factors, consistent with its pairwise *r*_G_ with other subphenotypes (**Supplementary Table 8, 11**).

With factor-level GWAS feasible only at moderate inter-factor correlations, our multivariate SNP discovery used the Genomic SEM common-factor GWAS and Q_SNP_ heterogeneity test^8^; established methods for estimating shared and heterogeneous liability across highly correlated factors^6,25,26^. The common-factor model underperformed the four-factor solution, yet it still had an acceptable fit (SRMR=0.089 vs 0.071, both less than 0.10; **Supplementary Table 12**).

### Cross-disorder embedding preserves the psychotic dimension

We next asked how these dimensions sit within the broader psychiatric landscape: which remain distinct axes when modeled alongside other disorders, and which resolve into facets of cross-disorder (_CD_) factors. We embedded the 10 subphenotypes among 10 external psychiatric traits— selected for their BD associations^1,3,4,14,24^. The 10 external traits were—SCZ^27^, MDD^28^, anorexia nervosa (AN)^29^, Tourette’s syndrome (TS)^30^, autism spectrum disorder (ASD)^31^, attention deficit/hyperactivity disorder (ADHD)^32^, BPD^33^, ANX^34^, PTSD^35^, and risk-taking behavior (RISK)^36^. This identified a five-factor cross-disorder model (CFI=0.974; SRMR=0.083; an acceptable fit; **Fig. 4; Supplementary Table 12**).

**Fig. 4.**
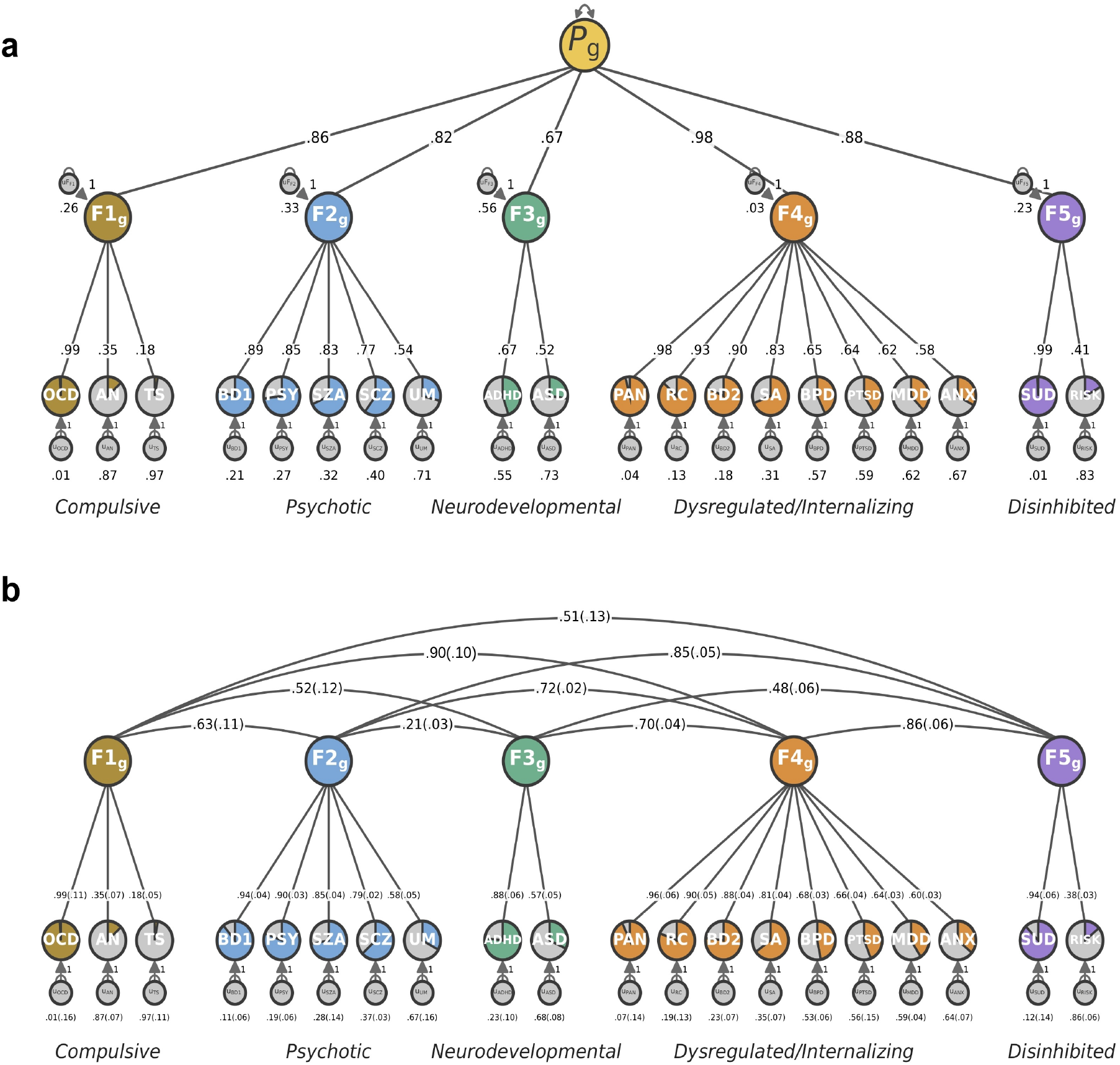
Cross-disorder modeling with 10 bipolar disorder subphenotypes. **a**. Standardized factor loadings from the higher-order *p*-factor hierarchical model in the cross-disorder framework, with 10 BD subphenotypes (Fig. 3). Subphenotypes were embedded alongside 10 external psychiatric and behavioral traits (AN, TS, SCZ, ASD, ADHD, BPD, ANX, MDD, PTSD, and RISK). The common-factor *P*g (yellow) represents the cross-disorder liability shared across the five first- order factors (F1–F5), colored by domain: F1_CD_-Compulsive (gold), F2_CD_-Psychotic (blue), F3_CD_- Neurodevelopmental (green), F4_CD_-Dysregulated/Internalizing (orange), and F5_CD_-Disinhibited (purple). All estimates are standardized relative to SNP-based heritability. Residual variance (U, gray circles) reflects variance not explained by the factor solution. **b**. Inter-factor genetic correlations from the correlated five-factor model in the same 20-trait framework (standard errors in parentheses). The five-factor model explained 69.1% of shared genetic variance under transdiagnostic embedding, reduced from 82.8% in the bipolar-restricted framework; the BD four-factor structure dispersed across the five factors. F4_CD_-Dysregulated/Internalizing loaded strongest contrasting with F2_BD_-Psychotic and F3_BD_-Dysregulated in the bipolar-restricted framework. Full model fit indices are in **Supplementary Tables 11-12**. *n* = 226,032 individuals (38,022 cases, 188,010 healthy controls).

The five factors comprised (1) F1_CD_-Compulsive (OCD_BD_, AN, TS); (2) F2_CD_-Psychotic (BD1_BD_, PSY_BD_, SZA_BD_, SCZ, UM_BD_); (3) F3_CD_-Neurodevelopmental (ADHD, ASD); (4) F4_CD_-Dysregulated/Internalizing (PAN_BD_, RC_BD_, BD2_BD_, SA_BD_, BPD, PTSD, MDD, ANX); and (5) F5_CD_-Disinhibited (SUD_BD_, RISK). F2_BD_-Psychotic mapped intact to F2_CD_-Psychotic, joined by SCZ, replicating the psychotic dimension’s robustness to broader psychiatric structure^24^. In contrast, F1_BD_-Compulsive is divided between F1_CD_-Compulsive (OCD) and F5_CD_-Disinhibited (SUD). F3_BD_-Dysregulated and F4_BD_-Internalizing, merged with cross-disorder internalizing traits (BPD, ANX, MDD, PTSD) onto F4_CD_-Dysregulated/Internalizing, indicating shared latent internalizing liability across BD and cross-disorders.

The variance explained by the models was 82.8% in the bipolar-restricted model and 69.1% under cross-disorder embedding, a difference expected under the larger cross-disorder covariance matrix, with BD-specific structure most fully recovered (**Supplementary Note 3.4**).

### The psychotic–internalizing axis persists at the locus level

Global correlation (*r*_G_) averages signals across the genome and can mask divergence at specific loci. We therefore applied local analysis of [co]variant association (LAVA)^37^ to partition genome-wide *r*_G_ into 2,495 approximately independent LD blocks, identifying where genetic overlap between subphenotypes and external traits concentrates, and where it diverges (see **Methods**). We detected Bonferroni-significant local correlations across 176 loci, comparing 11 subphenotypes (with SI as positive control) against 19 external traits (**see Supplementary Note 4.1**).

The psychotic–internalizing axis extended to our locus level *r*_G_ across the subphenotypes: psychotic traits dominated BD1’s local overlap—with the most Bonferroni-significant regions between BD1–PSY (**Fig. 5**) and BD1–SCZ **(Supplementary Tables 13–16)**. Internalizing traits accounted for a greater share of BD2’s false discovery rate (FDR)-significant overlap (22.0%) than BD1’s (9.6%; nominal Fisher *P* = 0.029)—consistent with BD1’s psychotic-dominated local overlap. Local correlation was distributed along these two factors, rather than localized at BD1-BD2 specific regions. Two convergent ‘hotspots’ harbored significant local *r*_G_ across subphenotype pairs: 21q22.13 (*KCNJ6*) and 3p22.2 (*TRANK1*) (**Supplementary Tables 13-16; Supplementary Note 4.2-4.4**).

**Fig. 5.**
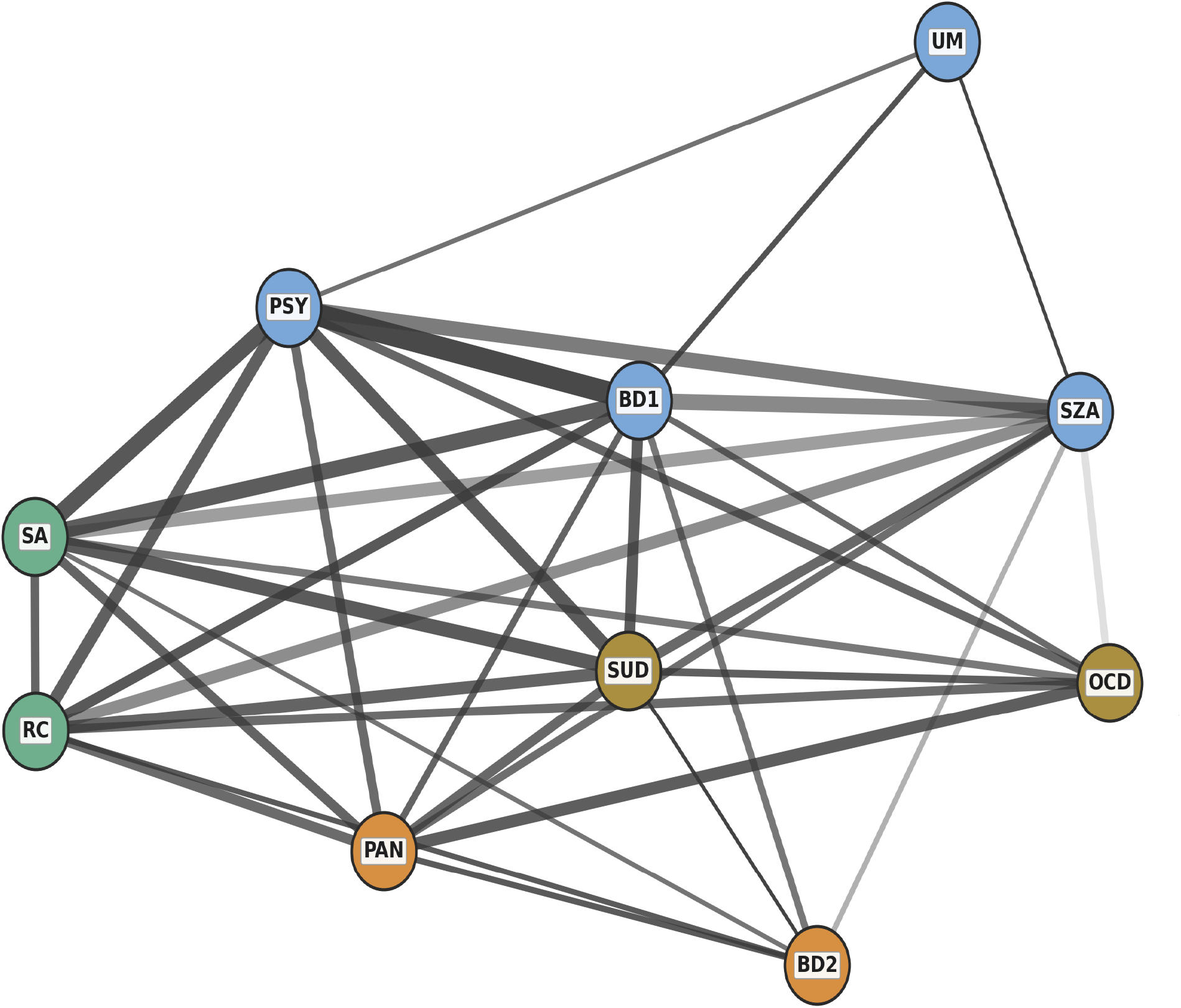
Local genetic correlations across 10 bipolar disorder subphenotypes. Nodes (circles) represent the 10 BD subphenotypes, colored by factor for convenience (see Fig. 3). Edges (lines) connect subphenotype pairs with FDR-significant local genetic correlation (ρ; FDR < 0.05); edge width is proportional to the number of FDR-significant loci and edge opacity to the magnitude of the average local genetic correlation. FDR-significant BD1-BD2 local correlations emerged, no Bonferroni-significant, despite the relatively high global *r*_G_ (0.829, s.e.=0.01), and these were mostly pleiotropic: BD1’s local correlations were strongest with PSY not BD2, consistent with factor-differential pleiotropy (F2_BD_-Psychotic versus F4_BD_-Internalizing) rather than shared subtype architecture. See **Supplementary Tables 13-16** and **Supplementary Note 4.3**. *n* = 226,032 individuals (38,022 cases, 188,010 healthy controls).

### Genome-wide discovery across subphenotypes identifies novel risk loci

Subphenotype-resolved discovery yielded 356 unique, independent genome-wide significant (GWS) loci, including to our knowledge the first univariate-GWAS associations for psychosis, unipolar mania, rapid cycling, and schizoaffective disorder. Univariate GWAS identified 60 GWS (*P* < 5.0×10^−8^) associations across BD subphenotypes—31 novel and 29 replicating prior BD associations (**Supplementary Table 7**). The strongest univariate associations implicated *TRANK1* for PSY (rs9834970, 3p22.2; *P*=5.99×10^−14^)^1,38–40^, *IMMP2L* for UM (rs2613588, 7q31.1; *P*=4.14×10^−8^)^14^, *TMEM130–TRRAP–NPTX2* for RC (rs113396024, 7q22.1; *P*=1.61×10^−10^)^1,3,14^, and SZA at *SCN3B/GRAMD1B* (rs3851101, 11q24.1; *P*=1.54×10^−9^) previously sub-threshold^14^. For BD2, we replicated the earlier identified lead variant at *SLIT3* (rs7720655, 5q35.1; *P*=4.21×10^−8^)^1^, and an additional novel locus upstream of *PCDH7* (rs61795936, 4p15.1; *P*=4.68×10^−8^).

Integration across analyses yielded 356 unique, independent GWS loci—158 (44.4%) were novel at the subphenotype level (no prior GWS association for the respective subphenotype) in NHGRI-EBI GWAS Catalog^41^-indexed studies (accessed 2026-04-02). Per-analysis contributions were: 60 univariate (**Supplementary Note 5.1.1**), 162 Genomic SEM common-factor (**Fig. 6**; **Supplementary Table 17**), 12 Q_SNP_ (single-SNP heterogeneity) (**Supplementary Table 18**), and 148 MTAG associations (**Supplementary Tables 19-20**). Of the 162 common-factor loci, 141 (87.0%) reached GWS in neither BD1 nor BD2 (**Supplementary Table 17**). Of these, 76 (53.9%) were also not GWS within 500 kb in the largest univariate BD GWAS^3^, indicating novel discovery from the subphenotype-resolved common-factor model.

**Fig. 6.**
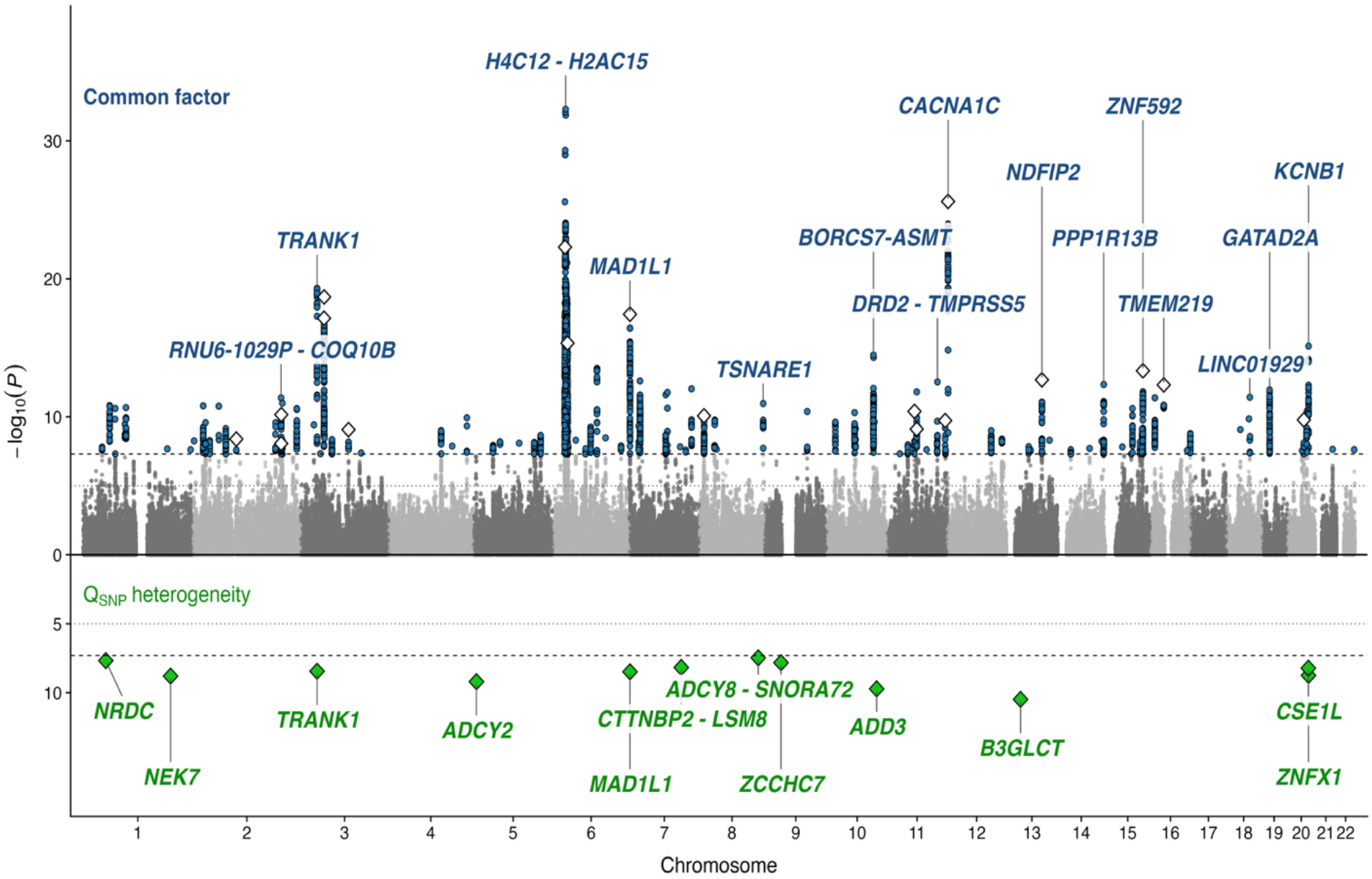
Multivariate genome-wide association analysis: common-factor and Q_SNP_ heterogene- ity loci. Miami plot of the BD subphenotype common-factor GWAS. Above the axis, the common-factor GWAS: background, all tested SNPs by chromosome (alternating gray); blue circles, common-factor genome-wide significant SNPs (*P*_CF_ < 5×10^−8^; **Supplementary Table 17**); white dia- monds, the top credible genes (of 249 in **Supplementary Table 21**) whose strongest lead SNP also reaches common-factor genome-wide significance, plotted at the lead SNP’s −log_10_(*P*_CF_). Below the axis (mirrored), the Q_SNP_ heterogeneity: green diamonds, the 12 loci with significant between-subphenotype heterogeneity (*P*_QSNP_ < 5×10^−8^; **Supplementary Table 18**), plotted at −log_10_(*P*_QSNP_). Q_SNP_ test is not for discordance rather whether a SNP’s effects across the factor’s indicators depart from what its effect through the common factor predicts. Italic gene labels mark the top common-factor hit per chromosome (blue, above) and each Q_SNP_ locus (green, below). Loci such as *TRANK1* and *MAD1L1* appear in both tracks at the same chromosomal position, at distinct lead SNPs—a signal shared through the common factor and a separate heterogeneous signal that departs from it, co-located in one locus. Threshold lines: dashed = genome-wide significance (*P* < 5×10^−8^); dotted = suggestive (*P* < 1×10^−5^). The credible genes are detailed in **Supplementary Table 2**1. *n* = 226,032 individuals (38,022 cases, 188,010 healthy controls).

Heterogeneity testing identified 12 additional loci that diverged from the common factor BD model (*P*_QSNP_ < 5×10^−8^), undetectable in the 10 univariate subphenotype input GWAS. These heterogeneity loci are shown in **Fig. 6** and **Supplementary Table 18**. Subphenotype Q_SNP_ effects were sign-discordant with SCZ and MDD, comparators outside common-factor/Q_SNP_ modeling.

Directional concordance was quantified by a two-sided binomial sign test of each trait’s per-locus effect signs against the consensus direction of the other subphenotypes. PSY was the most concordant (*P* < 0.001) and UM the least. Neither external comparator showed significant directional concordance (SCZ^27^and MDD^28^), with MDD trending inverse. High-confidence credible genes implicated in BD1–UM sign-discordant loci included *ADCY2*^42^ (rs78308718, 5p15.31; *P*_QSNP_ = 6.29 × 10^−10^) and *MAD1L1* (rs56180486, 7p22.3; *P*_QSNP_ = 3.27 × 10^−9^)^1,7,14^, while *TRANK1* remained concordant (**Supplementary Fig. 2; Supplementary Note 3.5**). This test spans only the 12 Q_SNP_ loci; therefore per-trait concordance ranking is descriptive rather than establishing directionality.

Subsequently, each of the 10 univariate subphenotype GWAS was pairwise MTAG- augmented with the largest, most recent BD GWAS^3^ to increase effective sample sizes producing 10 subphenotype-specific MTAG summary statistics. These served as improved inputs^22^ for gene-based association (**Supplementary Fig. 3**), gene prioritization, functional annotation, cell-type, tissue and developmental association, and polygenic risk estimation.

### Convergence prioritizes high-confidence credible genes

Consistent with convergence-based frameworks^3,43^, candidate genes were prioritized from each subphenotype’s resolved signal across six complementary tests (Evidence Score 0–6), yielding 249 credible genes (Evidence Score ≥ 2) across MAGMA^44^, Transcriptome-Wide Association Study (TWAS)-colocalization^45–47^, Summary-based Mendelian Randomization (SMR)^48^, ANNOVAR^49^, Functional Mapping and Annotation (FUMA)^50^ proximity, and rare-variant burden. Of these, 89 credible genes supported by three or more tests, were considered high confidence (HC; Evidence Score ≥ 3), of which 29 were robust (Evidence Score ≥ 4) with 25 shown in **Fig. 7** (**Supplementary Note 6.2; Supplementary Table 21**). *SP4* and *FADS1* replicated prior BD GWAS findings^1,3^, and *STK4*^1,42^ showed systematic SMR discordance against ADHD and BPD (**Supplementary Tables 22-23**; **Supplementary Note 6.4**).

**Fig. 7.**
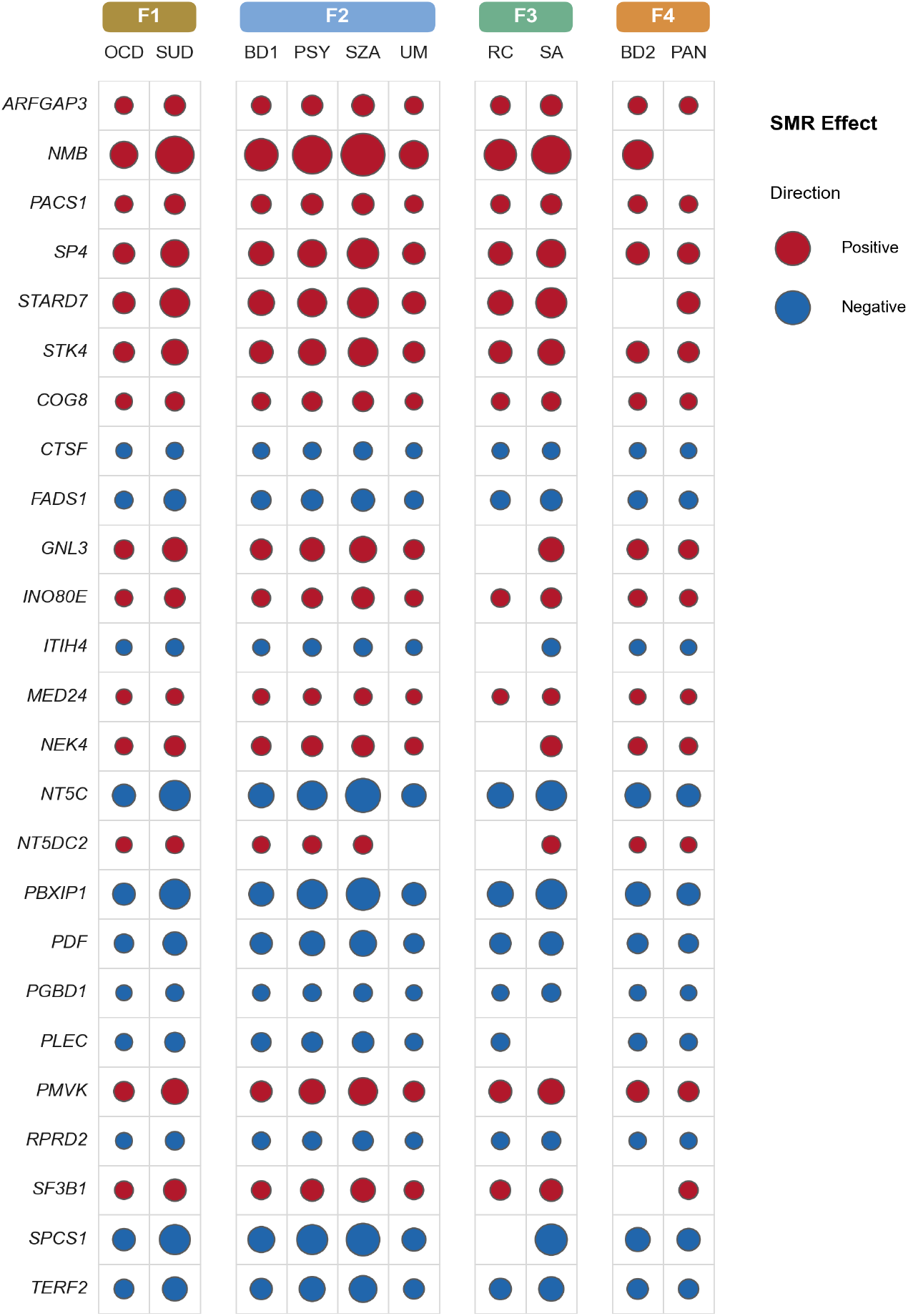
SMR associations of robust credible genes across the 10 bipolar disorder subphenotypes. Summary-based Mendelian randomization (SMR) associations of the 25 robust credible genes (Evidence Score ≥ 4) with available SMR data, ordered by descending Evidence Score; all 29 robust credible genes are listed in **Supplementary Table 21**. Subphenotype columns are grouped by genomic-SEM factor for convenience (Fig. 3). Each circle marks a Bonferroni-significant SMR association (*P*_SMR_ < 3.11 × 10^−6^; **Supplementary Table 22**); circle size is proportional to the SMR effect magnitude and color indicates direction (red, positive; blue, negative). A cell without a circle indicates no significant SMR association. For effect sizes, standard errors and *P* values, see **Supplementary Table 22**. *n* = 226,032 individuals (38,022 cases, 188,010 healthy controls).

Of the 249 credible genes, 238 mapped to OpenTargets (https://genetics.opentargets.org/; accessed 2026-03-29), of which 75 (31.5%) were small-molecule-tractable (*P*=1.79×10^−26^). Twelve carry approved-drug or clinical-phase annotations, including the high-confidence credible genes *CACNA1C*^51–53^ (12p13.33), *HDAC5*^1,14,54^ (17q21.31) and *KCNS1* (20q13.12). *STK4* was the only Evidence-Score-5 credible gene that is small-molecule-tractable at the structure-with-ligand level (**Supplementary Tables 24-25; Supplementary Note 6.5**).

### Cell-type association shows shared substrate and midbrain gradient

Tissue, pathway, and cell type ‘enrichment’ (referred to as association)^55^ converged exclusively on the brain (**Supplementary Tables 26-28**). Gene-based associations in FUMA reached significance in brain tissue only, spanning cortical, subcortical, cerebellar, and pituitary regions. Subphenotypes mostly shared substrates; although amygdala association reached Bonferroni significance in every subphenotype except UM (**Supplementary Table 26**). At the cell-type level, of 64 cell types reaching FDR significance, 24 reached pan-BD-spectrum significance, 40 showed differential association, and 16 were specific to one subphenotype; the 24 included gestational week (GW) 26 GABAergic neurons, GW 26 astrocytes, and L5.6 THEMIS deep-layer cortical pyramidal neurons. VIP interneurons were FDR-significant in seven of 10 subphenotypes (**Supplementary Table 28a-b**). Above this baseline, midbrain dopaminergic and GABAergic sub-classes (DA1, DA2, Gaba, NbGaba) showed a gradient along the psychotic factor, ascending SZA→PSY→UM→BD1, with deep-layer pyramidal neurons likewise strongest in BD1. Across midbrain subclasses BD2 exceeded SZA, PSY and UM, and matched or exceeded BD1 at all but DA1, despite BD1’s larger effective sample; UM likewise exceeded PSY despite its smaller size— inversions of the power ranking that support biological-specificity, rather than power differentials (**Supplementary Tables 28b, 3; Fig. 8a–b**), with standardized z-scores confirming both reversals (**Supplementary Table 28e**).

**Fig. 8.**
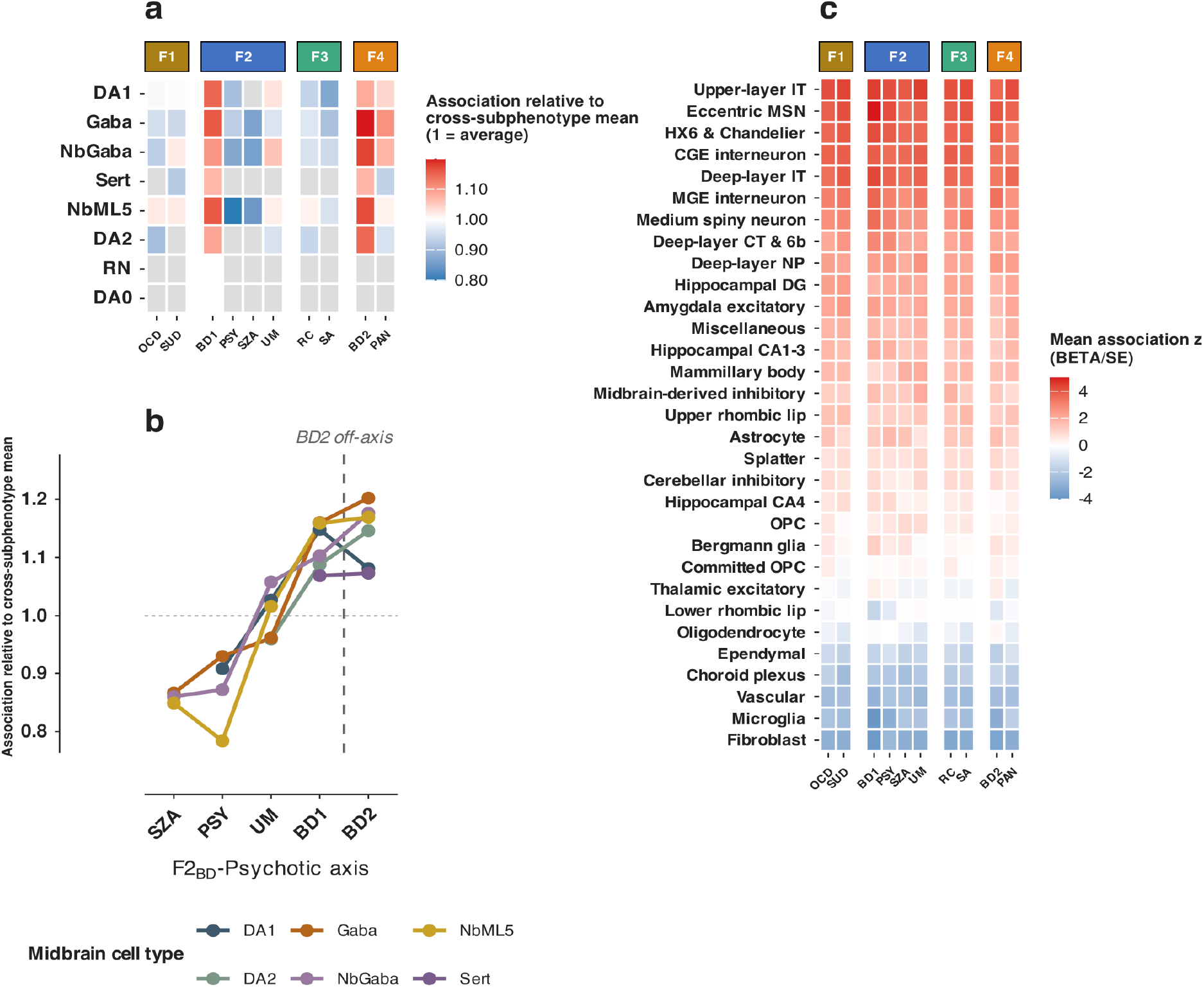
Cell-type association across 10 bipolar disorder subphenotypes. **a.** Midbrain neuronal subclasses show a psychotic gradient across the bipolar disorder spectrum. Heatmap of cell-type association relative to the cross-subphenotype mean (*z* ÷ mean *z* across the 10 subphenotypes per cell type; *z* = BETA/s.e.; **Supplementary Tables 28a-c**) for eight midbrain neuronal subclasses (see **Methods**). Color is centered at 1: red, above the cell type’s pan-spectrum average; blue, below; gray, not FDR-significant. The gradient is supported by sample-size-independent rank reversals and standardized values (sample-size-normalized *Z*, **Supplementary Table 28d**; within-sub- phenotype standardized *Z* and ranks, **Supplementary Table 28e**), not on absolute *z*. Subphenotype columns are grouped by factor (Fig. 3); rows are ordered by mean association. **b**. Midbrain association grades along the F2_BD_-Psychotic axis. Six midbrain neuronal subclasses (DA1, DA2, sequential dopaminergic maturation stages; Gaba, NbGaba, GABAergic neurons and neuroblasts; NbML5, mediolateral neuroblast 5; Sert, serotonergic), plotted as association relative to the cross- subphenotype mean, order by SZA→PSY→UM→BD1 along the F2_BD_-Psychotic axis. The dashed vertical line marks the F2_BD_-Psychotic loading boundary; BD2 (loads on F4_BD_-. Internalizing) is shown for comparison. The dotted horizontal line marks the cross-subphenotype average (ratio = 1) (**Supplementary Table 28d-e**; per-cell-type association values in **Supplementary Table 28b**). This direction replicated in the independent atlas: across midbrain-predominant clusters, UM and BD2 exceeded PSY. **c**. Telencephalic neurons carry the strongest shared association across all 10 subphenotypes in the adult human brain atlas. Heatmap of mean association *z* (*z* = BETA/s.e.) for each of the 31 superclusters, averaged across constituent clusters and ordered top to bottom by mean association. *Z* indexes strength of evidence; the shared signal is the consistent top ranking of telencephalic superclusters within every subphenotype (per-trait FDR < 0.05 across 461 clusters, **Supplementary Table 28f**). Red, positive association; blue, negative association. Subphenotype columns are grouped by factor (**Supplementary Table 28f**). *n* = 226,032 individuals (38,022 cases, 188,010 healthy controls).

An additional test of cell types in an adult human brain atlas,^56^ localized the strongest shared association to telencephalic neurons. Of 461 clusters, 228 were FDR-significant, 126 across all ten subphenotypes (**Fig. 8c, Supplementary Table 28f).** In the adult atlas, the direction of the gradient replicated: UM and BD2 exceeded PSY and SZA, UM despite lower power (Supplementary Note 6.1.1). Among glia, association localized to astrocytes, the sole glial population implicated across both reference sets; UM carried the most trait-specific singletons, although its broader cluster profile was shared across subphenotypes (**Supplementary Note 6.1.4**). Among populations represented in both reference panels, this signal resolved to cortical GABAergic interneurons (from the medial and caudal ganglionic eminences) and intratelencephalic projection neurons, recurring across all ten subphenotypes (**Supplementary Tables 28a, 28f**), indicating a neuronal basis shared across the bipolar spectrum.

The developmental association identified Bonferroni-significant signals at early mid-prenatal (PSY), late mid-prenatal (PSY, UM, SA, SUD), and late infancy (BD1, BD2) stages (**Supplementary Table 29**). Age-specific analyses showed four Bonferroni-significant associations at one year postnatal (BD1, BD2, SZA, PAN; **Supplementary Table 30**).

### Subphenotype polygenic scores stratify risk

PRS-CS-auto^57^ was applied to subphenotype-MTAG summary statistics in a leave-one-cohort-out framework, isolating each subphenotype’s PRS from its discovery cohorts. Individuals in the top one percent of polygenic risk showed absolute risks of 10.4–18.5% across the 10 subphenotypes. PRS achieved held-out liability R² between 0.080–0.142 (PAN to UM; s.e.=0.011–0.027) under weighted random-effects meta-analysis to account for between-cohort heterogeneity (**Supplementary Table 31; Supplementary Fig. 4**).

### Replication and rare-variant burden converge on novel loci

Four lines of evidence support these findings. (1) Within-BD recovery was high but is expected by construction—the subphenotype cohorts are a subset of, and MTAG incorporated, the prior BD GWAS—and is therefore reported as an internal consistency check, not independent replication. By exact lead-SNP match (**Supplementary Table 32, Table 32.1**), 87.1–94.7% of BD1, BD2, and SZA loci coincided with prior BD subtype-resolved GWAS, and 81.1–83.3% of PSY, UM, and RC loci with the aggregate BD GWAS^1,3,14,53^. Eight of the 10 subphenotypes were recovered at GWS within ±100 kb — BD1, BD2, SZA, PSY, UM, RC, OCD, and SUD — with SA additionally recovered within ±500 kb, and PAN below GWS, *P*=7×10⁻⁷; **Supplementary Table 32, Table 32.2**. (2) Independent cross-disorder replication was broad: 71.9–89.5% of each subphenotype’s lead SNPs overlapped independent GWAS of other psychiatric disorders by exact lead-SNP match at GWS (**Supplementary Note 5.2; Supplementary Table 32, Table 32.1**). (3) Internal multi-method convergence was evident, 208 of the 356 unique loci were not recovered by MTAG — independent of the borrowed BD-GWAS effects — of which 137 (65.9%) are novel relative to the previous BD GWAS within ±500 kb^1,3,14,53^ (**Supplementary Note 5.1**). (4) Rare-variant burden convergence occurred on five credible genes—three high-confidence (*SP4, PACS1, GIGYF1*) and two credible (*ZNF584, FAM83H*): *SP4* in Schizophrenia Exome Meta-Analysis (SCHEMA)^58^ (24,248 cases; *P*=0.040), and *PACS1, GIGYF1, ZNF584*, and *FAM83H* in Bipolar Exome Consortium^59^ (BipEx; 13,933 cases; *P*=4.67×10^−6^) (**Supplementary Table 33**).

Discovery analyses passed quality-control safeguards: LDSC intercepts remained near 1 (**Supplementary Table 2**; **Supplementary Fig. 5**), and chromosome-split analysis recovered the factor structure, supported by parallel analyses (**Supplementary Fig. 6**). MTAG maxFDR fell below 0.05 across subphenotypes, and univariate versus MTAG genetic-correlations correlated highly (Pearson *r*=0.887–0.987), indicating MTAG amplified each subphenotype’s own signal rather than substituting that of the larger BD GWAS (**Supplementary Fig. 7**).

## DISCUSSION

BD’s genomic architecture **appears** multi-dimensional, **resolvable** only at sufficient phenotypic resolution. Our findings suggest it is hierarchical: a general bipolar liability resolving into dimensions of clinical course and comorbidity. Within the subphenotypes analyzed, our dual framework resolved four candidate dimensions—compulsive, psychotic, dysregulated, internalizing—and which appear more BD-intrinsic versus shared with the psychiatric landscape. These dimensions capture within-BD heritability the BD1/BD2 dichotomy cannot: **87% of common-factor loci reached significance in neither subtype**. Although strongly inter-correlated, these factors remained distinguishable, with distinct subphenotype loadings, divergent external genetic correlations, and SNP heterogeneity.

The psychotic–internalizing dissociation emerged across analyses, newly resolved in subphenotype lcal genetic correlations. At the global genetic correlation level, BD1 retained moderate internalizing overlap. However, at the locus level, BD1 identified LAVA associations with MDD, PAN, PTSD, and BPD, not with ANX—consistent with BD-PRS burden being inversely associated with comorbid anxiety^60^.

Unipolar mania also aligned with psychosis over internalizing liability, showing reduced genetic correlations across internalizing traits—consistent with clinical^61,62^ and polygenic evidence of a lower internalizing burden and a distinguishable mania-predominant signal within BD^17,18,63^. The lead UM locus implicates *IMMP2L*, a mitochondrial inner-membrane peptidase; mitochondrial energetics was implicated in treatment-emergent mania^64^ and, through copy-number variants, to SCZ^65^—consistent with UM’s psychotic factor positioning. BD1–UM sign-discordant further implicated heterogeneous loci, including two high-confidence credible genes, *MAD1L1* and *ADCY2*.

Although UM falls outside current nosology, its genetically distinguishable signature is inconsistent with early or attenuated BD1 only. Pending replication, this divergence may help distinguish patients likely to retain a unipolar course from those predisposed to depressive conversion. However, as UM ascertainment depends on age, with some cases converting to depression later^62,66^, longitudinal validation is required.

RC bridged the psychotic–internalizing divide, loading across all four factors, inconsistent with its clinical characterization as transient^67–69^. Highest *r*_G_s were against SA, PAN, BD1 and PSY, indexing a broad spectrum burden^9,20,67,70^; under cross-disorder embedding, RC merged onto the Dysregulated/Internalizing factor alongside PAN and ANX, consistent with evidence of PAN and RC familial co-aggregation^71^, and PAN internalizing liability^72^. Rapid cycling indexes cross-domain liability, including suicide attempt—a potential marker for risk stratification, pending validation, consistent with elevated suicide risk in rapid-cycling patients^11,67,73^.

The lead RC locus (*TMEM130–TRRAP–NPTX2*) harbors *NPTX2*, a candidate BD biomarker: *NPTX2* contributes to synaptic organization, associates with SCZ^74,75^, and shows reduced cerebro-spinal fluid (CSF) levels in BD1^75,76^. The same locus harbors *TRRAP*, a chromatin remodeling factor with rare-variant ASD association^77^.

Despite a high pairwise genetic correlation, BD1 and BD2 loaded on distinct factors, paralleling correlations (BD1–SCZ, BD2–MDD)^2,15^, in which subtype’s genetic variance was more strongly shared with SCZ or MDD respectively, than with the other subtype. LAVA added locus-level resolution: BD1–BD2 shared variance was distributed across regions linked to their respective psychotic or internalizing genomic factors, rather than concentrated at subtype-specific loci, with internalizing traits accounting for a greater share of BD2’s overlap.

The 29 highest-confidence credible genes span transcriptional regulation, protein trafficking, chromatin remodeling, cytoskeletal organization, and cell cycle control.

*SP4*, a neuronal transcription factor, replicated the top BD GWAS finding^1,3^ in all 10 subphenotypes, with common and rare coding-variant support paralleling SCZ^27,58^. *FADS1*, the rate-limiting desaturase in long-chain polyunsaturated fatty acid (PUFA) biosynthesis^1,3,78^, reinforced lipid metabolism as a candidate shared pathway. *STK4*, a Hippo-pathway serine/threonine kinase, showed SMR discordance from ADHD and BPD across BD subphenotypes, paralleling a prior *STK4* BD-versus-MDD differentiation^42^; the signal was detectable only at the subphenotype-level. It maps to 20q13.12, where the approved-drug channel gene *KCNS1* lies; both genes were SMR- concordant with schizophrenia, aligning the locus with the psychotic axis. Twelve such credible genes carry approved-drug or clinical-phase annotations; such annotations indicate tractability, not proven efficacy in BD (**Supplementary Tables 34-36**).

Two local pleiotropic ‘hotspots’ dominated. The leading within-BD hotspot, 21q22.13, was a common-factor locus with concordant local correlations across subphenotypes—shared, not antagonistic, effects; its signal maps to *KCNJ6* (a G-protein-gated potassium channel in dopamine and reward circuitry^79,80^), though the locus did not resolve to a subphenotype-level credible gene. The second, 3p22.2, resolved to *TRANK1*—a high-confidence BD credible gene^38,40^ that spans psychotic-aligned subphenotypes and is secondarily associated with SCZ^81,82^, educational attainment^83^, and hypothyroidism^84^.

Tissue-specificity converged on the brain across all 10 subphenotypes. At the cell type level, prenatal GABAergic neurons, astrocytes and deep-layer cortical pyramidal neurons were FDR-.significant across all 10 subphenotypes, consistent with early neurodevelopmental programming as a shared foundation. Telencephalic neuron association replicated in the independent adult human brain atlas, while glial replication was selective and developmental-stage-specific in adult cortical astrocytes only.

Within the psychotic factor, midbrain association graded SZA→PSY→UM→BD1, while SZA’s strongest cell type, cortical excitatory populations, paralleled SCZ^27^ associations. This gradient is not driven by statistical power—at midbrain subclasses BD2 exceeded BD1, and UM exceeded PSY, each the smaller-N_EFF_ trait—and in the adult atlas UM again exceeded PSY, with BD1 and BD2 comparable despite BD1’s larger sample; though the full ordering and its dopaminergic peak remain a developmental-stage-specific observation.

Medium spiny neurons emerged in the atlas (**Supplementary Fig. 8**), replicating striatal association reported for schizophrenia, obsessive-compulsive disorder and cross-disorder analyses^27,43,85^, and extending the previous bipolar GWAS^1,3^, which implicated pyramidal and inhibitory neurons. VIP interneurons (inhibit other inhibitory neurons^86–88^) reached FDR significance across subphenotypes. The most recent BD GWAS reported association in enteroendocrine cells of the large intestine^3^— together raising the possibility of VIP involvement at both the brain and gut ends of the gut-brain axis.

Several limitations apply. Analyses were limited to European-ancestry cohorts with deep phenotyping and lacked in-sample LD estimates for all cohorts; multi-ancestry extension is a priority. Our analysis represents a two- to three-fold increase over prior subphenotype efforts^9,14^, however samples remain limited for those with lower population prevalences. Subphenotype-specific loci await replication in independent, comparably deep-phenotyped cohorts, partially mitigated by cross-disorder lead-SNP overlap, exome-burden convergence, and held-out PRS. Resolution and scale serve different functions: subphenotype-resolved signals emerged only through subphenotype-level designs. Three of the four factors are defined by two indicators, with precedence^24^; identification relies on the full correlated model, and is supported by within-factor genetic correlations and chromosome-split replication (**Supplementary Note 3.2**). Subphenotype loci carry shared and feature-specific liability, shown by Q_SNP_ heterogeneity, distinct loadings and divergent *r*_G_. Other genetic variation may contribute, requiring joint modeling of common and rare factors. Phenotypes were collected retrospectively, though longitudinal studies show low BD1– BD2 conversion. RC and UM depressive episodes were indexed as lifetime, not current, partially offsetting cross-sectional underestimation. Genomic SEM identifies covariance, not causation. Credible genes converged across analyses; however causal claims will require functional validation. Cross-trait assortative mating may affect *r*_G_ estimates^89^, though it alone is unlikely to produce the structured, factor-differential pattern of correlations we observed^90^. PRS were derived from largely clinically ascertained samples, with variance explained insufficient for clinical prediction^3,27,91^. Independent replication was not possible, as the analysis uses essentially all available BD-spectrum samples; robustness rested on out-of-sample PRS, independent external *r*_G_, and multi-method convergence. Sex-stratified analyses confirmed near-unity *r*_G_ (**Supplementary Note 1.10**), supporting combined-sex analysis. Finer sex-stratified subphenotype analyses await larger samples.

In summary, BD’s genetic architecture extends beyond BD1/BD2 into four dimensions of course and comorbidity—with psychosis, rapid cycling and unipolar mania genetically distinguishable—pairing shared spectrum liability with loci that differentiate subphenotypes. Because these dimensions **span BD’s most clinically consequential features—psychosis, mania, rapid cycling, substance use and suicidality—resolving them uncovers biology and candidate drug targets that pooled, diagnosis-level analyses cannot reach**.

---

**Extended Data Fig. 1.**
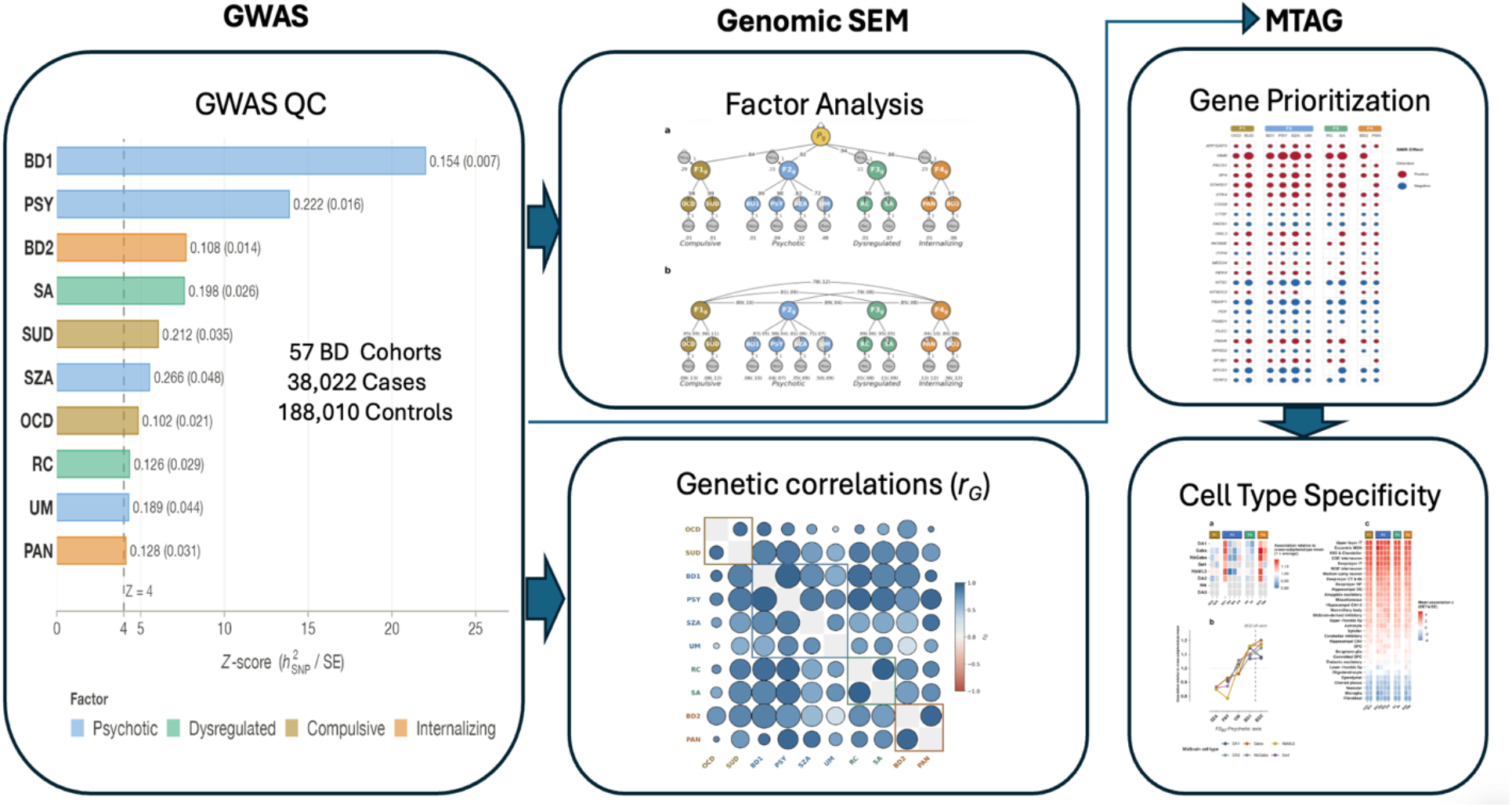
Schematic of the analytical workflow. GWAS summary statistics from 57 cohorts (226,032 individuals, 38,022 cases; 188,010 healthy controls; **Supplementary Tables 1 and 3**) covered 16 subphenotypes; 10 advanced to multivariate modeling, spanning diagnostic subtypes (BD1, bipolar I; BD2, bipolar II; SZA, schizoaffective bipolar type), illness course (PSY, psychosis within BD; UM, unipolar mania; RC, rapid cycling), and psychiatric comorbidities (PAN, panic; OCD, obsessive-compulsive; SUD, substance use; SA, suicide attempt). **Left:** heritability *z*-score for subphenotypes on the liability scale, sorted descending; bar color indicates four factor assignment. All 10 subphenotypes exceeded the z ≥ 4 multivariate adequacy threshold (**Supplementary Table 2**). **Center top**: Genomic SEM on the LDSC genetic covariance matrix identified four factors—F1_BD_-Compulsive, F2_BD_-Psychotic, F3_BD_-Dysregulated, and F4_BD_-Internalizing—explaining 82.8% of shared genetic variance (**Supplementary Tables 11-12**). **Center bottom**: LDSC genetic correlations between each subphenotype and 1,409 external benchmark traits, alongside the internal between-subphenotype correlations (*r*_G_) (**Supplementary Tables 9-10**). **Right top**: integration of univariate GWAS, Genomic SEM common-factor analysis, Q_SNP_ heterogeneity testing, and MTAG yielded 356 unique independent genome-wide significant loci (158 novel) and 249 prioritized credible genes (**Supplementary Tables 20-21**). **Right bottom**: cell type association against single-cell RNA-sequencing data identified shared prenatal GABAergic and glial substrates across all 10 subphenotypes, with factor-specific signals detected above this baseline (**Supplementary Table 28a–f**). Factor colors carry through main-text and supplementary figures: F1_BD_-Compulsive (gold), F2_BD_-Psychotic (blue), F3_BD_-Dysregulated (green), F4_BD_-Internalizing (orange), unless otherwise stated.

## METHODS

### Ethics

The lead principal investigator of each contributing cohort warranted that the study protocol was approved by their local ethics committee or institutional review board and that all participants provided written informed consent. All analyses in the present study were performed on de-identified, aggregated GWAS summary statistics. Details of each cohort, including ascertainment, diagnostic instruments, and the corresponding ethics approvals, are provided in **Supplementary Table 1** and **Supplementary Note 12**.

### Study participants and GWAS datasets

The primary dataset comprised 57 international, European ancestry cohorts contributing to the PGC Bipolar Disorder Working Group, with deep phenotyping data sufficient to derive harmonized phenotypes for multivariate genomic modeling. Fifty-two cohorts were internal to the PGC with access to individual genotypes. The five external cohorts provided phenotype-specific summary statistics. Twenty-two cohorts, included in O’Connell et al., 2025^3^ were excluded as they lacked item-level data for deep-phenotyping, required for harmonized bipolar disorder phenotype definitions. The study population comprised 226,032 individuals (cases: 38,022; healthy controls: 188,010), including an internal PGC analytic dataset with deep phenotyping of 52,222 genotyped individuals (17,908 cases; 34,314 healthy controls), used in *a priori* EFA modeling to genetic analyses. The total N_EFF_, equivalent to an equal number of cases and controls in each cohort, was calculated as 4 × n_CASES_ × n_CONTROLS_ / (n_CASES_ + n_CONTROLS_). Cases met DSM (Diagnostic and Statistical Manual)-IV^93^, DSM-5^94^, or ICD (International Classification of Diseases)-10^95^ criteria for a lifetime diagnosis of bipolar disorder (I, II, or schizoaffective type), including specifiers and comorbid disorders, established using semi-structured or structured clinical interviews, clinician-administered diagnostic checklists, or systematic medical record review conducted by trained interviewers. Of the 57 cohorts, 49 were clinically ascertained and eight were community-based. All analyses were restricted to individuals of European genetic ancestry. Ten harmonized bipolar phenotype GWAS were defined and carried forward across all analyses: bipolar I disorder (BD1) and bipolar II disorder (BD2) both with clinical and community ascertainment, schizoaffective disorder bipolar type, psychosis within bipolar disorder, unipolar mania (UM), rapid cycling, suicide attempt, panic disorder comorbidity, OCD comorbidity, and alcohol or substance use disorder comorbidity. Univariate GWAS case-control design contrasted individuals with a BD diagnosis and the phenotype, against healthy controls within each cohort. Sample overlap across subphenotypes, was corrected throughout by the LDSC bivariate cross-trait intercepts. GWAS cohort metadata, diagnostic definitions, subphenotype harmonization, missingness handling, and inclusion/exclusion criteria are in **Supplementary Tables 1–7**; **Supplementary Notes 1.1–1.7**.

### Terminology

We use the term “subphenotype” when referring to BD-specific characteristics, the BD subphenotypes (**Supplementary Note 1**), and “external trait” for traits external to BD including the 20 traits in the cross-disorder five-factor model. “Transdiagnostic” refers to a cross-factor shared signal captured by the *p*-factor—a hierarchical liability common across psychiatric disorders. “Cross-disorder” refers to analyses spanning multiple diagnostic categories—the 20-trait Genomic SEM model embedding 10 BD subphenotypes with 10 external traits, and independent locus replication and genetic correlations against external GWAS. See **Supplementary Note 1.3**, **Supplementary Table 4** for the list of harmonized subphenotypes and their definitions.

### Genotyping, quality control, and imputation

Genotyping was performed within each contributing cohort using standard genotyping arrays (see **Supplementary Table 1**). A centralized QC/imputation pipeline was performed using RICOPILI (Rapid Imputation for COnsortias PIpeLIne^21^) (version 2024_Nov_21.001). Standardized quality control, imputation, and statistical analyses were performed centrally. Quality control parameters for retaining SNPs and samples were SNP missingness <0.05 (before sample removal), sample missingness <0.02, autosomal heterozygosity deviation (|Fhet| < 0.2), SNP missingness < 0.02 (after sample removal), differential SNP missingness between cases and controls < 0.02, and SNP Hardy-Weinberg equilibrium (*P* >1.0 ×10^−10^ in cases, *P* >1.0 ×10^−6^ in controls). Sex mismatches were checked based on X chromosome F-statistic (female F < 0.2, male F > 0.8).

Relatedness was assessed across cohorts using identity-by-descent; one member of each pair with pi_hat > 0.2 was excluded, prioritizing exclusion of individuals related to the most others, controls over cases, and individuals from larger cohorts. Duplicate detection across cohorts was performed using genotype-based checksums. Population structure was assessed by principal component analysis (PCA) using EIGENSTRAT v6.1.4 (https://www.hsph.harvard.edu/alkes-price/software/)^96^, with samples projected onto the 1000 Genomes Phase 3 reference panel^97^; individuals falling more than six standard deviations from the European cluster centroid on PC1 and PC2 were excluded. Genotype imputation was performed using Eagle v2.3.5 (https://alkesgroup.broadinstitute.org/Eagle/)^98^ for pre-phasing followed by Minimac3 (https://genome.sph.umich.edu/wiki/Minimac3/)^3,99^ against the Haplotype Reference Consortium (HRC r1.0) reference panel^100^. Postimputation filtering retained variants with imputation quality score INFO ≥ 0.8; minor allele frequency (MAF) > 0.01, and minor allele count (MAC) > 10 in the smaller of the case or control groups^1^. Within-cohort genome-wide association analyses were conducted using additive logistic regression in PLINK v1.90 (https://www.cog-genomics.org/plink2/)^101^, conditioning on the first five ancestry principal components (PCs), as previously described^3^. Cohort-level summary statistics were combined using inverse-variance weighted fixed-effects meta-analysis in METAL (https://genome.sph.umich.edu/wiki/METAL_Documentation)^102^. Variants were required to be present in at least 50% of contributing cohorts and to represent at least 75% of total N_EFF_. DENTIST (https://github.com/Yves-CHEN/DENTIST)^103^ was applied in post-meta-analysis to detect and remove allelic inconsistencies.

GWS loci were defined as the region around a lead SNP (*P* < 5.0 × 10^−8^) encompassing SNPs in LD (r² > 0.1) within ±500 kb, using the HRC European reference panel^100^, following the locus definition used in comparable psychiatric and comorbidity GWAS^7,14,43^. This window is narrower than the ±3,000 kb of the primary BD meta-analysis (O’Connell et al.)^3^, whose larger effective sample size produces a broader spread of genome-wide-significant association around major loci. At the smaller effective sample sizes of the subphenotype GWAS, significant SNPs are expected to extend less far from the lead, so a narrower window is appropriate, while the r² > 0.1 criterion prevents adjacent independent signals from being merged.

The number of post-QC SNPs included across all 16 univariate BD GWAS is 3,706,932- 7,665,779 (**Supplementary Table 2**).

### SNP-heritability and genetic correlations

Sample-size differences and overlap were addressed through standard error-weighting in covariance-matrix methods (Genomic SEM, LDSC, LAVA, MTAG; **Supplementary Notes 2.3**) and through N-normalized *z*-scoring in cell type association. Prior to LDSC and Genomic SEM analyses, summary statistics were harmonized using the LDSC munge() function in LDSC, v1.0.1, restricting to approximately 1.2 million HapMap3^104^ SNPs, removing indels, structural and strand-ambiguous SNPs, and filtering variants on MAF < 0.01 or INFO < 0.9. SNP-based heritability (h^2^_SNP_) was estimated using LDSC with precomputed LD scores from 1000 Genomes Phase 3 European samples^97^. Standard bivariate LDSC was used for pairwise genome-wide genetic correlations among the 10 primary phenotypes; the multivariable version of LDSC^23^ implemented within Genomic SEM^6,8^ was used for all multivariate analyses. For binary phenotypes, h²SNP was converted to the liability scale^105^ using a population prevalence of 2% (bipolar disorder lifetime prevalence)^3^ and a sample prevalence of 0.5, the value required when the summed effective sample size across cohorts is supplied as the input sample size, which yields unbiased liability-scale heritability for meta-analyzed case/control data^106^. For the cohorts contributing summary statistics without accompanying case/control counts, the per-SNP effective sample size was estimated from the standard errors as N_EFF_ = 4 / (2·MAF·(1−MAF)·SE²) and bounded between 0.5 and 1.1 times the total effective sample size to prevent inflation from extremely small standard errors^107^.

Population prevalence for external traits used matched reporting in the corresponding literature for each trait. Subphenotype-specific population prevalences (*K*) followed two rules: diagnostic categories (BD1, BD2, schizoaffective; unipolar mania modeled as BD1) used the lifetime population prevalence of the diagnosis, and course specifiers and comorbidities used the joint phenotype, the base bipolar prevalence (2%) multiplied by the within-bipolar prevalence of the feature, since cases are bipolar patients carrying the feature contrasted against general-population controls. The cohort case proportions (**Supplementary Table 5**) entered only the per-cohort effective-sample-size calculation; the liability-scale conversion itself used a sample prevalence of 0.5 with the summed effective sample size. Univariate subphenotype associations were tested against healthy controls rather than BD cases (with subphenotype) to cases (without subphenotype), which avoids the collider bias of within-case (feature-versus-no-feature) contrasts; consequently, these loci index general bipolar liability together with feature-specific liability, and subphenotype-specific differentiation is established by the Q_SNP_, local and global genetic-correlation analyses.

Subphenotype and external traits were required to have a heritability *z*-score (h^2^_SNP_/SE) > 4, as genetic correlation estimates for traits with low heritability signal may be characterized by high noise and limited reliability^23^. For the 10 primary subphenotype GWASs, mean LDSC intercepts ranged from 1.002 to 1.047 (closer to 1), consistent with polygenic signals. A summary of subphenotype LDSC QC metrics including attenuation ratio range is provided in **Supplementary Table 6**; **Supplementary Note 2.3**.

Pairwise genome-wide genetic correlations (*r*_G_) were estimated using bivariate LDSC on the Complex-Traits Genetics Virtual Lab (CTG-VL) platform^108^. Cross-trait phenome-wide genetic correlations were computed against 1,409 external phenotypes comprising 1390 phenotypes and 19 curated psychiatric, cognitive, personality and sleep traits (**Supplementary Note 4.1**). Within-matrix Bonferroni thresholds were applied: intra-BD subphenotype *r*_G_ (**Supplementary Table 8; Fig. 1a**), *P* < 1.11 × 10⁻³; and BD-subphenotype × external-trait *r*_G_ (**Supplementary Table 9; Supplementary Fig. 1**), *P* < 2.63 × 10⁻⁴. A single Bonferroni-corrected significance threshold of *P <* 3.55 × 10^−5^ was also applied across all 1,409 phenotypes (**Fig. 2, Supplementary Table 10**). Some cognitive traits have been aligned so positive *r*_G_ reflects better cognitive performance^109^;(Matrix = Matrix Pattern Completion task; Memory = Memory – Pairs Matching Test; RT = Reaction Time; Symbol Digit = Symbol Digit Substitution Task; Trails-B = Trail Making Test – B; Tower = Tower Rearranging Task; VNR = Verbal Numerical Reasoning Test)^92^.

The major histocompatibility complex (MHC) was removed from SNP-heritability and genome-wide genetic-correlation analyses (chr6:28–34 Mb) and from transcriptome-wide colocalization (chr6:28–34 Mb), consistent with the previous BD GWAS^3^. The MHC was retained under several analyses following recent precedent, including SMR with HEIDI filtering^43^, MAGMA gene-based association^1,43^, and LAVA local genetic correlation^37^ (**Supplementary Note 1.8**).

### Genomic Structural Equation Modeling (Genomic SEM)

Genomic SEM (v0.0.5)^8^ was used to model the joint genetic architecture of the 10 bipolar phenotypes and, in a separate analysis, an expanded 20-trait cross-disorder framework incorporating 10 additional psychiatric and behavioral traits (ADHD, AN, ANX, ASD, BPD, MDD, PTSD, RISK, SCZ, and TS). Genomic SEM is unbiased in the presence of varying and unknown sample overlap across contributing GWAS samples, as the cross-trait intercepts estimated using multivariable LD score regression are used to estimate and account for sample overlap and phenotypic correlation^7^. Matrices were restricted to overlapping HapMap3^104^ SNPs across all included traits. Standard errors (s.e.) were corrected for residual population stratification by multiplying the standard errors by the square root of the univariate LDSC intercept when this exceeded 1, using the default genomic-control correction in Genomic SEM.

Alternative multivariate methods were considered and rejected as Genomic SEM uniquely controls for sample overlap, evaluates model fit, and tests heterogeneity (**Supplementary Note 3.1.1**).

### Robustness and sensitivity analyses for discovery

Three quality control steps supported the current analyses. (1) Univariate GWAS LDSC intercepts remained close to 1 across all 10 subphenotypes, consistent with polygenic signal rather than residual stratification^110^ (**Supplementary Table 2; Supplementary Fig. 5**). (2) For multivariate Genomic SEM, a chromosome split with parallel analysis confirmed the four-factor structure (**Supplementary Fig. 6**); the four-factor solution showed superior fit over alternatives, including a bifactor model (**Supplementary Note 3.2.1**). (3) MTAG maxFDR fell well below the 0.05 threshold across all 10 subphenotypes, supporting that MTAG-enhanced findings reflect each subphenotype rather than the larger BD GWAS (**Supplementary Table 2**); further supported by univariate versus MTAG subphenotype external *r*_G_ profiles which correlated highly (Pearson *r*=0.887– 0.987; **Supplementary Fig. 7**). See **Supplementary Note 11** for limitations and safeguards.

#### Factor model specification

Subphenotype-resolved discovery combined complementary analyses: univariate GWAS, multivariate Genomic SEM (common-factor and Q_SNP_ heterogeneity), and multi-trait MTAG. To guard against overfitting, EFA was performed on even autosomes and used to inform the confirmatory factor analysis fitted on odd autosomes, within the Genomic SEM framework. EFA was conducted with the ‘factanal’ function (maximum likelihood) for one through seven factors using both promax (oblique) and varimax (orthogonal) rotations. The promax solution was more stable and produced better-fitting confirmatory models and was therefore retained for model specification; consistent with the high genetic overlap among bipolar subphenotypes. Subphenotypes were assigned to a factor when their standardized EFA loading exceeded 0.3, with cross-loading subphenotypes assigned to their highest-loading factor. Factor retention was guided by eigenvalue magnitude, proportion of explained genetic variance, model fit indices. (CFI and SRMR), parallel analysis, and replicability across chromosome splits. All models specifying two through five factors fit better than the single common-factor model; models specifying six and seven factors failed to converge. The retained solutions were a four-factor model for the 10-trait analysis and a five-factor model for the 20-trait framework. Full factor-retention criteria, bifactor-model evaluation, and two-indicator factor justification are in **Supplementary Note 3.2**. CFA was subsequently specified and evaluated in Genomic SEM^8^; a good model fit was defined as CFI > 0.90 and SRMR < 0.08, with SRMR between 0.08 and 0.10 taken as acceptable fit, following conventional structural equation modeling standards and prior Genomic SEM applications^8^. Only traits with EFA loadings > 0.3 were included in CFA modeling. We tested the common liability models with and without factor loading constraints; with and without correlated residuals. In addition, we assessed if the traits were better modeled using a bifactor model and found this to be a less adequate fit than the two to five factor solutions. A correlated four-factor solution (compulsive, psychotic, dysregulated, and internalizing domains) was selected based on superior fit relative to two- to five-factor solutions; models specifying six and seven factors failed to converge. Factor labels were chosen to reflect the constituent subphenotypes of each factor, using HiTOP (Hierarchical Taxonomy of Psychopathology) spectrum names where applicable and the bipolar clinical literature otherwise (**Supplementary Note 3.1**). Full selection and retention criteria included parallel analysis, CFI/SRMR, eigenvalues, absence of Heywood cases, and chromosome-split replicability (**Supplementary Note 3.2)**; AIC, which favored the four-factor model (lowest AIC), agreed, and traits cross-loading > 0.30 were assigned to their highest-loading factor. A hierarchical second-order bipolar *p*-factor model was specified in which the four first-order domain factors loaded onto a second-order factor representing shared bipolar liability. The same methods used for the intra-bipolar model (10 subphenotypes) were repeated for the cross-disorder (10 subphenotypes and 10 external traits) model.

#### Common-factor GWAS

Multivariate GWAS of the hierarchical *p*-factor was conducted using the SNP-effects framework in Genomic SEM^6,8^, restricted to approximately 2.4 million HapMap3^104^ SNPs present across all 10 subphenotypes. The sumstats() function was used to align SNP effects across summary statistics and standardize relative to total variance. The effective sample size was used for the common-factor GWAS. After removing SNPs producing highly non-positive-definite matrices, 2,440,003 post-QC SNPs remained for the common liability analysis. A common-factor GWAS was conducted using the userGWAS() function in Genomic SEM^6,8^, estimating SNP effects on the single common bipolar factor. Genome-wide significance was defined as *P* < 5.0 × 10^−8^. As only a single common-factor GWAS was conducted, the standard genome-wide significance threshold was applied without further Bonferroni correction across factors. Loci were identified using the clumping functionality in PLINK v1.9^101^ with r^2^ <0.1 within −/+250 kb windows based on the 1000 Genomes Phase 3 European reference panel^97^.

#### Q_SNP_ Heterogeneity test

SNP-level heterogeneity statistics were computed to identify variants whose effects deviated significantly from the latent factor structure. Q_SNP_ is a χ²-distributed test statistic, with significant Q_SNP_ indicating heterogeneous SNP effects across the constituent subphenotypes. Significant Q_SNP_ (*P*<5×10^−8^) indicates SNP effects deviating from the common factor; the genome-wide threshold is applied without further Bonferroni correction^7,25^. Loci not showing significant Q_SNP_ heterogeneity (Q_SNP_ *P >* 5.0 × 10^−8^) were interpreted as acting through the common factor. Q_SNP_ statistics were computed using the same thresholds as above. After running the 10 BD subphenotypes common-factor GWAS, comparator GWAS from MDD and SCZ were added for comparison of their SNP-effects to the identified Q_SNPs_. See **Supplementary Tables 18**, and **Supplementary Table 18a** for Q_SNP_ GWAS-catalog annotations.

Three of the four latent factors were defined by two subphenotypes, with precedent in cross-disorder Genomic SEM^24^; their stability supported by their relatively strong internal BD *r*_G_ profiles and convergence in the *a priori* EFA, followed by CFA in the phenotypic data (**Supplementary Note 3.2–3.3**).

### Multi-trait analysis of GWAS

MTAG was applied to each BD subphenotype to increase N_EFF_ for phenotype-specific discovery while preserving trait fidelity. Each subphenotype summary statistics was jointly modeled with the largest available bipolar disorder GWAS meta-analysis (O’Connell et al.)^3^; European-ancestry, excluding self-report data; BD cases 59,287 and controls 781,022; N_EFF_=220,586. Sample and cohort overlap between each subphenotype and the O’Connell et al.^3^ BD framework was accounted for using LDSC-derived intercept correction^22^. Only phenotypes with non-zero h^2^_SNP_ and significant genetic correlation with framework traits were included. Only results with maxFDR < 0.05 were retained. To confirm that MTAG did not distort trait architecture, genome-wide genetic correlations between MTAG-enhanced and univariate summary statistics were estimated using.

LDSC; *r*_G_ profiles were highly concordant (Pearson *r*=0.887–0.987 across 10 phenotypes, *P <* 3.51 × 10⁻⁴; **Supplementary Fig. 7**). As an additional sensitivity check, 228 of the 356 unique lead SNPs (64.0%) were also genome-wide significant in at least one pre-MTAG discovery analysis (univariate, common-factor, or Q_SNP_), indicating that the majority of associations were already present before MTAG and that MTAG primarily amplified existing signals rather than generated new ones.

Independent loci for Genomic SEM and MTAG results were defined using LD clumping with r^2^ < 0.1 within −/+250 kb windows based on the 1000 Genomes Phase 3 European reference panel^97^, consistent with published multivariate GWAS applications of Genomic SEM^6,8,7,25^. Analyses included 4,338,104-5,807,084 post-QC SNPs per subphenotype (**Supplementary Table 2**). Within each of the four discovery analyses (univariate, Genomic SEM common-factor, Q_SNP_, MTAG), independent loci were defined by LD-clumping at r² < 0.1 (Genomic SEM and MTAG within ±250 kb, univariate within ±500 kb as using PGC convention)^21^. The union across the four discovery analyses was constructed by exact rsID deduplication of post-clumping lead SNPs, yielding 356 unique, independent GWS lead SNPs (**Supplementary Table 20**).

### Novel and replication of loci

#### Novelty

Following PGC convention within BD, novelty was assessed at the subphenotype level: a locus was considered novel if no prior genome-wide significant association (*P* < 5.0 × 10^−8^) for the same BD subphenotype had been reported in any GWAS indexed in the NHGRI-EBI GWAS Catalog^41^ at time of analysis (release e111, accessed 2026-04-02; **Supplementary Table 32-32a**). Each of the 10 subphenotypes was treated as an independent unit of discovery, consistent with the precedent^1,14^, reporting BD1 and BD2 hits as novel when the SNP was previously associated with generic BD. The first per-subphenotype genome-wide-significant associations reported here—for psychosis, unipolar mania, rapid cycling and schizoaffective disorder—arose in the univariate subphenotype GWAS (**Supplementary Table 7**), not from MTAG or common-factor analyses. Cross-disorder replication of all identified loci was additionally assessed against the GWAS Catalog (e111, ±500kb, *P* < 5.0 × 10^−8^); previous PGC BD GWAS^53,14,1^, were excluded as they share BD case cohorts with the present study and are reported separately. Per-discovery-method novelty rates and prior-PGC subphenotype comparison are in **Supplementary Notes 5.1–5.2**.

#### Replication

Lead SNPs were tested against the NHGRI-EBI GWAS Catalog (release e111, accessed 2026-04-02) and the most recent OCD^43^ GWAS, not yet indexed in the catalog. **Supplementary Table 32** contains two summary sub-tables. (1) Table 32.1 reports per-subphenotype counts of BD and cross-disorder replication for all 356 unique, independent BD-spectrum loci across the four discovery analyses (univariate, Genomic SEM common factor, Q_SNP_, MTAG), by exact lead-SNP lookup at GWS (*P* < 5.0 × 10^−8^). Common-factor lead SNPs were attributed cross-subphenotype (one per locus, replicated under each subphenotype trait pattern); Q_SNP_ lead SNPs were attributed to the driving subphenotype. (2) Table 32.2 reports subphenotype-specific lead-SNP replication for the univariate (**Supplementary Table 7**) and MTAG (**Supplementary Table 19**) lead SNPs—the two analyses with native one-to-one per-subphenotype attribution—against trait-matched or BD-spectrum entries at ±100/±250/±500 kb windows and exact-rsID, under GWS and suggestive (*P* < 10^−5^) thresholds. PGC BD studies sharing case cohorts^53,14,1^ were flagged. Replication detail of the full 356 unique, independent SNPs is provided in **Supplementary Table 20**, with cross-disorder detailed replications in **Supplementary Table 32a**.

### Local genetic correlation analysis

Local genetic correlations were estimated using LAVA v0.1.0, partitioning the genome into 2,495 approximately independent LD blocks, derived from 1000 Genomes Phase 3 European samples^97^. Thirty traits were tested which included (11 BD-spectrum traits (10 core BD subphenotypes plus suicide ideation [SI] as positive control), 19 external traits) (**Supplementary Note 4.1; Supplementary Tables 13-16**). For each LD block, local SNP-heritability was first estimated; bivariate local genetic correlations were computed only in blocks where both traits showed significant local signal (*P* < 2.00 × 10^−5^=0.05/2,495). Across all phenotype pairs and loci, 16,801 bivariate tests were performed, yielding a Bonferroni-corrected significance threshold of *P* < 2.98 × 10^−6^. Sample overlap was accounted for using LDSC^23^-derived bivariate intercepts. Local genetic correlation hotspots were defined as LD blocks demonstrating significant local genetic correlation across two or more bipolar phenotype pairs within the 10-trait model. Block-size sensitivity using 1,225 larger blocks yielded unstable local-*r*_G_ estimates; the developer-recommended 2,495-block partition was therefore retained (**Supplementary Note 4.1-4.4**).

### Functional annotation and gene mapping (FUMA)

Functional mapping and annotation were performed using FUMA v1.8.3^50^, implementing the SNP2GENE, GENE2FUNC, and Cell Type v1.8.3^111^ modules, applied consistently across all 10 MTAG-enhanced bipolar phenotype summary statistics. The 1000 Genomes Phase 3 European reference panel^97^ was used for LD estimation throughout. Default FUMA^50^ parameters were applied unless otherwise stated.

SNPs were mapped to genes using three complementary strategies. First, positional mapping assigned candidate SNPs to protein-coding genes using ANNOVAR (https://annovar.openbioinformatics.org/)^49^ annotations. Second, expression quantitative trait loci (eQTL) mapping linked SNPs to genes based on significant cis-eQTL associations (FDR < 0.05) across 13 (Genotype-Tissue Expression; GTEx, v8)^112^ brain tissues (amygdala, anterior cingulate cortex BA24, caudate basal ganglia, cerebellar hemisphere, cerebellum, cortex, frontal cortex BA9, hippocampus, hypothalamus, nucleus accumbens basal ganglia, putamen basal ganglia, spinal cord cervical c-1, and substantia nigra) and the PsychENCODE^113^ prefrontal cortex panel. Third, chromatin interaction mapping identified genes whose promoter regions (250 bp upstream to 500 bp downstream of transcription start sites) physically interacted with SNP-containing loci at a chromatin-interaction FDR ≤ 1 × 10^−6^ using Hi-C data from adult and fetal human cortex^114^ and 21 additional tissue/cell types^115^, alongside PsychENCODE^113^, Hi-C-derived enhancer-promoter interactions.

Gene-based association tests were conducted using MAGMA v1.10^44^ with a SNP-wise mean model, applied to 19,132 protein-coding genes using a window of 35 kilobases (kb) upstream and 10 kb downstream of gene boundaries, and the 1000 Genomes Phase 3 European reference panel^97^ for LD. SNPs with imputation INFO < 0.8, MAF < 0.01. The Bonferroni-corrected significance threshold for gene-based tests was *P* < 2.61×10^−6^. Gene-set association of the prioritized genes was tested using FUMA’s GENE2FUNC hypergeometric over-representation analysis against MSigDB v2023.1.Hs113 collections, WikiPathways, and the GWAS Catalog, with Benjamini– Hochberg FDR correction applied within each gene-set category (significant at adjusted *P* < 0.05). Tissue expression association was evaluated using MAGMA gene-property analysis across 54 GTEx(v8)^112^ tissue types, conditioning on average expression across all tissues (one-sided test); the Bonferroni-corrected threshold was *P <* 9.26 × 10⁻⁴. Gene expression heatmaps and tissue specificity association tests were generated using the GENE2FUNC module with GTEx(v8)^112^ expression data; tissue specificity was assessed against differentially expressed gene sets per tissue using hypergeometric tests with Bonferroni correction applied per differentially expressed gene (DEG) category (upregulated, downregulated, two-sided) separately. Variants were annotated for functional impact using Combined Annotation Dependent Depletion (CADD) v1.4^116,117^ and RegulomeDB v1.1^118^. Gene annotation used protein-coding gene definitions from Ensembl build 85 (GRCh37/hg19), as implemented in FUMA ANNOVAR-based positional annotation pipeline.

For developmental stage association analyses, Bonferroni correction was applied across 11 developmental stages (*P* < 4.55 × 10⁻³) and across 29 individual age timepoints (*P* < 1.72 × 10⁻³) separately.

Cell type association analysis was performed using the FUMA Cell Type^111^ module, applying MAGMA gene-property analysis across 31 single-cell RNA sequencing (scRNA-seq) datasets derived from eight underlying studies, human-brain datasets in FUMA. The datasets included: the Allen Brain Cell Atlas human middle temporal gyrus (MTG, levels 1 to 2) and lateral geniculate nucleus (LGN, levels 1 to 2)^119^; the DroNc-seq human hippocampus dataset^120^; the human prefrontal cortex developmental survey across gestational ages GW8–GW26 (GSE104276)^121^; the human cortex dataset with and without fetal cells (GSE67835)^122^; the Linnarsson lab human temporal cortex dataset (GSE101601)^123^; the GSE76381^124^ Linnarsson lab human ventral midbrain dataset—source of the eight midbrain subclasses (DA0, DA1, DA2, RN, Gaba, NbGaba, NbML5 and Sert): panel 8a shows all eight, and panel 8b the six FDR-significant subclasses (DA1, DA2, Gaba, NbGaba, NbML5 and Sert); PsychENCODE^113^ adult and developmental brain datasets; and the human prefrontal cortex developmental atlas spanning fetal through adult stages at three levels of cellular resolution (levels 1 to 3, across fetal, neonatal, infancy, childhood, adolescence, and adult timepoints; GSE168408)^125^.

For each dataset, gene expression specificity per cell type was defined as the average log₂- transformed normalized expression across cells of that type, conditioned on mean expression across all cell types within the dataset (one-sided test, beta >0). Multiple testing followed FUMA’s three-step protocol: per-dataset Bonferroni; within-dataset conditional; and cross-dataset conditional. Post-FUMA normalization was required for cross-trait and cross-factor comparison. FUMA’s MAGMA gene-property analysis outputs a standardized regression coefficient (BETA_STD) and its s.e. for each cell type × trait combination. BETA_STD reflects the regression of gene-based *z*-scores on cell type expression specificity scores; since gene-based *z*-scores scale with GWAS N_EFF_, BETA_STD is not directly comparable across traits with different N_EFF_. The 10 bipolar phenotypes span a range of MTAG N_EFF,_ meaning that raw BETA_STD comparisons across traits confound effect magnitude with statistical precision.

To enable cross-trait and cross-factor comparison, BETA_STD values were converted to *z*-scores by dividing by their standard errors (*z*=BETA_STD /s.e.) for all cell type × trait combinations reaching FDR < 0.05 in at least one trait. This *z*-score transformation preserves effect direction and relative magnitude while partially decoupling signal strength from N_EFF_, since the s.e. also scales with N_EFF_ such that the ratio is more stable across traits of differing power than BETA_STD alone. For each cell type, we computed (i) a factor-level mean *z*-score, the mean across traits assigned to each factor, and (ii) a cross-trait mean *z*-score (MeanZ_all), the mean across all traits reaching FDR < 0.05 (N_sig > 1) for that cell type. Factor *z*-score ratios were then computed as MeanZ_factor / MeanZ_all (**Supplementary Table 28c-e**). Ratios above 1.0 indicate association concentrated in that factor above the spectrum-wide baseline; ratios near 1.0 indicate pan-BD-spectrum contribution.

Two findings run counter to an N-inflation explanation. First, for midbrain Gaba and NbGaba neurons, there was FDR-significance across all 10 subphenotypes—BD2 showed a higher *z*-score than BD1 for Gaba despite its smaller effective sample size (**Supplementary Tables 28b and 28d-e**). Second, PSY showed a lower NormRatio than UM at midbrain despite its larger N_EFF_ (**Supplementary Tables 3 and 28d**). As N-confounding would inflate the higher-N trait in each pair, both reversals are inconsistent with N-inflation and support a biological-specificity interpretation.

To assess whether the cell-type signal generalized beyond the FUMA reference panels, MAGMA gene-property analysis, was applied outside FUMA to an adult human brain atlas,^56^ which resolves 461 transcriptomic clusters within 31 superclusters across the adult human central nervous system. Continuous cell-type specificity profiles served as gene-level covariates (one-sided test, β > 0), conditioned on MAGMA’s internal gene-level covariates (gene size, gene density, sample size, inverse minor-allele count, and their logarithms), with the 1000 Genomes Phase 3 European reference panel and genome build GRCh37 as in the primary analysis. Each of the 10 MTAG-enhanced subphenotypes was tested against all 461 clusters, with Benjamini-Hochberg FDR correction applied within each subphenotype across the 461 clusters. For each supercluster, the one-sided gene-property *z*-statistic was averaged across constituent clusters for cross-subphenotype comparison (**Supplementary Table 28f**).

### Transcriptome-Wide Association Studies (TWAS)

TWAS were conducted using FUSION (http://gusevlab.org/projects/fusion/)^45^ with gene expression prediction weights from 13 brain tissue panels derived from GTEx(v8)^112^: Cerebellum, Cerebellar Hemisphere, Pituitary, Cortex, Caudate Basal Ganglia, Nucleus Accumbens Basal Ganglia, Frontal Cortex BA9, Putamen Basal Ganglia, Anterior Cingulate Cortex BA24, Hypothalamus, Hippocampus, Amygdala, and Substantia Nigra; and the dorsolateral prefrontal cortex (DLPFC; CommonMind Consortium, CMC) downloaded at http://gusevlab.org/projects/fusion/; and a fetal brain panel^126^, using the 1000 Genomes Phase 3 European reference panel^97^ for LD. Across all 15 tissue panels, a total of 45,989 gene–tissue tests were conducted per trait. Gene-level *P*-values were aggregated across all 15 tissues using the Aggregated Cauchy Association Test (ACAT)^127^ with FDR < 0.05 applied to ACAT-aggregated *P*-values; this aggregated significance was used for initial gene discovery and as the basis for the prior credible gene framework. Bayesian colocalization was then performed on ACAT-significant genes using COLOC within FUSION (-- coloc P 0.05; *P* < 10⁻⁴), with strong support defined as a posterior probability for a shared causal variant (PP4) > 0.80. Colocalization superseded ACAT-aggregated significance as the scored transcriptomic paradigm, so that only genes with shared-causal-variant evidence entered the credible-gene framework. Within loci containing multiple significant TWAS signals, conditional analyses were performed using the FUSION^45^ post-processing framework (FUSION.post_process.R, --locus_win 100000) to distinguish independent transcriptomic associations.

Transcriptomic Structural Equation Modeling (T-SEM^109^) was conducted by running FUSION using MTAG-derived summary statistics for all 10 bipolar-spectrum subphenotypes as input, applying the same 15 brain tissue expression panels described above. This approach leverages cross-trait information to improve power for gene discovery. Gene-level *P*-values were aggregated across tissues using ACAT and FDR < 0.05 was applied for significance, consistent with the primary TWAS discovery pipeline; T-SEM results were used as a locus-level independence check rather than a separately scored paradigm in the final credible gene framework (**Supplementary Note 8**).

### Summary-based Mendelian Randomization (SMR)

SMR was performed using cis-eQTL instruments from BrainMeta v2^128^, a meta-analysis of brain cortex eQTL data, to leverage brain-specific expression data rather than whole-blood resources. LD was computed using the HRC EUR reference panel r1.0. Significance was defined as *P*_SMR_ < 3.11 × 10^−6^ (=0.05/16,062), the Bonferroni-corrected threshold across cis-eQTL probes tested in BrainMeta v2.

The HEIDI^48^ heterogeneity test was applied to all significant SMR results; associations were retained only when *P* SMR-HEIDI > 0.05, indicating a shared causal variant rather than linkage (Supplementary Tables 22-23). See **Supplementary Note 7** for additional details.

### Rare variant convergence

The 249 credible genes (Evidence Score ≥ 2) were cross-referenced against published gene-level burden results from BipEx^59^ (13,933 BD cases; 14,422 controls) and SCHEMA^58^ (24,248 SCZ cases; 97,322 controls), using each study’s published significance threshold: *P* < 5 × 10^−6^ for protein-truncating or damaging-missense variant burden in BipEx, and nominal *P* < 0.05 in SCHEMA. Association of credible genes among rare-variant hits was quantified using one-sided Fisher’s exact tests for excess overlap, evaluated against each study’s set of tested genes as the background, with overlap statistics reported in **Supplementary Table 33**. For genes overlapping rare-variant hits, we report the common-variant support from the present GWAS, extracting MAGMA gene-based association statistics to indicate the degree of corroboration across rare- and common-variant evidence; Bonferroni significance was defined as *P* < 2.61 × 10^−6^, consistent with the primary credible-gene framework (**Supplementary Note 9**).

### Drug target association

To assess whether credible genes from the multivariate bipolar spectrum analysis are associated with known or tractable drug targets, we applied one-sided hypergeometric tests to two predefined gene sets (**Supplementary Table 24-25**): (i) all credible genes with Evidence_Score≥2 (*N*=249; protein-coding genes mappable to OpenTargets [https://genetics.opentargets.org/]; accessed 2026-03-29); only 238 were mappable, and (ii) high-confidence genes with Evidence_Score ≥3 (*N*=89 mappable). The background was defined as all protein-coding genes testable by GWAS-based gene-scoring methods (MAGMA background; *N*=19,132).

Small-molecule (SM) tractability annotations were retrieved from the OpenTargets Platform targets parquet file (2024 release; downloaded 2026-03-29; https://genetics.opentargets.org/) using ChEMBL-derived (https://www.ebi.ac.uk/chembl/) SM modality tractability tiers 3. Three drug target categories were tested: (i) Approved Drug targets (Phase 4; background *n*=780), (ii) Clinical Targets (Phase 1 or above, including advanced clinical and approved; background *n*=1,110), and (iii) SM Tractable targets (any tractability tier from ‘Approved Drug’ through ‘Tractable Family’; background *n*=6,132; 32.1% of protein-coding genes). Six hypergeometric tests (2 gene sets × 3 drug categories) were run with Bonferroni *P* < 0.008. The test statistic was computed using the R function ‘phyper’. Full association statistics, annotated drug-target gene lists, and high-confidence gene tractability annotations are provided in **Supplementary Tables 24-25, 34-35**.

### Polygenic risk scoring

PRS were constructed from the 10 BD MTAG-enhanced phenotypes summary statistics. A leave-one-cohort-out framework was applied such that each target cohort’s discovery summary statistics excluded that cohort to prevent upward bias from sample overlap. Unlike the discovery MTAG (full-sample, with sample overlap to O’Connell et al. handled by the LDSC intercept), the polygenic-score MTAG was recomputed per fold: for each target cohort, both the subphenotype GWAS and the O’Connell et al. bipolar disorder meta-analysis used as the MTAG partner were the leave-that-cohort-out versions, therefore MTAG-enhanced discovery statistics for each fold contained no individuals from the projected cohort, ensuring fully out-of-sample evaluation. PRS were generated using PRS-CS^57^ with the ‘auto’ model to estimate the global shrinkage parameter, incorporating LD information from the UK Biobank European reference panel supplied with the PRS-CS software (**Supplementary Note 10**). Discovery statistics were restricted to autosomal SNPs with MAF > 0.01, INFO ≥ 0.8, and variants present in at least 75% of total N_EFF_. PRS-CS used per-locus median MTAG N_EFF_. Analyses included a median range of post-QC SNPs of 1,397,555-4,045,933 (**Supplementary Table 31**).

PRS were standardized to *z*-scores within each target cohort and evaluated by logistic regression in R (‘glm’ function; family=binomial, link=logit) with sex, the first five ancestry principal components, and genotyping batch as covariates (see **Supplementary Note 1.10**). Performance was quantified by liability-scale R^2^ (using a BD population prevalence of 2% combined with phenotype-specific prevalences)^3^, converted from the Nagelkerke’s pseudo-R2 (computed using the. ‘fmsb’ R package, https://cran.r-project.org/web/packages/fmsb/index.html) to the liability scale^105^. Area under the receiver operating characteristic curve (AUC), was computed using the ‘pROC’ R package (https://cran.r-project.org/web/packages/pROC/index.html), and odds ratios for individuals in the top decile versus the middle quintile of the PRS distribution. Absolute risk was additionally calculated for individuals in the top one and 10 percent PRS strata as the predicted probability from the logistic-regression model, calibrated to each subphenotype’s population prevalence (**Supplementary Table 31**). Bootstrap confidence intervals were computed for R^2^ estimates using 10,000 nonparametric bootstrap replicates demonstrating no significant difference between sex-stratified scores across the 10 BD traits. Cohort-specific PRS estimates were performed using both fixed-effects and random-effects meta-analyses; only random-effects weighted results are reported, providing conservative inference under the between-cohort heterogeneity observed across cohorts.

### Data availability

Genome-wide association summary statistics for all subphenotypes analyses presented here will be made publicly available through the PGC download page (https://www.med.unc.edu/pgc/download-results/) upon publication of this article. Genotype data are available for a subset of contributing cohorts, including dbGaP accession numbers and/or access restrictions, as described in the cohort descriptions of the **Supplementary Information**. The analyses used the most recent PGC bipolar disorder GWAS (2025); these summary statistics are available at the PGC download page (https://www.med.unc.edu/pgc/download-results/). LD reference panels are publicly available: the 1000 Genomes Project Phase 3 European reference panel (https://www.internationalgenome.org/), the HapMap3 SNP set (distributed with LDSC; https://github.com/bulik/ldsc), and the Haplotype Reference Consortium r1.0 panel, available through the European Genome-phenome Archive (https://ega-archive.org/). The UK Biobank European LD reference used for PRS-CS is distributed with the PRS-CS software (https://github.com/getian107/PRScs). Gene-expression, eQTL and chromatin-interaction resources used for functional mapping, TWAS and SMR are publicly available: GTEx v8 (https://gtexportal.org/), the PsychENCODE resource (http://resource.psychencode.org/), and the dorsolateral prefrontal cortex (CommonMind Consortium) and fetal-brain expression weights distributed with FUSION (http://gusevlab.org/projects/fusion/). Adult- and fetal-cortex Hi-C interaction maps were obtained from the sources cited in the Methods. Brain-cortex cis-eQTL instruments for SMR were obtained from BrainMeta v2 (https://yanglab.westlake.edu.cn/software/smr/). Single-cell and single-nucleus transcriptomic reference datasets used for cell-type association are available through FUMA (https://fuma.ctglab.nl/) and from their original sources, including the Allen Brain Cell Atlas, the DroNc-seq human hippocampus dataset, the PsychENCODE adult and developmental datasets, and datasets deposited in the Gene Expression Omnibus under accessions GSE104276, GSE67835, GSE101601, GSE76381 and GSE168408. The adult human brain atlas of Siletti et al. is publicly available (DOI: 10.1126/science.add7046). Gene-set, variant-annotation and drug-target resources are publicly available: the NHGRI-EBI GWAS Catalog (release e111, accessed April 2 2026; https://www.ebi.ac.uk/gwas/), the Molecular Signatures Database (MSigDB v2023.1.Hs; https://www.gsea-msigdb.org/gsea/msigdb/), the Open Targets Platform (2024 release, accessed March 29 2026; https://genetics.opentargets.org/) and ChEMBL (https://www.ebi.ac.uk/chembl/). Gene-level rare-variant burden results were obtained from the Bipolar Exome consortium (BipEx; https://bipex.broadinstitute.org/) and the Schizophrenia Exome Meta-Analysis (SCHEMA; https://schema.broadinstitute.org/).

### Code availability

No custom code was developed for this study. All software and tools used for the analyses presented are publicly available and referenced within the respective sections of the **Methods** and **Supplementary Information**. Quality control and imputation used RICOPILI (https://sites.google.com/a/broadinstitute.org/ricopili/), incorporating PLINK v1.90 (https://www.cog-genomics.org/plink2/), EIGENSOFT/EIGENSTRAT v6.1.4 (https://www.hsph.harvard.edu/alkes-price/software/), Eagle v2.3.5 (https://alkesgroup.broadinstitute.org/Eagle/), Minimac3 (https://genome.sph.umich.edu/wiki/Minimac3) and METAL (https://genome.sph.umich.edu/wiki/METAL_Documentation); allelic-inconsistency filtering used DENTIST (https://github.com/Yves-CHEN/DENTIST). SNP-heritability, genetic correlations and summary-statistic harmonization used LDSC v1.0.1 (https://github.com/bulik/ldsc). Modeling used Genomic SEM v0.0.5 (https://github.com/GenomicSEM/GenomicSEM; documentation at https://github.com/GenomicSEM/GenomicSEM/wiki). Multi-trait analysis used MTAG (https://github.com/JonJala/.mtag) and local genetic correlations used LAVA (https://github.com/josefin-werme/LAVA). Functional mapping and gene-based, gene-set, tissue, developmental and cell-type association used FUMA v1.8.3 (https://fuma.ctglab.nl/) and MAGMA v1.10 (https://ctg.cncr.nl/software/magma). Transcriptome-wide association and colocalization used FUSION with COLOC (http://gusevlab.org/projects/fusion/); summary-based Mendelian randomization and the HEIDI test used SMR (https://yanglab.westlake.edu.cn/software/smr/). Polygenic scores used PRS-CS (https://github.com/getian107/PRScs). Variant annotation used ANNOVAR (https://annovar.openbioinformatics.org/), CADD v1.4 (https://cadd.gs.washington.edu/) and RegulomeDB v1.1 (https://regulomedb.org/). Liability-scale and predictive metrics were computed with the R packages ‘fmsb’ (https://cran.r-project.org/web/packages/fmsb/index.html) and ‘pROC’ (https://cran.r-project.org/web/packages/pROC/index.html).

## Acknowledgments

Permission was obtained for statistical analyses to be carried out on the Genetic Cluster Computer (http://www.geneticcluster.org) hosted by SURFsara (The Netherlands).

## Author contributions

The management group, comprising a subset of authors, was responsible for the study design, conduct, primary and final interpretation, and included M.A., R.A.O., K.S.O., N.M., A.J.F., M.G- S., H.J.E., F.J.M., O.A.A., A.D.F., and A.M. The writing group was led by A.D.F. and A.M. and was responsible for primary drafting and editing of the manuscript. M.T. contributed clinical assessments and analyses of the clinical data. T.V. was responsible for the main analyses presented in the paper. Numerous authors beyond the initial writing group contributed to data interpretation and provided edits, comments and suggestions to the paper. All authors reviewed the manuscript critically for important intellectual content and approved the final version of the manuscript for publication. The Chair of the PGC is P.F.S. The Bipolar Disorder Working Group of the PGC is led by O.A.A.

## Additional information

Supplementary information. Supplementary information accompanies this paper, see **SupplementaryInformation.docx** and **SupplementaryTables.xlsx.**

**Correspondence and requests for materials**. Correspondence and requests for materials should be addressed to Tracey van der Veen.

