## Supplementary Information for "Genomic dimensions deconstruct the clinical heterogeneity of bipolar disorder"

**Supplementary Note 1 - Diagnostic Criteria, Harmonization, and GWAS Quality Control .. 3**

### Supplementary Note 1 - Diagnostic Criteria, Harmonization, and GWAS Quality Control

##### 1.1 Terminology and Scope

Our use of "subphenotype" follows established usage in psychiatric genetics, referring to clinically characterizable features ascertained within a case cohort of a primary diagnostic category and analyzed to reduce genetic heterogeneity. Three classes of clinical heterogeneity within BD are covered: (1) diagnostic subtypes (BD1, BD2, schizoaffective disorder bipolar type); (2) clinical specifiers reflecting illness course (unipolar mania, rapid cycling, psychosis within BD); and (3) co-occurring psychiatric conditions reaching diagnostic threshold in BD cases (panic disorder, OCD, substance use disorder, and suicide attempt). These subphenotypes are observable, trait-like clinical features suitable for heritability estimation and multivariate structural modeling.

Throughout, "subphenotypes" refer to the bipolar-relevant clinical units that are the primary analytic objects of the intra-bipolar Genomic SEM; "external disorders" or "external traits" refer to psychiatric diagnoses and behavioral traits outside BD (including MDD, SCZ, anxiety disorders, PTSD, and risk-taking behavior). Following PGC Cross-Disorder Group convention, we refer to the joint analysis of BD subphenotypes and external disorders as the "cross-disorder framework" rather than "transdiagnostic", reserving "transdiagnostic" for its established Genomic SEM meaning: the highest-order shared genetic component spanning multiple cross-disorder factors.

##### 1.2 Ten-Trait Model, Clinical Axes, and Sample Composition

Ten of 16 candidate BD subphenotypes met inclusion for joint Genomic SEM and MTAG modeling (**Supplementary Note 1.4**). The 10 subphenotypes span three clinical axes: diagnostic subtypes (BD1, BD2, SZA), course specifiers (UM, PSY, RC), and comorbidities (PAN, OCD, SUD, SA). Cohort inclusion required item-level ascertainment of (1) lifetime hallucinations/delusions; (2) ≥ 4 distinct mood episodes within 12 months (RC); (3) absence of lifetime major depressive episode (UM); (4) lifetime suicide attempt with at least some intent to result in death (SA); and (4) comorbid PAN, OCD, and SUD reaching diagnostic threshold under international consensus criteria (DSM-IV, ICD, or equivalent). The present design builds on the prior work of the Bipolar Disorder and Schizophrenia Working Group[1,2], here expanding the BD case sample from approximately 13,000 to 38,022 across 57 cohorts; differences from prior work are detailed in **Supplementary Note 5.1.2**. Subphenotypes were ascertained at each study site using diagnostic instruments, case records, and participant interviews; per-site ascertainment details are given in **Supplementary Note 12**. Phenotype selection was based on prior evidence of heritability and further refined by data availability across study sites and consistency of definitions. See **Supplementary Note 1.3 below** for harmonized diagnostic definitions and inclusion criteria, and **Supplementary Table 5** for population prevalence (*K*) parameters used as input for liability-scale transformations.

##### 1.3 Diagnostic Definitions and Operationalization

BD subphenotypes were collected by each study site using a combination of diagnostic instruments, case records and participant interviews. The selection of phenotypes for collection by this group was determined by literature searches to determine phenotypes with prior evidence for heritability. It was further refined dependent on the availability of phenotype data across a range of study sites and the consistency by which the phenotypes were defined. Across the 57 contributing cohorts, lifetime BD and BD subphenotype diagnoses were established through a combination of structured psychiatric interviews (most commonly SCID, SADS-L/SADS, SCAN, DIGS, MINI, OPCRIT, or AMDP), clinician-administered checklists, medical record review, and the family history method, applied according to international consensus criteria (DSM-IV, DSM-IV-TR, DSM-5, DSM-III, ICD-10, ICD-9, RDC, or combinations thereof depending on cohort era and country). Cohorts variably employed consensus best-estimate procedures with two or more independent diagnosticians, expert clinical review, or for population-based biobank cohorts, register-based ICD coding from inpatient discharge records. Per-cohort diagnostic systems, instruments, procedures, and case ascertainment are detailed in **Supplementary Note 1-2**, and **12**. Phenotype data were collected retrospectively; however, longitudinal studies show low BD1–BD2 conversion, supporting subtype stability. Rapid cycling and the depression episodes in unipolar mania were both indexed as lifetime rather than current, partially offsetting cross-sectional underestimation.

Harmonized subphenotype definitions for the 10 BD subphenotypes used in univariate, multivariate Genomic SEM, and MTAG discovery are provided below.

| **Subphenotype** | **Harmonized Definition** |
|---|---|
| Bipolar I (BD1) | Bipolar I disorder, defined by the occurrence of at least one manic episode (Ruderfer et al., 2014[2], 2018[1]; Stahl et al., 2019[3]). |
| Bipolar II (BD2) | Bipolar II disorder, defined by the occurrence of at least one hypomanic episode and at least one major depressive episode, with no manic episode (Ruderfer et al., 2014, 2018; Stahl et al., 2019[3]). |
| Schizoaffective, Bipolar Disorder Type (SZA) | Schizoaffective disorder, bipolar type, defined by concurrent psychotic and manic features meeting the criteria for both schizophrenia and a manic episode (Ruderfer et al., 2014, 2018; Stahl et al., 2019[3]). |
| Psychosis (PSY) | Presence of any of the following: delusions, hallucinations, positive formal thought disorder, catatonia or grossly disorganized behavior (Ruderfer et al., 2014, 2018). |
| Rapid Cycling (RC) | Occurrence of four or more distinct mood episodes within a 12-month period at any point during bipolar illness (Ruderfer et al., 2014, 2018; Miola et al., 2023). |
| Suicide Attempt (SA) | Suicide attempt, defined as a deliberate act of self-harm with at least some intent to result in death; excludes suicidal ideation without action and non-suicidal self-injury (Mullins et al., 2019[4]; Ruderfer et al., 2014, 2018). |
| Unipolar Mania (UM) | Occurrence of one or more manic episodes with no major depressive episode (Ruderfer et al., 2018). |
| Panic Disorder (PAN) | Comorbid panic disorder, defined as recurrent unexpected panic attacks and anticipatory anxiety (Ruderfer et al., 2014, 2018). |
| Obsessive Compulsive Disorder (OCD) | Comorbid obsessive-compulsive disorder, defined as recurrent intrusive obsessions and ritualistic compulsions (Ruderfer et al., 2014, 2018). |
| Substance Use Disorder (SUD) | Comorbid alcohol use disorder and/or substance use disorder; cohort-level definitions encompass both abuse and dependence (Ruderfer et al., 2014, 2018). |

##### 1.4 Candidate Subphenotypes Evaluated and Excluded

Six subphenotype GWAS were excluded from multivariate analyses: (1) Suicide ideation (SI) was evaluated for inclusion; however, it showed extremely high genetic correlation with SA ($r_G$ >

0.99), resulting in multicollinearity and unstable parameter estimation in Genomic SEM. SA was retained as the more clinically severe subphenotype. Clinical-only versions of (2) BD1 (BD1 Clinical) and (3) BD2 (BD2 Clinical) were also evaluated; the combined clinical-and-community-ascertained traits (BD1 Clin/Com, BD2 Clin/Com) were retained as the primary subtype traits with the larger effective sample sizes and broader ascertainment for multivariate modeling. Three age of onset subphenotypes were excluded for negligible SNP-heritability ($h^2_{SNP} < 0.05$ on the observed scale); (4) age of onset depression (AOD), (5) age of onset mania (AOM), (6) age of onset of BD (AOO), along with numerous clinical phenotypes for which we lacked sufficient data for inclusion in our genetic analyses.

Variables external to the patient — adverse life events and socio-economic status (SES) — were not considered for inclusion in the 10-trait model, as they index environmental exposures rather than within-case genetic heterogeneity. Sex showed minimal main-effect association with bipolar disorder trait risk in our sex-stratified $r_G$s, even after down-sampling; however, sex was retained as a covariate in PRS for sex-stratified risk prediction (see **Supplementary Note 1.10**).

**1.5 Subphenotype Inclusion Criteria**

Subphenotype inclusion required: (1) diagnostic validity (subclinical cases were excluded); (2) reliability across diverse international settings; (3) intrinsic biology (SES and geography excluded to avoid confounding with ascertainment bias); and (4) statistical power ($h^2_{SNP}$ $z$-score > 4; **Supplementary Table 2**). All 10 retained subphenotypes met this criterion. Inverse-variance weighted meta-analysis used also ensured low-N cohorts contribute information proportional to precision.

Subclinical cases were excluded rather than retained as a lower-severity stratum since sub-threshold presentations fail two of the four inclusion criteria simultaneously: they lack diagnostic validity under any of the harmonized diagnostic systems applied across the 57 cohorts, and their reliable ascertainment cannot be assumed across clinical settings with differing thresholds for case identification. Unlike full-threshold cases, sub-threshold presentations are not routinely documented or clinically confirmed in the structured assessments used for subphenotype derivation; their inclusion could introduce systematic misclassification that is not correctable by downstream quality control.

##### 1.6 Harmonization, Missingness, and Denominator Definition

Since subphenotypes are nested, individuals with a BD diagnosis, absent of the subphenotype, were removed, reducing confounding by overall bipolar liability. Cases without explicit assessment for a given subphenotype were excluded rather than assumed status, to minimize misclassification bias. Prevalence percentages were calculated as the proportion of explicitly assessed cases.

##### 1.7 Genotyping, Quality Control, and Imputation

A centralized QC/imputation pipeline was performed using RICOPILI (Rapid Imputation for COnsortias PIpeLIne[5]) (version 2024_Nov_21.001). Genotyping, QC, imputation, and association analysis procedures are described in the Methods section. Cohort-specific details are summarized in **Supplementary Table 1**, for full detailed cohort descriptions, see **Supplementary Note 12**.

##### 1.8 MHC Region Handling

The extended major histocompatibility complex (MHC; chr6:28–34 Mb, GRCh37) combines high gene density with long-range LD; whether it was excluded, depended on the method. The MHC was filtered out in SNP-heritability and global (LDSC) genetic-correlation estimation, and transcriptome-wide association with colocalization, applied similarly to recent literature[6,7]; the SMR–HEIDI procedure included the MHC and reported MHC genes among HEIDI-supported SMR signals. MAGMA gene-based association applies competitive correction for gene size, gene density, and between-gene linkage disequilibrium; MHC genes were correspondingly retained in the MAGMA gene-based results. Tissue and cell-type association with MAGMA gene-property regression, retained the MHC. LAVA local genetic correlation spanned the MHC following the method's developers[8], who retained this as a recognized site of cross-trait pleiotropy.

##### 1.9 Independent Lead SNP Identification and Novelty Assessment

For univariate GWAS (**Supplementary Table 7**), independent lead SNPs were identified using LD-based clumping in PLINK ($r^2 < 0.1$ within 500 kb), using a European ancestry LD reference panel, with lead SNPs required to have INFO ≥ 0.8 and $P < 5 \times 10^{-8}$. For multivariate analyses (MTAG[9], Genomic SEM[10]), independent loci were defined using FUMA[11] ($r^2 < 0.1$, 250 kb merge window).

##### 1.10 Covariates, Ancestry Control, and Sensitivity Checks

SES and geography were not incorporated as covariates, as these likely reflect external modifiers rather than intrinsic biology and risk confounding biological heterogeneity, with e.g., healthcare access disparities. Sex was excluded from primary GWAS pipelines, consistent with prior large-scale analyses, after we reconfirmed near-unity sex-stratified $r_G$ for BD autosomal architecture ($r_G > 0.95$; Mullins et al., 2021)[12] in our sex-stratified sensitivity analyses for BD1, BD2, and SZA estimated near-unity $r_G > 0.95$ between male-only and female-only summary statistics; down sampled sex-ratio-equated analyses (female and male excess respectively), also yielded $r_G > 0.95$. These sex-stratified analyses were performed in the internal cohorts for which full individual-level data were available. Based on genotype derived (PLINK .fam) sex, the sample composition was: Bipolar I cases (58.8% female); Bipolar II cases (63.2% female); schizoaffective disorder, bipolar type, cases (51.7% female); and the shared control set (52.3% female). Individuals of undetermined genotype sex were excluded from these percentages. The corresponding male-only and female-only GWAS for each subtype comprised these sex-specific case counts together with the sex-matched control set.

Additional subphenotypes lacked an adequate $N_{EFF}$ for further sex-stratification. Biological sex was included as a covariate in the final PRS regression models applied to independent target samples; the main effect of sex was not statistically significant across any of the 10 subphenotype models (all $P > 0.05$), indicating that within the target sample, sex did not significantly explain additional variation in bipolar subphenotypes, after accounting for PRS and population structure.

### Supplementary Note 2 - LDSC Estimation, Trait Stability, and Sample Overlap

##### 2.1 Summary Statistics Harmonization

Summary statistics were harmonized using munge function, as described in the Methods.

##### 2.2 Heritability Estimation and Trait Stability Filtering

Three ‘Age of onset’ subphenotypes (AOO, AOM, AOD) were excluded due to $h^2_{SNP} < 0.05$ to avoid destabilizing the covariance estimates (**Supplementary Table 2**). Subphenotypes with lower BD population prevalences were retained despite smaller effective sample sizes as their $h^2_{SNP}$ $z$-statistics were > 4.

**Liability-scale conversion**. Observed-scale SNP-heritability’s were converted to the liability scale using a population prevalence ($K$) for each subphenotype, with the sample prevalence (case proportion; **Supplementary Table 6**). Two rules set $K$: diagnostic categories (BD1, BD2, schizoaffective; unipolar mania modeled as BD1) used the lifetime population prevalence of the diagnosis; course specifiers and comorbidities used the joint phenotype, the base bipolar prevalence (2%) multiplied by the within-bipolar prevalence of the feature, since cases are bipolar patients carrying the feature contrasted against general-population controls. Per-subphenotype values, derivations, and sources are in **Supplementary Table 5**.

##### 2.3 LDSC Intercept and Attenuation Ratios

LDSC intercepts for the 10 subphenotypes remained close to 1, <1.05 consistent with polygenic signal rather than residual stratification[13]. Attenuation ratios remained low to moderate, ranging from 0.029 to 0.204. Since the attenuation ratio is defined as (Intercept-1)/(Mean chi square-1)[13], values inflate in smaller samples where mean chi square approaches 1; elevated ratios, confined to the smallest datasets, likely represent denominator contraction[13] rather than uncontrolled confounding, as intercepts remained well-controlled. A comparable pattern is seen in a related design[14], which reported attenuation ratios up to 0.42, with the largest values confined to the two smallest datasets, whereas their well-powered latent factors remained low. To correct for residual stratification in s.e. estimation, s.e. of SNP effects were multiplied by the LDSC univariate intercept (when > 1) prior to Genomic SEM[10] covariance matrix construction.

**Nested subphenotype design, sample overlap and $r_G$ interpretation**. The 10 bipolar subphenotypes shared samples, creating potential sample overlap. This was explicitly controlled for using the LDSC bivariate cross-trait intercepts in Genomic SEM[10]. Genomic SEM ingests a sample-overlap-corrected genetic covariance matrix using bivariate LDSC intercepts, so overlapping cases between subphenotypes do not bias factor estimates. Beyond sample overlap correction, three features of the design rule against artifactual inflation of intra-BD genetic correlations. (1) for nested subphenotypes, BD cases not meeting the subphenotype criterion were removed rather than retained as controls, meaning the case-control contrast indexes subphenotype liability against a healthy baseline rather than against the complementary BD subgroup - the standard case-healthy-control design used in the previous[1] analyses. Every subphenotype is BD-affected individuals contrasted against the same healthy controls, so all contrasts share a general-BD-liability axis. That is not statistical bias; it is part of what the factors represent. (2) LDSC univariate intercepts remained well-controlled across all 10 subphenotypes, indicating low residual stratification beyond what polygenic signal accounts for, (3) and most directly, $Q_{SNP}$ heterogeneity testing recovers 12 GWS loci whose cross-subphenotype effects depart from the single common-factor pattern, diverging in magnitude and, at several loci, in direction — incompatible with a single confounded liability axis. At these loci BD1 and PSY depart concordantly but with differing magnitude, consistent with genetic proximity between psychosis-within-BD and BD1 liability rather than a design-induced correlation.

#### 2.4 Phenome-wide Genetic Correlation Analyses

Pairwise $r_G$ were estimated using bivariate LDSC. Differences between $r_G$ estimates were tested using block-jackknife and interpreted relative to sampling precision ($r_G$/s.e.). Cross-trait phenome-wide $r_G$ were computed against 1,390 external traits using CTG-VL (https://vl.genoma.io) and a curated set of 19 psychiatric and cognitive disorders (1,409 total external GWAS); Bonferroni-corrected threshold $P < 3.55 \times 10^{-5}$ (**Supplementary Table 10**). Intra-BD subphenotype $r_G$ (**Supplementary Table 8**) used a Bonferroni threshold of $P < 1.11 \times 10^{-3}$; and BD-subphenotype × external-trait $r_G$ (**Supplementary Table 9; Supplementary Fig. 1**) used $P < 2.63 \times 10^{-4}$.

##### 2.5 MTAG Q-Q Plot Comparison

Comparative quantile-quantile (Q-Q) plots display the distribution of association statistics for the baseline subphenotype univariate GWAS meta-analyses (black) against the re-weighted MTAG, (which includes BD GWAS SNP effects; O'Connell et al. 2025)[6] model (blue) in **Supplementary Fig.5**. Univariate intercepts remained close to 1, consistent with polygenicity rather than stratification. The significant leftward deflection of the blue curves illustrates the increased discovery power of leveraging the bipolar disorder framework, while preserving valid null distributions.

##### 2.6 Heritability and Genetic Correlation Parameter Estimates

Intra-bipolar genetic correlations ranged from low to high; the weakest pairing was between BD2-UM. Pairwise intra-bipolar genetic correlations are provided in **Supplementary Table 8**. Local genetic correlations (LAVA) were largely directionally consistent with global LDSC patterns, with locus-specific pleiotropic hotspots linking psychosis-tied (BD1, PSY) and dysregulated (RC, SA) subphenotypes, the two factors loading strongest to the general *P*g (bipolar disorder spectrum) in the four-factor model (**Fig.3**) (**Supplementary Tables 13, 15-16**).

#### Supplementary Note 3 - Genomic SEM Model Specification, Fit Indices, and Multivariate GWAS

**Multi-dimensional modeling framework**. We hypothesized that BD harbors a structured multi-dimensional genomic architecture that is only recoverable at sufficient phenotypic resolution, and that identifying this architecture may reveal genomic substrates of clinically meaningful heterogeneity. The dual-framework design allows us to ask not only which genomic dimensions exist within BD; however, which are BD-intrinsic versus shared with the broader psychiatric landscape. No prior study has resolved this simultaneously at the scale and phenotypic breadth of the present analysis.

##### 3.1 Model Specification and Input Construction

Genomic SEM was used to model the joint genetic architecture of the 10 bipolar subphenotypes and, in a separate analysis, an expanded 20-trait cross-disorder framework incorporating (1) schizophrenia, (2) ADHD, (3) ASD, (4) anorexia nervosa, (5) Tourette's syndrome, (6) MDD, (7) PTSD, (8) anxiety disorders, (9) borderline personality disorder, and (10) risk-taking behavior.

**Factor Labeling**. Factor names in the factor models emerged using existing nomenclature, with reference to the HiTOP framework[15] where HiTOP spectra provide a clean mapping, and to the BD-specific clinical-psychopathology literature where they do not. The convention follows the precedent established in prior multivariate genomic analyses of psychiatric cross-disorder architecture[16,17], in which existing nomenclature labels are used when indicator sets do not correspond to a single HiTOP spectrum:

**$F1_{BD}$-Compulsive** (OCD, SUD). HiTOP places OCD within the Internalizing spectrum (Fear subfactor) and SUD within the Disinhibited Externalizing spectrum (Substance Abuse subfactor), such that no single HiTOP spectrum unifies these indicators[15]. The label follows the convention used by Grotzinger et al. (2022)[16], who identified a Compulsive factor encompassing OCD alongside other compulsive-spectrum conditions in Genomic SEM analyses[17].

**$F2_{BD}$-Psychotic** (BD1, PSY, SZA, UM). All four indicators share positive psychotic features as a defining or prominent clinical characteristic. HiTOP assigns these subphenotypes to the Thought Disorder spectrum[15,18]. The label is tied to the psychotic symptom dimension that unites these traits, and parallels the Psychotic factor identified in prior Genomic SEM studies across psychiatric disorders[16].

**$F3_{BD}$-Dysregulated** (RC, SA). This factor captures a BD-specific indicator pairing for which HiTOP provides no unifying spectrum: rapid cycling is not represented in the HiTOP model, and suicidality is placed within the Internalizing-Distress subfactor[15]. The label follows the bipolar clinical-psychopathology literature, which identifies affective instability and emotion dysregulation as the latent dimension uniting these phenomena. Koenigsberg (2010)[19] defines rapid-cycling mood changes as one of seven constituent phenomena of affective instability. MacKinnon and Pies (2006)[20] argue that rapid mood switching and affective instability share a common underlying mechanism, and cite earlier family-study evidence that rapid mood switching co-aggregates with suicide attempts in bipolar pedigrees[21]. Affective lability has been identified as a core euthymic-phase dimension of bipolar disorder[22].

**F4$_{BD}$-Internalizing** (PAN, BD2). Panic disorder is a prototypical Internalizing-Fear indicator in HiTOP[15]. Hypomania, which defines bipolar II disorder (BD2), has been shown to load on the Internalizing spectrum in hierarchical structural psychopathology analyses[23], consistent with the predominantly affective rather than psychotic character of its clinical presentation. The label reflects this shared Internalizing liability.

**Hierarchical consistency with broader taxonomies**. When the 10-trait BD model is extended to the 20-trait common-disorder model, F3$_{BD}$-Dysregulated and F4$_{BD}$-Internalizing merge into a single F4$_{CD}$-Dysregulated/Internalizing factor (**Supplementary Note 3.4**). This convergence mirrors the HiTOP framework, in which suicidality, emotional lability, and anxiety components are all placed within the broader Internalizing spectrum[15]. The BD-specific separation of F3$_{BD}$-Dysregulated from F4$_{BD}$-Internalizing, therefore, likely captures a finer level of BD specific granularity.

##### 3.1.1 Alternative multivariate frameworks considered

Three alternatives to multivariate Genomic SEM modeling were considered and rejected[24–26]. Genomic SEM uniquely combined case-control GWAS summary statistic input, bivariate LDSC overlap correction, pre-specified confirmatory factor testing with formal fit indices (CFI, SRMR, $\chi^2$), and $Q_{SNP}$ heterogeneity testing. Grotzinger et al. (2019)[10] established it as the canonical multivariate framework for psychiatric-disorder consortium GWAS; the present application extends it from cross-disorder to within-bipolar disorder subphenotype architecture.

#### 3.2 Exploratory and Confirmatory Factor Analysis

EFA was performed using the ‘factanal’ R package within the Genomic SEM[10] framework, using a chromosome-split cross-validation approach: factor extraction on even-numbered autosomes, and CFA on odd-numbered autosomes. Factor extraction used diagonally weighted least squares (DWLS) estimation with oblique (promax) rotation to permit inter-factor correlation, consistent with expectations of partially overlapping liability domains. Models specifying common-factor, bifactor and two- to five-factor solutions were evaluated; six and seven factor models failed to converge in both 10-trait and 20-trait analyses. Factor retention criteria included parallel-analysis simulation, eigenvalues > 1.0, proportion of explained genetic variance, interpretability of loading structure, model fit indices (CFI, SRMR), absence of Heywood cases (negative residual variances), and replicability across chromosome splits.

##### 3.2.1 Bifactor model evaluation

A bifactor model was estimated to evaluate whether high inter-factor correlations reflect factor indistinction rather than genuine, shared liability, with a general bipolar *p*-factor and four orthogonal domain-specific factors (s_Compulsive, s_Psychotic, s_Dysregulated, s_Internalizing) simultaneously predicted all 10 subphenotypes. Model fit was evaluated using the same criteria as all other specifications reported previously[16]. The bifactor model produced worse approximate fit (CFI = 0.945, SRMR = 0.114) than the correlated four-factor model, the bifactor SRMR exceeding the 0.10 acceptability threshold (**Supplementary Table 12**).

**Two-indicator factor justification**. The $F1_{BD}$-Compulsive (OCD, SUD), $F3_{BD}$-Dysregulated (RC, SA) and $F4_{BD}$-Internalizing (PAN, BD2) factors each comprise two primary indicators. Two-indicator factors are minimally identified in standard SEM; estimable within a correlated factor model when the remaining factors provide additional constraints on the solution — a condition satisfied here given the four-factor structure and the high inter-factor correlations. Two-indicator factors are not without precedent in Genomic SEM applications[17]. Stability across leave-one-trait-out specifications supports the empirical identifiability of these factors in the present 10-trait model, demonstrating the structure was not overly reliant on subphenotypes with the larger sample sizes.

To establish that no single subphenotype drove the factor solution, each of the 10 subphenotypes was removed in turn and the four-factor model re-estimated (10 leave-one-trait-out solutions). Across all 10 iterations the standardized loadings and inter-factor correlations were stable, and all iterations retained CFI > 0.95 and SRMR < 0.10. Decisively, removing the most highly powered subphenotypes, left the loading pattern and inter-factor correlations unchanged. The four-factor structure was therefore not an artifact of the best-powered traits.

CFA was specified and evaluated in Genomic SEM; acceptable fit was defined as close to CFI > 0.95 and SRMR < 0.10, following conventional structural equation modeling fit standards and consistent with prior Genomic SEM applications[10,16,17], see **Supplementary Note 3.4**. Nested models were compared using changes in fit indices. The correlated four-factor solution was selected based on superior fit (CFI = 0.976, SRMR = 0.071) relative to the common-factor (CFI = 0.953, SRMR = 0.089) and two- to five-factor alternatives. Inter-factor correlations were high (0.778–0.892) but less than unity. As these factors partition a single disorder rather than distinct disorders, they share more of a common bipolar liability and are expected to correlate more strongly than cross-disorder factors in current research[17]; separability rests on the bifactor rejection, distinct loadings, and SNP-level heterogeneity, not on low correlation. Full model fit indices are provided in **Supplementary Tables 12**.

##### 3.3 Phenotypic Structure Validation

To evaluate whether the clinical subphenotype covariance structure supported the same dimensional organization as the genetic model, subphenotype-only factor analyses were performed using individual-level data in PGC participants with complete subphenotype assessment (BD cases = 17,908; healthy controls = 34,314). Phenotypic indicators were z-standardized prior to analysis. EFA and CFA (CFI = 0.946; SRMR = 0.084, < 0.10 acceptable fit)[10] on phenotypic covariance matrices also supported a four-factor structure consistent with the genetic model. Subphenotype-only factor analysis was treated as supportive validation rather than a primary determinant of model specification.

##### 3.4 Cross-disorder Model Fit Comparison

The bipolar-restricted 10-trait model explained 82.8% of shared genetic variance, whereas embedding within the 20-trait model (which best fit a five factor model), explained 69.1%, reflecting modest attenuation under cross-disorder expansion; and similar to ~66.0% variance explained by the five factor model in Grotzinger et al., 2026[17]. Although higher, the SRMR of 0.083 here for the 20-trait model falls within the acceptable range (< 0.10)[10]. Hierarchical loading patterns shifted under 20-trait embedding: psychotic and dysregulated domains projected stronger onto the bipolar *p*-factor in the 10-trait model, whereas embedding redistributed projection toward the internalizing domain.

##### 3.5 Structured $Q_{SNP}$ Heterogeneity: Detailed Per-Axis Results

The $Q_{SNP}$ heterogeneity test was estimated within Genomic SEM across the 10 BD subphenotypes only; the external comparators SCZ and MDD were not inputs to the $Q_{SNP}$ model and contribute to no $Q_{SNP}$ $\chi^2$ or *P*-value. Following the test, the 12 $Q_{SNP}$-significant loci were characterized by per-trait *z*-score profiling against the 10 univariate BD subphenotypes, with SCZ and MDD SNP effects added solely for post-hoc interpretation. A cross-trait *z*-score heatmap for all 12 $Q_{SNP}$ loci is provided in **Supplementary Fig.2**, with the BD subphenotypes grouped by factor domain and SCZ and MDD shown separately as external comparators.

**BD1/UM Discordant Sign-effects**. Sign flips occurred across subphenotypes, representing discordance between BD1-UM: *ADCY2* is an established bipolar I disorder GWAS locus, differentiating between bipolar disorder and major depression[27]; *ADD3* and *MAD1L1*.

**UM and SZA versus BD1 residual variance on $F2_{BD}$-Psychotic**. UM and SZA showed higher residual variance on $F2_{BD}$-Psychotic than BD1, which continued into the embedded model with SCZ. As UM and SZA are within the lowest-powered subphenotypes despite relatively high $h2_{SNP}$, this divergence is interpreted relative to sampling precision and awaits replication in better-powered samples.

### Supplementary Note 4 - Local Genetic Correlation (LAVA) Parameterization and Results

##### 4.1 LAVA Results Summary

Local genetic correlations were estimated in the 10 BD subphenotypes (and BD SI for a positive control), analyzed against 19 external psychiatric, cognitive, personality, and sleep traits: (1) SCZ[28], (2) MDD[29], (3) anorexia nervosa (AN)[30], (4) Tourette's syndrome (TS)[31], (5) autism spectrum disorder (ASD)[32], (6) attention deficit/hyperactivity disorder (ADHD)[33], (7) Borderline Personality Disorder (BPD)[34], (8) ANX[35], (9) post-traumatic stress disorder (PTSD)[36], (10) risk-taking behavior (RISK)[37], (11) Insomnia[38], (12) Intelligence[39], (13) Reaction Time[40], (14) Symbol

Digit[40], (15) T-MTB (Trail-Making B)[40], (16) Tower task[40], and (17) Verbal Numerical Reasoning[40] , (18) Matrix reasoning[40], and (19) Memory[40].

##### 4.2 Intra-Bipolar Hotspots

The most shared hotspot was at 21q22.13, with complete directional concordance across most subphenotypes. This region contains *KCNJ6*, a brain-expressed G-protein–activated potassium channel involved in dopaminergic modulation and affective regulation. A second hotspot at 3p22.2 encompassed *TRANK1/SCN5A/SLC22A13/14*, demonstrating dense multi-phenotype pleiotropy spanning predominantly psychosis-factor subphenotypes. BD1 and PSY shared the most significant loci (**Supplementary Table 13**) - the strongest concordant intra-bipolar overlap - whereas BD1 and BD2 shared no Bonferroni significant regions despite a relatively high global $r_G$.

##### 4.3 BD1–BD2 Local Genetic Architecture

Despite high genome-wide $r_G$, BD1 and BD2 shared no Bonferroni-significant local genetic correlations. Loci surviving correction (FDR-Benjamini–Hochberg [BH] $q < 0.05$), all showed positive local correlation; most regions were however shared rather than BD1-BD2 specific.

**Factor-concordant dimensional channeling**. BD1 and BD2 local $r_G$ may selectively overlap genetically with factor-concordant external partners, rather than converging mostly on shared loci. BD1 shared FDR-significant local loci with MDD; however, zero with ANX, despite MDD and ANX sharing global $r_G > 0.877$ (**Supplementary Table 9**). BD2 showed no FDR-significant local overlap with PSY, however it shared loci with both MDD and ANX, consistent with its internalizing factor loading. BD1's concentrated psychotic-spectrum local contrasts with its absence of ANX overlap; BD2 overlaps with MDD and ANX, however not at PSY regions — mirroring the BD1–BD2 dislocation identified by Genomic SEM (**Supplementary Tables 13-16)**.

**Interpretation**. The most parsimonious reading is that BD1 and BD2 represent differential reweighting of a largely shared dimensional liability toward psychosis-spectrum and internalizing poles. Their FDR-significant BD1–BD2 local correlations were predominantly pleiotropic rather than subtype-specific, and none survived Bonferroni correction. Internalizing traits formed a greater share of BD2's overlap than BD1's. As these are within-subtype proportions, they are not likely driven by BD2's lower testing density, consistent with their psychotic versus internalizing factor placement. We note that this local null is at least partly power-limited and so results here do not alone establish this diffuse architecture.

##### 4.4 Local Genetic Correlation Blocks

**LAVA block-size sensitivity**. The bivariate analysis was repeated for six BD-relevant trait pairs (BD1–BD2, BD1–PSY, BD1–SZA, BD1–UM, BD2–PSY, BD2–SZA) using 1,225 larger LD-independent blocks (target 5,000 SNPs per block). With larger blocks, bivariate local $r_G$ estimates were largely undeterminable across all trait pairs, including well-powered within-factor pairs. The developer-recommended 2,495-block partition was therefore retained for the primary analysis.

##### 4.5 Rapid Cycling: Cross-domain liability

Clinically, rapid cycling is linked to high depressive morbidity, serious suicide attempts, and comorbidity with anxiety and substance use disorders, in both familial and population-based samples. Here, RC course indexes aggregated liability across partially separable dimensions rather than domain-specific, and that pharmacological complexity in this group may reflect distributed genetic burden, rather than a single targetable pathway for polarity instability.

#### Supplementary Note 5 - MTAG Implementation and maxFDR Filtering

##### 5.1 Discovery Summary Across Analysis Frameworks

**Novelty by discovery method**. Of the 356 unique GWS loci, 228 were genome-wide significant in at least one pre-MTAG analysis (univariate GWAS, 60; Genomic SEM common-factor, 162; $Q_{SNP}$, 12; deduplicated), and 208 were not recovered by MTAG. Of these 208 MTAG-independent loci, 137 (65.9%) fall outside any GWS locus reported within ±500 kb in the prior BD GWAS[3,6,12,41], representing discovery attributable to the multivariate framework rather than power borrowed from earlier studies. MTAG validity was indexed by MaxFDR, which fell far below 0.05 across the 10 analyses (**Supplementary Table 2)**, which confirmed that the additional SNP effects likely reflect power gain rather than spurious signal[9]; this was supported by a high Pearson $r$ in a comparison of univariate to MTAG genetic correlations (**Supplementary Fig.7**).

##### 5.1.1 Univariate GWAS: Per-Subphenotype Lead Loci

GWS univariate associations ($P < 5 \times 10^{-8}$; **Supplementary Table 7**) emerged in 10 of 16 BD subphenotype GWAS meta-analyses (60 GWS hits; 31 novel). Several subphenotypes yielded no GWS univariate hits, reflecting lower heritability's; signal was recovered through MTAG (Supplementary Table 19). BD2 GWAS replicated the *SLIT3* lead variant from an earlier study[12] and identified a novel locus upstream of *PCDH7*. One of the strongest univariate signal across subphenotypes was at *TRANK1*, reaching GWS in two psychotic-factor subphenotypes (BD1, PSY); *TRANK1* (here a high confidence credible gene) and *SCN2A* were independently fine-mapped as high-confidence BD risk genes[42]. The novel SZA locus here was sub-threshold in an earlier BD GWAS for SZA[3].

The single GWS RC locus at *TMEM130-TRRAP-NPTX2* is novel and the first for RC; *TRRAP* encodes a core component of histone acetyltransferase complexes involved in chromatin remodeling, and rare coding variants in *TRRAP* have been implicated in neurodevelopmental disorders with behavioral dysregulation[43]; *NPTX2* implicates parvalbumin (PV) interneurons and is a candidate biomarker for BD[44].

The single GWS UM locus harboring the gene *IMMP2L* is also novel and the first UM association, its SNP is located within the copy-number variant (CNV) region linked to the *IMMP2L; IMMP2L* encodes an inner mitochondrial-membrane peptidase subunit. Structural variants spanning *IMMP2L* have prior associations with schizophrenia[45], consistent with UM's loading on the $F2_{BD}$-Psychotic factor; *IMMP2L* was also previously implicated in an earlier BD GWAS[3].

The single GWS SA locus *PRKN*(*PARK2)* was here novel for SA. *PRKN(PARK2)* has established GWS associations across multiple psychiatric traits: post-traumatic stress disorder[46,47]; depression[48]; neuroticism[49]; smoking initiation[50]; and insomnia[38].

##### 5.1.2 Comparison with Prior PGC BD Subphenotype Discovery

The present 10-subphenotype discovery framework builds on the efforts of Ruderfer et al., 2018, which established significant heritable signal for psychotic features in BD and suicide attempt within a BD–SCZ joint framework spanning ~13,000 BD cases. The present design provides an approximately threefold larger BD case sample, BD-restricted analytic framework, multivariate Genomic SEM decomposition supplementing univariate GWAS, identifying 356 unique, independent GWS loci across 10 subphenotypes (and 158 novel), including the first univariate GWS loci for psychosis-within-BD, unipolar mania, rapid cycling, and schizoaffective disorder.

##### 5.1.3 Functional Annotation of MTAG Lead SNPs (CADD)

FUMA CADD v1.4 deleteriousness scores for the per-subphenotype MTAG lead SNPs are reported in **Supplementary Table 19**. The strongest deleteriousness signals were largely shared rather than trait-specific: the most deleterious lead SNP genome-wide, an exonic variant at *BTN2A1*, reached genome-wide significance in all 10 subphenotypes, as did other relatively high CADD genes, *CTD-2026C7.1*, *FURIN* and *THSD7A*—consistent with these loci indexing common-spectrum liability. Against this shared background, one high-CADD exonic lead SNP was unique to a BD1, *SCN2A*, corroborated by independent fine-mapping of *SCN2A* as a high-confidence BD risk gene[42].

#### 5.2 Replication of Genome-wide Significant Loci

As the subphenotype cases derive from the same cohorts as prior BD GWAS, and MTAG incorporated the PGC BD meta-analysis (O'Connell et al., 2025), recovery of established BD loci is expected by construction and is reported only as an internal consistency check, not independent replication (**Supplementary Table 32**; cohort-sharing studies flagged). Independent replication was therefore assessed against GWAS of other psychiatric disorders, which also do not share the MTAG input: across the 10 subphenotypes, 71.9–89.5% of lead SNPs matched a trait-matched or BD-spectrum independent psychiatric GWAS within ±500 kb (**Supplementary Table 32**). Per-subphenotype replication rates were: nine of 10 subphenotypes replicated with GWS loci within ±500 kb and PAN at $P$=7×10$^{-7}$. PAN overlapped two panic disorder GWAS Catalog loci; see **Supplementary Table 32a**. Critically, 208 of the 356 unique loci — 137 of them novel — were not recovered by MTAG, and so are independent of the BD GWAS that MTAG leverages. At the gene level, of the 36 high-confidence credible genes reported by O'Connell et al.[6] (Evidence Count ≥ 3), 16 were recovered here, by non-MTAG methods.

### Supplementary Note 6 - Gene Mapping, Functional Annotation, and Credible Gene Prioritization

##### 6.1 Cell Type and Tissue Association

This study uses the term 'association'[51] to describe the MAGMA test of cell-type expression specificity against subphenotypes.

###### 6.1.1 Psychosis-factor gradient in midbrain neurons

Across the $F2_{BD}$-Psychotic gradient (**Supplementary Table 28d**), midbrain dopaminergic and GABAergic cell types differentiate the four psychotic-factor subphenotypes, ranking SZA → PSY → UM → BD1 at most subclasses. DA1 reaches FDR significance in nine subphenotypes, not SZA; DA2 in six, and Sert in four (BD1, BD2, PAN, SUD; SZA was non-significant) (**Supplementary Table 28a**). These orderings run counter to sample size: raw association $Z$ increases with effective sample size (**Supplementary Table 28b**), yet smaller-sample BD2 exceeds BD1, and smaller-sample UM exceeds PSY, at specific midbrain cell types—these reversals inconsistent with sample-size inflation (**Supplementary Table 28d**). This direction replicated in the independent adult atlas. Across the 26 clusters with midbrain as the predominant region (**Supplementary Table 28f**), UM exceeded PSY (24 of 26 clusters; paired Wilcoxon $P < 0.001$) at roughly one-quarter of its effective sample size, and BD and BD2 were comparable ($P = 0.12$) despite BD1's 3.6-fold larger effective sample. Both BD1 and UM exceeded the schizophrenia-spectrum subphenotypes (BD1 versus PSY, $P < 0.001$; versus SZA, $P = 0.014$), as in the developmental panel.

###### 6.1.2 Factor-level differentiation

The midbrain signal partitions by factor (**Supplementary Table 28c**). Among the non-psychotic factors, only $F4_{BD}$-Internalizing carries relatively elevated midbrain ratios—highest at GABAergic cells, driven by its BD2 indicator.

##### 6.1.3 Contextualization within the broader cell-type framework

These associations recover and extend the PGC4 BD GWAS[6]. Where that study implicated cortical and hippocampal interneurons, pyramidal neurons, and striatal medium spiny neurons in the same adult human brain atlas[52], the present analysis recovers these superclusters across all 10 subphenotypes—126 of 461 the atlas[52] clusters are significant in every subphenotype (**Supplementary Table 28f**)—and adds a within-subphenotype standardization across effective sample sizes (**Supplementary Table 28e**) and a suggestive midbrain dopaminergic and GABAergic gradient along the $F2_{BD}$-Psychotic axis (**Supplementary Table 28d**). The adult atlas contains a single, sparsely represented midbrain dopaminergic cluster (Splat_395) versus the developmental panel's DA0/DA1/DA2 resolution — so it cannot resolve the dopaminergic peak. Relative to the largest OCD GWAS[7], which localized OCD heritability to cortical and hippocampal excitatory neurons and D1/D2 striatal medium spiny neurons, the present OCD subphenotype recovered those populations and additionally implicated midbrain dopaminergic (DA1, DA2) and GABAergic (Gaba, NbGaba) source neurons with VIP interneurons (**Supplementary Tables 28a, 28b**), resolving the dopamine signal to dopamine-producing rather than receptor-bearing cells. Likewise, where the PGC3 schizophrenia GWAS[28] identified cortical excitatory neurons, D1/D2 medium spiny neurons, and cortical and hippocampal interneurons as leading associations, the psychotic-factor subphenotypes recovered these and add a graded midbrain dopaminergic and GABAergic association. Reference-atlas differences—developing human midbrain and adult human cortex here versus adult mouse nervous system there—limit one-to-one correspondence.

##### 6.1.4 Independent replication in the adult human brain atlas

Across the 64 FUMA cell-type tests, 24 reached pan-BD-spectrum FDR significance and 40 were differential, 16 of them specific to a single subphenotype (**Supplementary Tables 28a, 28b**); sample-size-standardized scores (**Supplementary Table 28e**) enabled between-subphenotype comparison. VIP interneurons reached FDR significance in seven of 10 subphenotypes (**Supplementary Table 28a-b**).

To test generalization beyond the FUMA reference panels, MAGMA analysis was applied to an external adult human brain atlas[52] (461 clusters in 31 superclusters); 126 of 461 clusters reached significance in all 10 subphenotypes, the strongest shared signal localizing to telencephalic superclusters—upper- and deep-layer intratelencephalic neurons, CGE and MGE interneurons, and medium spiny neurons—independent of the FUMA panels (**Fig. 8c; Supplementary Fig. 8; Supplementary Table 28f**). Glial replication was selective: only astrocyte clusters reached FDR significance, peaking in PSY, with associations also in OCD, BD2, SA, BD1, SZA and RC. Trait-specific signals concentrated in BD1, including the only trait-specific glial signal in the developmental panel (prenatal GW26 OPCs; **Supplementary Tables 28a, 28b**), deep-layer L5.6 cortical neurons (L5.6_TLE4_SCUBE1) (**Supplementary Tables 28a, 28b**) and intratelencephalic neurons (DLIT_142, DLIT_143) (**Supplementary Table 28f**), midbrain DA0, red nucleus (RN), and hippocampal exCA1 and exPFC2 (**Supplementary Tables 28a, 28b**). UM carried 11, predominantly Splatter and deep-layer near-projecting (DLNP_89) cell types (**Supplementary Table 28f**). PAN's two strongest singleton associations were two PAN only upper-layer cortical excitatory (Exc_L2.4_LINC00507_GLP2R,Exc_L2.3_LINC00507_ FREM3), and SUD aligned with early prenatal GW10_GABAergic_neurons (**Supplementary Tables 28a, 28b**).

##### 6.2 Credible gene prioritization framework

Genes were scored across six paradigms, each contributing one point to an Evidence Score (0–6): TWAS with colocalization; SMR with HEIDI filtering; MAGMA gene-based testing; rare-variant burden in BipEx or SCHEMA; ANNOVAR functional annotation; and FUMA positional mapping. OpenTargets L2G scores were annotated but not scored. Evidence Score ≥ 2 defined 249 credible genes and ≥ 3 defined 89 high confidence (HC), and ≥ 4 defined robust genes (**Supplementary Table 21**). SMR analyses also flagged subphenotype-specific divergence from schizophrenia: across the 10 subphenotypes, including SZA-associated genes which carried SMR effects directionally opposite to SCZ, supporting SZA inclusion in our BD analyses, differentiated from SCZ (**Supplementary Table 22**).

##### 6.3 SP4 transcription factor

*SP4* carried the highest convergent support in the study (Evidence Score = 5; MAGMA-significant in all 10 subphenotypes, with rare-variant burden in SCHEMA at nominal $P < 0.05$) and was identified at the common-factor level before MTAG (**Supplementary Table 21**). It indexes shared, pan-spectrum bipolar liability rather than a dimension-specific signal — tied to the common factor and confirmed by rare-variant evidence, without trait-level effects supporting domain-specific inference; this replicates its most recent finding as a top BD GWAS loci[6].

##### 6.4 Serine/threonine kinase 4 (STK4)

*STK4* encodes a serine/threonine kinase central to Hippo signaling and is a novel common-factor GWS hit, and significant across all 10 subphenotypes (Evidence Score = 5; **Supplementary Table 21**). Across all 10, its SMR effects are discordant with ADHD and Borderline Personality Disorder (BPD) and concordant with SCZ and risk tolerance (**Supplementary Table 22**). This direction tracks the established bipolar genetic-correlation sign structure and is not tested against a directional null; it is reported as convergent support rather than gene-specific directional evidence. *STK4* carries an OpenTargets Structure-with-Ligand tractability annotation but no approved or clinical-stage drug for any indication (**Supplementary Table 25**).

##### 6.5 Drug-target association

Of the 249 credible genes, 238 mapped to OpenTargets and all 89 HC genes mapped. One-sided hypergeometric tests across three small-molecule tractability categories for two gene sets (six tests; Bonferroni $P < 8.33\times10^{-3}$) were significant for both SM-tractability and approved-drug overlap (**Supplementary Tables 24, 34–3**5). Ten credible genes are approved-drug targets—*CACNA1C, CUL4A, HDAC5, KCNS1, ACHE, CACNA1B, HTR6, NDUFS2, SCN2A* and *THRA*—though none targets a primary bipolar phenotype (**Supplementary Table 25**).

##### 6.6 Biologically annotated subset

The 249 credible genes are listed in full in **Supplementary Table 21**. The Evidence-Score-2 genes warrant more caution in interpretation than the HC and robust tiers, resting on exactly two complementary lines of evidence.

##### 6.7 Calcium and potassium channel genes

*CACNA1B* (Evidence Score = 2) associated here encodes the α1B subunit of the N-type (Cav2.2) voltage-gated calcium channel, mediating presynaptic calcium influx for neurotransmitter release.

*CACNA1C* (12p13.33; Evidence Score = 3) encodes the L-type (Cav1.2) α1C subunit. *KCNN3* (1q21.3; Evidence Score = 3) encodes a calcium-activated potassium channel (**Supplementary Table 21**). Together they implicate calcium-mediated excitability as a broad shared mechanism across BD liability, extending pharmacological interest from the L-type to the N-type channel at the subphenotype-level.

### Supplementary Note 7 - SMR, HEIDI, Colocalization, and Transcriptomic Directionality

##### 7.1 Transcriptomic directionality (SMR effect-direction matrix)

Per-pair concordant and discordant SMR gene counts across the 10 BD subphenotypes versus six external traits (ADHD, ANX, BPD, MDD, RISK, SCZ) are in **Supplementary Table 22**. From these, **Supplementary Table 23** summarizes each subphenotype × external-trait cell: concordant and discordant gene counts, percent concordance, and the most subphenotype-specific concordant (↑) and discordant (↓) gene. Subphenotype-specific signal was defined as |z_BDx − mean (z across the other nine subphenotypes)| for the same gene–external pair, where z = β/s.e.

### Supplementary Note 8 - TWAS and Conditional TWAS Analyses

TWAS was performed in FUSION using expression-prediction weights from 13 GTEx v8 brain tissues, dorsolateral prefrontal cortex, and a fetal brain panel (15 panels). Cross-tissue signals were aggregated with the Aggregated Cauchy Association Test (ACAT) at FDR < 0.05. Conditional TWAS within loci carrying multiple significant signals used the FUSION post-processing framework.

### Supplementary Note 9 - Rare Variant Convergence

##### 9.1 Rationale and Dimension-Specific Convergence

Rare-variant burden was incorporated as an independent evidence line in the tiered credible-gene framework. Five credible genes overlapped a rare-variant burden hit (**Supplementary Table 33**):

*SP4* (SCHEMA, $P = 0.040$) and *GIGYF1, PACS1* and *ZNF584* (BipEx, $P = 4.67 \times 10^{-6}$) recurred across all 10 subphenotypes, with BD1 additionally implicating *FAM83H*.

### Supplementary Note 10 - Polygenic Risk Score Construction and Validation

##### 10.1 Interpretation

PRS-based absolute-risk estimates derive from clinical samples ascertained in psychiatric settings with high case density. They are not directly applicable to population-level prediction and require prospective validation in multi-ancestry and community cohorts in longitudinal designs (see **Methods** and **Supplementary Fig.4**).

### Supplementary Note 11 — Limitations and Interpretive Safeguards

##### 11.1 Cross-cohort phenotyping and missingness

Limitation. Cohorts varied in the clinical detail they recorded, so some subphenotypes were assessed in only a subset of cohorts. Despite standardized case definitions, this uneven coverage may have reduced power and attenuated some subphenotype-specific associations (**Supplementary Note 1.6**).

Safeguard. A nested case-only design was used: BD cases not meeting a subphenotype criterion were removed rather than retained as controls, and cases lacking explicit assessment were excluded rather than assumed negative, reducing confounding by overall bipolar liability.

##### 11.2 Meta-analysis heterogeneity and ascertainment differences

Limitation. PRS effect estimates varied substantially across cohorts ($I^2 = 81–96\%$; Supplementary Table 31). Consistent with Trubetskoy et al. (2022)[28], this heterogeneity likely reflects differences in case ascertainment (e.g., hospital versus community samples) rather than an unstable genetic effect.

Safeguard. All PRS estimates used random-effects meta-analysis, acknowledging the high between-cohort heterogeneity (**Supplementary Table 31**).

##### 11.3 Inflation, confounding, and LD-reference dependence

Limitation. LDSC estimates ($h^2$_SNP, $r_G$) are sensitive to LD-reference matching, residual stratification, subphenotype definition, and the weaker signal of narrowly defined traits; an external LD reference was used rather than in-sample LD.

Safeguard. LDSC univariate intercepts were well-controlled across all 10 subphenotypes (< 1.05; **Supplementary Table 2**), indicating minimal residual stratification, and the MHC region was excluded throughout. For MTAG, restriction to SNPs shared with the larger BD GWAS, a low maxFDR, and high pre-versus-post-MTAG effect concordance (Pearson $r$ = 0.887–0.987); indicate the subphenotype signal was largely preserved.

##### 11.4 Genomic SEM: covariance stability and model specification

Limitation. Genomic SEM results depend on the stability of LDSC-derived heritability/covariance estimates and correct model specification. Latent factors are statistical summaries of shared genetic covariance and should not be interpreted as causal entities.

Safeguard. Sample overlap across subphenotype GWAS was corrected by the bivariate LDSC intercepts incorporated in Genomic SEM, as applied in prior cross-trait psychiatric analyses, and the selected four-factor model showed good fit (**Supplementary Table 12**).

##### 11.5 LAVA: power imbalance and locus definition

Limitation. Local $r_G$ estimates are sensitive to locus definition, local LD structure, and unequal trait power; absence of local signal may reflect limited power rather than true absence of shared architecture.

Safeguard. LAVA's two-stage design uses the univariate local-heritability test as a filter ahead of the bivariate test: stable, interpretable local correlations require sufficient local genetic signal[8], so local genetic correlations were estimated only at loci where both traits showed significant local heritability, screening out underpowered loci before the bivariate step. Werme et al. further show this univariate filtering does not bias the resulting correlations. Additionally, the developer-recommended 2,495-block partition was retained for optimal performance: a block-size sensitivity analysis using larger blocks left local genetic correlation estimates largely undeterminable, including for well-powered within-factor pairs.

**11.6 Transcriptomic integration (TWAS, colocalization, SMR)**

Limitation. TWAS and SMR do not establish causal mediation; associations can reflect LD tagging, horizontal pleiotropy, or correlation between predicted expression and causal variants. Results are constrained by the available expression quantitative trait loci (eQTL) reference panel tissue, developmental stage, sample size, and ancestry.

Safeguard. SMR signals were filtered by HEIDI, and TWAS-prioritized genes required colocalization (**Supplementary Table 21**). A six-paradigm framework was used to grade the credibility of prioritized genes.

**11.7 Credible gene prioritization: avoiding single method overinterpretation**

Limitation. Any single mapping paradigm can generate false positives from LD, model assumptions, or reference-panel constraints.

Safeguard. Convergence across the six-paradigm framework was required—at least two paradigms for credible genes (Evidence Score $\geq 2$) and at least three for high-confidence genes ($\geq 3$)—to mitigate single-method false positives (**Supplementary Table 21**).

**11.8 Ancestry**

Limitation. Analyses were largely restricted to European-ancestry samples, limiting generalizability across populations. Multi-ancestry deep-phenotyping is an ongoing priority.

**11.9 Causal inference and mechanistic mediation**

Limitation. Multivariate genomic modeling identifies latent liability dimensions and their shared genetic architecture but does not establish causal directionality or mechanistic mediation between genomic factors and clinical outcomes. Credible gene–subphenotype associations reflect statistical colocalization rather than demonstrated causal pathways.

Safeguard. Findings are framed as hypothesis-driving, requiring future functional and experimental validation.

##### 11.10 Polygenic risk score generalizability

Limitation. PRS performance estimates derive from clinical samples ascertained in psychiatric settings with high case density. Absolute-risk figures are not directly applicable to population-level prediction and may overstate discriminative performance in unselected cohorts.

##### 11.11 Cross-trait assortative mating and LDSC assumptions

Limitation. Cross-trait assortative mating may inflate genetic correlation estimates, although such effects are unlikely to account for the magnitude of correlations observed across psychiatric dimensions.

### Supplementary Note 12 — Detailed Cohort Sample Descriptions

##### Sample descriptions

We included data from 57 cohorts (**Supplementary Table 1**), totaling 38,022 cases and 188,010 controls of predominantly European descent. For 52 cohorts, raw genotype and phenotype data were shared with the Psychiatric Genomics Consortium (PGC). Cases were required to meet international consensus criteria for a lifetime diagnosis of bipolar disorder (BD) established using structured diagnostic instruments from assessments by trained interviewers, clinician-administered checklists, or medical record review. For the remaining five external cohorts, GWAS summary statistics for BD were shared with the PGC. Most samples included here were also in previous PGC BD GWAS papers[3,12,41].

Below we describe the ascertainment and diagnosis of the participants in each individual cohort comprising this report. Most cohorts have been published individually, and the primary report can usually be found using the PubMed identifiers provided. The lead PI of each sample warranted that their protocol was approved by their local Ethical Committee and that all participants provided written informed consent. **Supplementary Table 1** provides additional detail, including sample sizes and genotyping array. As the lifetime prevalence of BD is around 0.4-2%, some cohorts use controls that are not screened for BD[53,54]. Given this low prevalence, unscreened controls introduce minimal case contamination, biasing subphenotype associations conservatively toward the null. The boldfaced first line for each sample indicates study PI, PubMed ID if published, country (study name), and the PGC internal tag or study identifier.

**======== PGC1 Samples ========**

**Rietschel, M; Nöthen, MM, Cichon, S | 21926972 [PGC1] | BOMA-Germany I | bip_bonn_eur**

Cases for the BOMA-Bipolar Study were ascertained from consecutive admissions to the inpatient units of the Department of Psychiatry and Psychotherapy at the University of Bonn and at the Central Institute for Mental Health in Mannheim, University of Heidelberg, Germany. DSM-IV lifetime diagnoses of bipolar I disorder were assigned using a consensus best-estimate procedure, based on all available information, including a structured interview with the SCID and SADS-L, medical records, and the family history method. In addition, the OPCRIT[55] checklist was used for the detailed polydiagnostic documentation of symptoms. Controls were ascertained from three population-based studies in Germany (PopGen, KORA, and Heinz-Nixdorf-Recall Study). The control subjects were not screened for mental illness. Study protocols were reviewed and approved in advance by Institutional Review Boards of the participating institutions. All subjects provided written informed consent.

**Corvin, A | 18711365 [PGC1] | Ireland | bip_dub1_eur**

Samples were collected as part of a larger study of the genetics of psychotic disorders in the Republic of Ireland, under protocols approved by the relevant IRBs and with written informed consent that permitted repository use. Cases were recruited from Hospitals and Community psychiatric facilities in Ireland by a psychiatrist or psychiatric nurse trained to use the SCID. Diagnosis was based on the structured interview supplemented by case note review and collateral history where available. All diagnoses were reviewed by an independent reviewer. Controls were ascertained with informed consent from the Irish GeneBank and represented blood donors who met the same ethnicity criteria as cases. Controls were not specifically screened for psychiatric illness.

**Blackwood, D | 18711365 [PGC1] | Edinburgh, UK | bip_edi1_eur**

This sample comprised Caucasian individuals contacted through the inpatient and outpatient services of hospitals in Southeast Scotland. A BD-I diagnosis was based on an interview with the patient using the SADS-L supplemented by case note review and frequently by information from medical staff, relatives and caregivers. Final diagnoses, based on DSM-IV criteria, were reached by consensus between two trained psychiatrists. Ethnically matched controls from the same region were recruited through the South of Scotland Blood Transfusion Service. Controls were not directly screened to exclude those with a personal or family history of psychiatric illness. The study was approved by the Multi-Centre Research Ethics Committee for Scotland and patients gave written informed consent for the collection of DNA samples for use in genetic studies.

**Kelsoe, J | 21926972 [PGC1] | USA (GAIN) | bip_gain_eur**

*Genetic Association Information Network (GAIN)/ The Bipolar Genome Study (BiGS)* The BD sample was collected under the auspices of the NIMH Genetics Initiative for BD (http://zork.wustl.edu/nimh/), genotyped as part of GAIN and analyzed as part of a larger GWAS conducted by the BiGS consortium. Approximately half of the GAIN sample was collected as multiplex families or sib pair families (waves 1-4), the remainder were collected as individual cases (wave 5). Subjects were ascertained at 12 sites: Indiana University, Johns Hopkins University, the NIMH Intramural Research Program, Washington University at St. Louis, University of Pennsylvania, University of Chicago, Rush Medical School, University of Iowa, University of California, San Diego, University of California, San Francisco, Howard University, and University of Michigan. All investigations were carried out after the review of protocols by the IRB at each participating institution. At all sites, potential cases were identified from screening admissions to local treatment facilities and through publicity programs or advocacy groups. Potential cases were evaluated using the DIGS[56], FIGS[57], and information from relatives and medical records. All information was reviewed through a best estimate diagnostic procedure by two independent and non-interviewing clinicians, and a consensus best-estimate diagnosis was reached. In the event of a disagreement, a third review was done to break the tie. Controls were from the NIMH Genetic Repository sample obtained by Dr. P. Gejman through a contract to Knowledge Networks, Inc. Only individuals with complete or near-complete psychiatric questionnaire data who did not fulfill diagnostic criteria for major depression and denied a history of psychosis or BD were included as controls for BiGS analyses. Controls were matched for gender and ethnicity to the cases.

**Scott, L; Myer, RM; Boehnke, M | 19416921 [PGC1] | Michigan, USA (Pritzker and NIMH) | bip_mich_eur**

The Pritzker Neuropsychiatric Disorders Research Consortium (NIMH/Pritzker) case and control samples were from the NIMH Genetics Initiative Genetics Initiative Repository. Cases were diagnosed according to DSM-III or DSM-IV criteria using diagnostic interviews and/or medical record review. Cases with low confidence diagnoses were excluded. From each wave 1-5 available non-Ashkenazi European-origin family, two BD1 siblings were included when possible and the proband was preferentially included if available (*n*=946 individuals in 473 sibling pairs); otherwise, a single BD1 case was included (*n*=184). The bipolar sibling pairs were retained within the NIMH/Pritzker sample when individuals in more than one study were uniquely assigned to a study set. Controls had non-Ashkenazi European origin, were aged 20-70 years and reported no diagnosis with or treatment for BD or schizophrenia, and that they had not heard voices that others could not hear. Individuals with suspected major depression were excluded based on answers to questions related to depressive mood. NIMH controls were further selected as the best match(es) to NIMH cases based on self-reported ancestry.

**Sklar, P; Smoller, J | 18317468 [PGC1] | USA (STEP1) | bip_stp1_eur**

The Systematic Treatment Enhancement Program for Bipolar Disorder (STEP-BD) was a seven-site, national U.S., longitudinal cohort study designed to examine the effectiveness of treatments and their impact on the course of BD that enrolled 4,361 participants who met DSM-IV criteria for BD1, BD2, bipolar not otherwise specified (NOS), schizoaffective manic or bipolar type, or cyclothymic disorder based on diagnostic interviews. From the parent study, 2,089 individuals who were over 18 years of age with BD1 and BD2 diagnoses consented to the collection of blood samples for DNA. BD samples with a consensus diagnosis of BD1 were selected for inclusion in STEP1. Two groups of control samples from the NIMH repository were used. One comprised DNA samples derived from US Caucasian anonymous cord blood donors. The second were controls who completed the online self-administered psychiatric screen and were ascertained as described above, by Knowledge Networks Inc. For the second sample of controls only those without a history of schizophrenia, psychosis, BD or major depression with functional impairment were used.

**Sklar, P; Smoller, J | 18711365 [PGC1] | USA (STEP2) | bip_stp2_eur**

The STEP2 sample included BD-1 and BD-2 samples from the STEP-BD study described above along with BD-2 subjects from UCL study also described above. The controls samples for this study were from the NIMH repository as described above for the STEP1 study.

**Andreassen, OA | PMID:21926972 [PGC1], PMID:20451256 | Norway (TOP) | bip_top7_eur**

In the TOP study (Tematisk omrade psykoser), cases of European ancestry, born in Norway, were recruited from psychiatric hospitals in the Oslo region. Patients were diagnosed according to the SCID[58] and further ascertainment details have been reported. Healthy control subjects were randomly selected from statistical records of persons from the same catchment area as the patient groups. The control subjects were screened by interview and with the Primary Care Evaluation of Mental Disorders (PRIME-MD)[59]. None of the control subjects had a history of moderate/severe head injury, neurological disorder, mental retardation or an age outside the age range of 18-60 years. Healthy subjects were excluded if they or any of their close relatives had a lifetime history of a severe psychiatric disorder. All participants provided written informed consent, and the human subjects protocol was approved by the Norwegian Scientific-Ethical Committee and the Norwegian Data Protection Agency.

**McQuillin, A; Gurling, H | 18317468 [PGC1] | UCL (University College London), London, UK | bip_uclo_eur**

The UCL sample comprised Caucasian individuals who were ascertained and received clinical diagnoses of bipolar 1 disorder according to UK National Health Service (NHS) psychiatrists at interview using the categories of the International Classification of Disease version 10. In addition, bipolar subjects were included only if both parents were of English, Irish, Welsh or Scottish descent and if three out of four grandparents were of the same descent. All volunteers read an information sheet approved by the Metropolitan Medical Research Ethics Committee who also approved the project for all NHS hospitals. Written informed consent was obtained from each volunteer. The UCL control subjects were recruited from London branches of the National Blood Service, from local NHS family doctor clinics and from university student volunteers. All control subjects were interviewed with the SADS-L to exclude all psychiatric disorders.

**Craddock, N, Jones, I, Jones, L | 17554300 | WTCCC | bip_wtcc_eur_sr-qc**

Cases were all over the age of 17 yr, living in the UK and of European descent. Recruitment was undertaken throughout the UK and included individuals who had been in contact with mental health services and had a lifetime history of high mood. After providing written informed consent, participants were interviewed by a trained psychologist or psychiatrist using a semi-structured lifetime diagnostic psychiatric interview (Schedules for Clinical Assessment in Neuropsychiatry) and available psychiatric medical records were reviewed. Using all available data, best-estimate life-time diagnoses were made according to the RDC[60]. In the current study we included cases with a lifetime diagnosis of RDC bipolar 1 disorder, bipolar 2 disorder or schizo-affective disorder, bipolar type.

Controls were recruited from two sources: the 1958 Birth Cohort study and the UK Blood Service (blood donors) and were not screened for history of mental illness.

All cases and controls were recruited under protocols approved by the appropriate IRBs. All subjects gave written informed consent.

#### ======== PGC2 Samples ========

**Adolfsson, R | Not published | Umeå, Sweden | bip_ume4_eur**

Clinical characterization of the patients included the Mini-International Neuropsychiatric Interview (MINI[61]), the Diagnostic Interview for Genetic Studies (DIGS[56]), the Family Interview for Genetic Studies (FIGS[57]) and the Schedules for Clinical Assessment in Neuropsychiatry (SCAN)[62]. The final diagnoses were made according to the DSM-IV-TR and determined by consensus of 2 research psychiatrists. The unrelated Swedish control individuals, consisting of a large population-based sample representative of the general population of the region, were randomly selected from the 'Betula study'.

**Alda, M; Smoller, J | Not published | Nova Scotia, Canada; I2B2 controls | bip_hal2_eur**

The case samples were recruited from patients longitudinally followed at specialty mood disorders clinics in Halifax and Ottawa (Canada). Cases were interviewed in a blind fashion with the Schedule of Affective Disorders and Schizophrenia-Lifetime version (SADS-L)[63] and consensus diagnoses were made according to DSM-IV[64] and Research Diagnostic Criteria (RDC)[60]. Protocols and procedures were approved by the local Ethics Committees and written informed consent was obtained from all patients before participation in the study. Control subjects were drawn from the I2B2 (Informatics for Integrating Biology and the Bedside) project[65]. The study consists of de-identified healthy individuals recruited from a healthcare system in the Boston, MA, US area. The de-identification process meant that the Massachusetts General Hospital Institutional Review Board elected to waive the requirement of seeking informed consent as detailed by US Code of Federal Regulations, Title 45, Part 46, Section 116 (46.116).

**Andreassen, OA | Not published | Norway (TOP) | bip_top8_eur**

The TOP8 bipolar disorder cases and controls were ascertained in the same way as the bip_top7_eur (TOP7) samples described above and recruited from hospitals across Norway.

**Biernacka, JM; Frye, MA | 27769005 | Mayo Clinic, USA | bip_may1_eur**

Bipolar cases were drawn from the Mayo Clinic Bipolar Biobank[66]. Enrollment sites included Mayo Clinic, Rochester, Minnesota; Lindner Center of **HOPE**/University of Cincinnati College of Medicine, Cincinnati, Ohio; and the University of Minnesota, Minneapolis, Minnesota. Enrollment at each site was approved by the local Institutional Review Board, and all participants consented to use of their data for future genetic studies. Participants were identified through routine clinical appointments, from in-patients admitted in mood disorder units, and recruitment advertising. Participants were required to be between 18 and 80 years old and be able to speak English, provide informed consent, and have DSM-IV-TR diagnostic confirmation of type 1 or 2 bipolar disorder or schizoaffective bipolar disorder as determined using the SCID. Controls were selected from the Mayo Clinic Biobank[67]. Potential controls with ICD9 codes for bipolar disorder, schizophrenia or related diagnoses in their electronic medical record were excluded.

**Breen, G; Vincent, JB | 24387768; 19416921; 21926972 [PGC1] |London, UK; Toronto, Canada [BACC] |bip_bac1_eur**

The total case/control cohort (*N*=1922) includes 871 subjects from Toronto, Canada (*N*=431 cases (160 male; 271 female); *N*=440 controls (176 male; 264 female)), 1051 subjects from London, UK (*N*=538 cases (180 male; 358 female); *N*=513 controls (192 male; 321 female)). A summary of mean and median age at interview, age of onset, diagnostic subtypes (BD 1 versus BD 2), presence of psychotic symptoms, suicide attempt and family history of psychiatric disorders has been provided previously for both the Toronto and London cohorts[68]. From the Toronto site (Centre for Addiction & Mental Health (CAMH)), BD individuals and unrelated healthy controls matched for age, gender and ethnicity were recruited. Inclusion criteria for patients: a) diagnosed with DSM-IV/ICD 10 BD 1 or 2; b) 18 years old or over; c) Caucasian, of Northern and Western European origin, and three out of four grandparents also N.W. European Caucasian. Exclusion criteria include a) Use of intravenous drugs; b) Evidence of intellectual disability; c) Related to an individual already in the study; d) Manias that only ever occurred in relation to or resulting from alcohol or substance abuse/dependence, or medical illness; e) Manias resulting from non-psychotropic substance usage. The SCAN interview (Schedule for Clinical Assessments in Neuropsychiatry) was used for subject assessment[69]. Using the SCAN interview along with case note review, each case was assigned DSM-IV and ICD 10 diagnoses by two independent diagnosticians, according to lifetime consensus best-estimate diagnosis. Lifetime occurrence of psychiatric symptoms was also recorded using the OPCRIT checklist, modified for use with mood disorders. Similar methods and criteria were also used to collect a sample of 538 BD cases and 513 controls for the London cohort (King's College London; KCL)[70].

Both studies were approved by respective institutional research ethics committees (the CAMH Research Ethics Board (REB) in Toronto, and the College Research Ethics Committee (CREC) at KCL), and informed written consent was obtained from all participants. GWAS results have previously been published for the entire KCL/CAMH cohort[71].

**Rietschel, M; Nöthen, MM; Schulze, TG; Reif, A; Forstner, AJ | 24618891 | BOMA-Germany II | bip_bmg2_eur**

Cases were recruited from consecutive admissions to psychiatric in-patient units at the University Hospital Würzburg. All cases received a lifetime diagnosis of BD according to the DSM-IV criteria using a consensus best-estimate procedure based on all available information, including semi-structured diagnostic interviews using the Association for Methodology and Documentation in Psychiatry[72], medical records and the family history method. In addition, the OPCRIT system was used for the detailed polydiagnostic documentation of symptoms.

Control subjects were ascertained from the population-based Heinz Nixdorf Recall (HNR) Study[73]. The controls were not screened for a history of mental illness. Study protocols were reviewed and approved in advance by Institutional Review Boards of the participating institutions. All subjects provided written informed consent.

**Rietschel, M; Nöthen, MM; Schulze, TG; Bauer, M; Forstner, AJ; Müller-Myhsok, B | 24618891 | BOMA-Germany III | bip_bmg3_eur**[74]

Cases were recruited at the Central Institute of Mental Health in Mannheim, University of Heidelberg, and other collaborating psychiatric hospitals in Germany. All cases received a lifetime diagnosis of BD according to the DSM-IV criteria using a consensus best-estimate procedure based on all available information including structured diagnostic interviews using the AMDP, Composite International Diagnostic Screener (CID-S)[75], SADS-L and/or SCID, medical records, and the family history method. In addition, the OPCRIT system was used for the detailed polydiagnostic documentation of symptoms.

Controls were selected randomly from a Munich-based community sample and recruited at the Max-Planck Institute of Psychiatry. They were screened for the presence of anxiety and mood disorders using the CID-S. Only individuals without mood and anxiety disorders were collected as controls. Study protocols were reviewed and approved in advance by Institutional Review Boards of the participating institutions. All subjects provided written informed consent.

**Hauser, J; Lissowska, J; Forstner, AJ | 24618891 | BOMA-Poland | bip_bmpo_eur**

Cases were recruited at the Department of Psychiatry, Poznan University of Medical Sciences, Poznan, Poland. All cases received a lifetime diagnosis of BD according to the DSM-IV criteria based on a consensus best-estimate procedure and structured diagnostic interviews using the SCID. Controls were drawn from a population-based case-control sample recruited by the Cancer-Center and Institute of Oncology, Warsaw, Poland and a hospital-based case-control sample recruited by the Nofer Institute of Occupational Medicine, Lodz, Poland. The Polish controls were produced by the International Agency for Research on Cancer (IARC) and the Centre National de Génotypage (CNG) GWAS Initiative for a study of upper aerodigestive tract cancers. The controls were not screened for a history of mental illness. Study protocols were reviewed and approved in advance by Institutional Review Boards of the participating institutions. All subjects provided written informed consent.

**Rietschel, M; Nöthen, MM; Rivas, F; Mayoral, F; Kogevinas, M; others | 24618891 | BOMA-Spain | bip_bmsp_eur**

Cases were recruited at the mental health departments of the following five centers in Andalusia, Spain: University Hospital Reina Sofia of Córdoba, Provincial Hospital of Jaen; Hospital of Jerez de la Frontera (Cádiz); Hospital of Puerto Real (Cádiz); Hospital Punta Europa of Algeciras (Cádiz); and Hospital Universitario San Cecilio (Granada). Diagnostic assessment was performed using the SADS-L; the OPCRIT; a review of medical records; and interviews with first and/or second degree family members using the Family Informant Schedule and Criteria (FISC)[76]. Consensus best estimate BD diagnoses were assigned by two or more independent senior psychiatrists and/or psychologists, and according to the RDC, and the DSM-IV. Controls were Spanish subjects drawn from a cohort of individuals recruited in the framework of the European Community Respiratory Health Survey (ECRHS, http://www.ecrhs.org/). The controls were not screened for a history of mental illness. Study protocols were reviewed and approved in advance by Institutional Review Boards of the participating institutions. All subjects provided written informed consent.

**Fullerton, J.M.; Mitchell, P.B.; Schofield, P.R.; Martin N.G.; Cichon, S. | 24618891 | BOMA-Australia | bip_bmau_eur**

Cases were recruited at the Mood Disorder Unit, Prince of Wales Hospital in Sydney. All cases received a lifetime diagnosis of BD according to the DSM-IV criteria based on a consensus best-estimate procedure[14] and structured diagnostic interviews using the DIGS, FIGS, and the SCID. Controls were parents of unselected adolescent twins from the Brisbane Longitudinal Twin Study. The controls were not screened for a history of mental illness. Study protocols were reviewed and approved in advance by Institutional Review Boards of the participating institutions. All subjects provided written informed consent.

**Grigoroiu-Serbanescu, M; Nöthen, MM | 21353194 | BOMA-Romania | bip_rom3_eur**

Cases were recruited from consecutive admissions to the Obregia Clinical Psychiatric Hospital, Bucharest, Romania. Patients were administered the DIGS[77] and FIGS[57] interviews. Information was also obtained from medical records and close relatives. The diagnosis of BP-I was assigned according to DSM-IV criteria using the best estimate procedure.  All patients had at least two hospitalized illness episodes. Population-based controls were evaluated using the DIGS to exclude a lifetime history of major affective disorders, schizophrenia, schizoaffective disorders, and other psychoses, obsessive-compulsive disorder, eating disorders, and alcohol or drug addiction.

**Kelsoe, J; Sklar, P; Smoller, J | [PGC1 Replication] | USA (FAT2; FaST, BiGS, TGEN) | bip_fat2_eur**

Cases were collected from individuals at the 11 U.S. sites described for the GAIN sample.  Eligible participants were age 18 or older meeting DSM-IV criteria for BD-I or BD-II by consensus diagnosis based on interviews with the Affective Disorders Evaluation (ADE) and MINI. All participants provided written informed consent, and the study protocol was approved by IRBs at each site. Collection of phenotypic data and DNA samples were supported by NIMH grants MH063445 (JW Smoller); MH067288 (PI: P Sklar), MH63420 (PI: V Nimgaonkar) and MH078151, MH92758 (PI: J. Kelsoe). The control samples were NIMH controls that were using the methods described in that section. The case and control samples were independent of those included in the GAIN sample.

**Kirov, G | 25055870 | Bulgarian trios | bip_butr_eur**

All cases were recruited in Bulgaria from psychiatric inpatient and outpatient services. Each proband had a history of hospitalization and was interviewed with an abbreviated version of the SCAN. Consensus best-estimate diagnoses were made according to DSM-IV criteria by two researchers. All participants gave written informed consent, and the study was approved by local ethics committees at the participating centers.

**Kirov, G | 25055870 | UK trios | bip_uktr_eur**

The BD subjects were recruited from lithium clinics and interviewed in person by a senior psychiatrist, using the abbreviated version of the SCAN. Consensus best-estimate diagnoses were made based on the interview and hospital notes. Ethics committee approval for the study was obtained from the relevant research ethics committees and all individuals provided written informed consent for participation.

**Landén, M; Sklar, P | [ICCBD] | Sweden (ICCBD) | bip_swa2_eur**

The BD subjects were identified using the Swedish National Quality Register for Bipolar Disorders (BipoläR) and the Swedish National Patient Register (using a validated algorithm[78] requiring at least two hospitalizations with a BD diagnosis). A confirmatory telephone interview with a diagnostic review was conducted. Additional subjects were recruited from the St. Göran Bipolar Project (Affective Center at Northern Stockholm Psychiatry Clinic, Sweden), enrolling new and ongoing patients diagnosed  with BD using structured clinical interviews. Diagnoses were made according to the DSM-IV criteria (BipoläR and St. Göran Bipolar Project) and ICD-10 (National Patient Register). The control subjects used were the same as for the SCZ analyses described above. All ascertainment procedures were approved by the Regional Ethical Committees in Sweden.

**Landén, M; Sklar, P | [ICCBD] | Sweden (ICCBD) | bip_swei_eur**

The cases and controls in the bip_swei_eur sample were recruited using the same ascertainment methods described for the bip_swa2_eur sample.

**Leboyer, M |[79]; [PGC1 replication] | France | bip_fran_eur**

Cases with BD1 or BD2 and control samples were recruited as part of a large study of genetics of BD in France (Paris-Creteil, Bordeaux, Nancy) with a protocol approved by relevant IRBs and with written informed consent. Cases of French descent for more than 3 generations were assessed by a trained psychiatrist or psychologist using structured interviews supplemented by medical case notes, mood scales and self-rating questionnaire assessing dimensions.

**Li, Q | 24166486; 27769005 | USA (Janssen), SAGE controls | bip_jst5_eur**

The study included unrelated patients with bipolar 1 disorder from 6 clinical trials (IDs: NCT00253162, NCT00257075, NCT00076115, NCT00299715, NCT00309699, and NCT00309686). Participant recruitment was conducted by Janssen Research & Development, **LLC** (formerly known as Johnson & Johnson Pharmaceutical Research & Development, **LLC**) to assess the efficacy and safety of risperidone. Bipolar cases were diagnosed according to DSM-IV-TR criteria. The diagnosis of bipolar disorder was confirmed by the Schedule for Affective Disorders and Schizophrenia for School-Age Children-Present and Lifetime Version (K-SADS-PL) in NCT00076115, by the SCID in NCT00257075 and NCT00253162, or by the MINI in NCT00299715 and NCT00309699, and NCT00309686, respectively. Additional detailed descriptions of these clinical trials can be found at ClinicalTrials.gov. Only patients of European ancestry with matching controls were included in the current analysis. Control subjects were drawn from the Study of Addiction: Genetics and Environment (**SAGE**, dbGaP Study Accession: phs000092.v1.p1). Control subjects did not have alcohol dependence or drug dependence diagnoses; however, mood disorders were not an exclusion criterion.

**Craddock, N; Jones, I; Jones, L | [ICCBD] | Cardiff and Worcester, UK (ICCBD-BDRN) | bip_icuk_eur**

Cases were all over the age of 17 yr, living in the UK and of European descent. Cases were recruited using systematic and not systematic methods as part of the Bipolar Disorder Research Network project (www.bdrn.org), provided written informed consent and were interviewed using a semi-structured diagnostic interview, the Schedules for Clinical Assessment in Neuropsychiatry. Based on the information gathered from the interview and case notes review, best-estimate lifetime diagnosis was made according to DSM-IV. Inter-rater reliability was formally assessed using 20 randomly selected cases (mean κ Statistic = **0.85**). In the current study we included cases with a lifetime diagnosis of DSM-IV bipolar disorder or schizo-affective disorder, bipolar type. The BDRN study has UK National Health Service (NHS) Research Ethics Committee approval and local Research and Development approval in all participating NHS Trusts/Health Boards.Controls were part of the Wellcome Trust Case Control Consortium common control set, which comprised healthy blood donors recruited from the UK Blood Service and samples from the 1958 British Birth Cohort. Controls were not screened for a history of mental illness. All cases and controls were recruited under protocols approved by the appropriate IRBs. All subjects gave written informed consent.

**Ophoff, RA | Not Published | Netherlands | bip_ucla_eur**

The case sample consisted of inpatients and outpatients recruited through psychiatric hospitals and institutions throughout the Netherlands. Cases with DSM-IV bipolar disorder, determined after interview with the SCID, were included in the analysis. Controls were collected in parallel at different sites in the Netherlands and were volunteers with no psychiatric history after screening with the (MINI[61]). Ethical approval was provided by UCLA and local ethics committees, and all participants gave written informed consent.

**Paciga, S | [PGC1] | USA (Pfizer) | bip_pf1e_eur**

This sample comprised Caucasian individuals recruited into one of three Geodon (ziprasidone) clinical trials (NCT00141271, NCT00282464, NCT00483548). Subjects were diagnosed by a clinician with a primary diagnosis of Bipolar 1 Disorder, most recent episode depressed, with or without rapid cycling, without psychotic features, as defined in the DSM-IV-TR (296.5x) and confirmed by the MINI (version 5.0.0). Subjects also were assessed as having a HAM-D-17 total score of >20 at the screening visit. The trials were conducted in accordance with the protocols, International Conference on Harmonization of Good Clinical Practice Guidelines, and applicable local regulatory requirements and laws. Patients gave written informed consent for the collection of blood samples for DNA for use in genetic studies.

**Pato, C | [ICCBD] | Los Angeles, USA (ICCBD-GPC)| bip_usc2_eur**

Genomic Psychiatry Consortium (**GPC**) cases and controls were collected using the University of Southern California healthcare system, as previously described[80]. Using a combination of focused, direct interviews and data extraction from medical records, diagnoses were established using the OPCRIT and were based on DSM-IV-TR criteria. Age and gender-matched controls were ascertained from the University of Southern California health system and assessed using a validated screening instrument and medical records.

**======== PGC2 Followup Samples ========**

**Kelsoe, J | [PGC1] | USA (BiGS/TGEN1) | TGEN1_eur**

Cases and controls for this sample were ascertained using the same procedures applied for the bip_gain_eur sample described above. These samples formed a distinct **PCA** cluster from the samples described above and were therefore analyzed separately.

**Li, Q | 24166486 | various Eastern Europe, shared T. Esku controls | JJ_EAST_eur**

The cases were drawn from the same six clinical studies described for bip_jst5_eur except that only patients of east European ancestry with matching controls were included in this cohort. Most of the Eastern European controls were from the Estonian Biobank project (EGCUT)[81] and were ancestrally matched with cases.

**Schulze, T | [ConLiGen] | Germany | BIP_KFO_eur**

The KFO sample was derived from the Clinical Research Group 241 (KFO241 consortium; www.kfo241.de) and the PsyCourse consortium (www.psycourse.de). The samples form part of a multi-site German/Austrian longitudinal study. Diagnoses were made according to DSM-IV. German Red Cross controls were collected by the Central Institute for Mental Health in Mannheim, University of Heidelberg, Germany. Volunteers who gave blood to the Red Cross were asked whether they would be willing to participate in genetic studies of psychiatric disorders. Control subjects were not selected based on mental health screening.

======== External studies **PGC3** ========

**Milani L | 24518929 | Estonia (Estonian Biobank) | EstonianBiobank**

The Estonian Biobank (EstBB) is a population-based cohort of 200,000 participants with a rich variety of phenotypic and health-related information collected for each individual[81]. At recruitment, all participants signed a consent to allow follow-up linkage of their electronic health records (EHR), thereby providing a longitudinal collection of phenotypic information. Health records have been extracted from the national Health Insurance Fund Treatment Bills (from 2004), Tartu University Hospital (from 2008), and North Estonia Medical Center (from 2005). The diagnoses are coded in ICD-10 format and drug dispensing data include drug ATC codes, prescription status and purchase date (if available). For the current study, cases of bipolar disease were determined by searching the EHRs for data on F31* ICD-10 diagnosis. All remaining participants who did not have any ICD-10 F* group diagnoses were defined as controls. Cases with bipolar I disorder were those with ICD codes of F31.1 and F31.2.

**Zwart JA | Unpublished | Norway (the Trøndelag Health Study) | HUNT**

The HUNT sample consisted of 905 subjects with BD and 41,914 population controls[82]. Patients and controls were of European ancestry and were recruited from the Nord-Trøndelag County, Norway. Diagnoses were assigned according to ICD-9 or ICD-10. The controls included individuals not diagnosed with substance use disorders, schizophrenia, bipolar disorder, major depressive disorder, anxiety disorders, eating disorders, personality disorders, or ADHD in hospitals (ICD-9 or ICD-10) or general practice (ICPC2). They also were >40 years of age, had low self-reported levels of anxiety and depression (HADS-A and HADS-D ≤ 11), and reported no use of antidepressants, anxiolytics, or hypnotics. Approval for the study was granted by the Data Inspectorate of Norway, the Health Directorate and the Regional Committee for Medical and Health Research Ethics. Cases of bipolar I disorder were those with ICD codes of F31.1, F31.2 or F31.6 and individuals with an ICD-9 code of 295 or ICD-10 codes F20-F29 were excluded. Cases of bipolar II disorder were those with ICD codes of F31.8 and individuals with an ICD-9 code of 295 or ICD-10 codes F20-F29, F31.1-.2 or F31.6 were excluded.

**======== PGC PsychChip Samples ========**

**Pato, C | Not published | [PGC Psychchip] | gpcw1**

The cases and controls in this study were ascertained in the same manner as those described above for bip_usc2_eur.

**Reif, A | Not published | [PGC Psychchip] | germ1**

Cases were recruited in the same manner as those described above for BOMA-Germany II | bip_bmg2_eur. Control subjects were healthy participants who were recruited from the community of the same region as cases. They were of Caucasian descent and fluent in German. Exclusion criteria were manifest or lifetime DSM-IV axis I disorder, severe medical conditions, intake of psychoactive medication as well as alcohol abuse or abuse of illicit drugs. Absence of DSM-IV axis I disorder was ascertained using the German versions of the Mini International Psychiatric Interview. IQ was above 85 as ascertained by the German version of the Culture Fair Intelligence Test 2[83]. Study protocols were reviewed and approved by the ethical committee of the Medical Faculty of the University of Würzburg. All subjects provided written informed consent.

**Serretti, A, Vieta E, Ribases M | Not published | [PGC Psychchip] | spsp3**

The sample includes 267 BD subjects (Spanish Wave2 Serretti PsychChip QC Summary), of which 180 Spanish and 87 Italian. Spanish sample: 180 subjects were enrolled in a naturalistic cohort study, consecutively admitted to the out-patient Bipolar Disorders Unit, Hospital Clinic, University of Barcelona. This is a systematic cross-sectional analysis deeply described in a previous paper on the same sample investigating rs10997870 **SIRT1** gene variant[84]. Inclusion criteria were a diagnosis of bipolar disorder (type 1 or 2) according to DSM-IV TR criteria and age of 18 years or older. The study was approved by the local ethical committee and carried out in accordance with the ethical standards laid down in the Declaration of Helsinki. Signed informed consent was obtained from all participants after a detailed and extensive description of the study and patient's confidentiality was preserved. The current and lifetime diagnoses of mental disorders were formulated by independent senior psychiatrists (diagnostic concordance: Kappa=**0.80**) according to DSM-IV TR clinical criteria and confirmed through the semi-structured interviews for Axis I disorders according to DSM IV TR criteria (SCID I). Furthermore, all available clinical data coming from follow-up at our unit and collateral information concerning illness history were cross-referred to ensure accuracy and obtain complete clinical information. Specific psychopathological dimensions were assessed by means of rating scales and clinical questionnaires administered by clinicians, adequately trained to enhance inter-rater reliability. Mood episodes were defined according to DSM-IV TR criteria, and their severity was measured through the administration of the 21-item Hamilton Depression Rating Scale (HDRS-21, Spanish version). The most severe depressive episode was defined based on the severity at the HDRS (total score > 14) and clinical judgment. Italian sample: 87 subjects with bipolar depression were enrolled into the study when admitted at the Department of Psychiatry, University of Bologna, Italy. A description of the subjects has been previously reported when analyzing clinical features[85]. Inclusion criteria were a diagnosis of bipolar disorder, most recent episode depressive as assessed by DSM-IV-TR criteria; Young Mania Rating Scale (YMRS) score <12; Hamilton Depression Rating Scale (HAM-D) <12. Exclusion criteria were presence of a bipolar disorder, most recent episode manic or hypomanic; presence of severe medical conditions; presence of moderate to severe dementia (Mini Mental State Examination score <20). The following scales were administered biweekly during the hospitalization: HAM-D, Hamilton Anxiety Rating Scale (HAM-A), YMRS and Dosage Record and Treatment Emergent Symptom Scale (DOTES). Written informed consent was obtained for each patient recruited. The study protocol was approved by the local Ethical

Committee, and it has been performed in accordance with the ethical standards laid down in the 1975 Declaration of Helsinki.

The Spanish controls were part of the Mental-Cat clinical sample or the INSchool population-based cohort. A total of 1,774 controls from the Mental-Cat cohort (60.5% males) were evaluated and recruited prospectively from a restricted geographic area at the Hospital Universitari Vall d'Hebron of Barcelona (Spain) and consisted of unrelated healthy blood donors. The INSchool sample consisting of 771 children (76.**2%** males) from schools in Catalonia were involved for screening using the Achenbach System of Empirically Based Assessment (ASEBA) with the Child Behavior Checklist CBCL/4-18 (completed by parents or surrogates), the Teacher Report Form TRF/5-18 (completed by teachers and other school staff) and the Youth Self-Report YSR/11-18 (completed by youths); the Strengths and Difficulties Questionnaire (SDQ) and the Conner's ADHD Rating Scales (Parents and Teachers). Genomic DNA samples were obtained either from peripheral blood lymphocytes by the salting out procedure or from saliva using the Oragene DNA Self-Collection Kit (DNA Genotek, Kanata, Ontario Canada). DNA concentrations were determined using the Pico- Green dsDNA Quantitation Kit (Molecular Probes, Eugene, OR) and genotyped with the Illumina Infinium PsychArray-24 v1.1 at the Genomics Platform of the Broad Institute. The study was approved by the Clinical Research Ethics Committee (CREC) of Hospital Universitari Vall d'Hebron, all methods were performed in accordance with the relevant guidelines and regulations, and written informed consent was obtained from participant parents before inclusion into the study. Detailed information has been published previously[86].

**Perlis, R; Sklar, P; Smoller, J, Goes F, Mathews CA, Waldman I | Not published | [PGC Psychchip] | usaw4**

Perlis, R; Sklar, P; Smoller, J: EHR data were obtained from a health care system of more than 4.6 million patients[87] spanning more than 20 years. Experienced clinicians reviewed charts to identify text features and coded data consistent or inconsistent with a diagnosis of bipolar disorder. Natural language processing was used to train a diagnostic algorithm with **95%** specificity for classifying bipolar disorder. Filtered coded data were used to derive three additional classification rules for case subjects and one for control subjects. The positive predictive value (PPV) of EHR-based bipolar disorder and subphenotype diagnoses was calculated against diagnoses from direct semistructured interviews of 190 patients by trained clinicians blind to EHR diagnosis. The PPV of bipolar disorder defined by natural language processing was **0.86**. Coded classification based on strict filtering achieved a value of **0.84**; however, classifications based on less stringent criteria performed less well. No EHR-classified control subject received a diagnosis of bipolar disorder based on direct interview (PPV=1.0). For most subphenotypes, PPV exceeded **0.80**. The EHR-based classifications were used to accrue bipolar disorder cases and controls for genetic analyses. Samples were genotyped on the Psychchip array.

Goes, FS: Cases represented independent probands from a European American family sample that was collected at Johns Hopkins University from 1988-2010. Families had at least 2 additional relatives with a major mood disorder (defined as bipolar disorder type 1, bipolar type 2 or recurrent major depressive disorder). Diagnostic interviews were performed using the Schedule for Affective Disorders and Schizophrenia-Lifetime Version (*N*=81) and the Diagnostic Instrument for Genetics Studies (*N*=161). All cases underwent best-estimate diagnostic procedures. After genotyping quality control there were 242 cases, of which 240 were diagnosed as bipolar disorder type 1 and 2 as schizoaffective disorder, bipolar type. Diagnoses were based on DSM-III and DSM-IV criteria. Probands from this sample have been previously studied in family based linkage and exome studies.[88–90]

Mathews CA: Control samples were ascertained as part of ongoing genetic and neurophysiological studies of hoarding, obsessive compulsive and tic disorders. Controls reported no current or lifetime history of mania or hypomania at the time of ascertainment. Sixty-two of the 104 controls were screened for psychiatric illness using the Structured Clinical Interview for DSM-IV TR diagnoses and diagnoses of bipolar disorder, lifetime or current, were ruled out through a best estimate consensus diagnosis. Other psychiatric diagnoses were not excluded. The remaining 42 participants were not formally screened; however, reported no lifetime or current history of bipolar disorder, obsessive compulsive, hoarding, or tic disorders. Samples were genotyped on the Psychchip array. Ethical approvals were obtained from the University of Florida Human Subjects Review Board.

Waldman I: Control samples were ascertained as part of an ongoing genetic study of ADHD and other Externalizing disorders (I.e., Oppositional Defiant Disorder and Conduct Disorder). Controls reported no current diagnoses of Externalizing or Internalizing disorders at the time of ascertainment. Controls were assessed for psychiatric conditions using the Emory Diagnostic Rating Scale (EDRS)[91], a questionnaire that assessed parent ratings of symptoms of common

DSM-IV Externalizing and Internalizing disorders (e.g., Major Depressive Disorder and various anxiety disorders). Samples were genotyped on the Psychchip array. Ethical approvals were obtained from the Emory University and University of Arizona Human Subjects Review Boards.

**Baune, BT; Dannlowski, U | Not published | [PGC Psychchip] | bdtrs**

The Bipolar Disorder treatment response Study (BP-TRS) comprises BD inpatient cases and screened controls of Caucasian background. Psychiatric diagnosis of bipolar disorders was ascertained using SCID or MINI 6.0 using DSM-IV criteria in a face-to-face interview by a trained psychologist / psychiatrist for both cases and controls. Healthy controls were included if no current or lifetime psychiatric diagnosis was identified. Cases were included if current or lifetime diagnosis of bipolar disorder was ascertained by structured diagnostic interview. Cases and controls are of similar age range (≥18 yrs of age) and were collected from the same geographical areas. Other assessments including symptom ratings, psychiatric history, treatment history, treatment response were based on interview and carried out by trained psychologists/psychiatrists. Samples were genotyped on the Psychchip array. Ethical approval was obtained from the University of Münster Human Ethics Committee, Münster, Germany.

**Ophoff R, Posthuma D, Lochner C, Franke B | Not published | [PGC Psychchip] | dutch**

Ophoff R: Cases and controls were collected using the same protocol as described above for the "ucla" sample.

Lochner C: Controls include South African Caucasian population based-controls ascertained from blood banks and controls recruited through university campuses and newspaper advertisements, who underwent a psychiatric interview and had no current or lifetime psychiatric disorder[92,93].

Franke B: The controls included are healthy individuals from the Dutch part of the International Multicenter ADHD Genetics (IMAGE) project[94,95].

Posthuma D: Data were provided for 960 unscreened Dutch population controls from the Netherlands Study of Cognition, Environment and Genes (NESCOG)[96]. The study was approved by the institutional review board of Vrije Universiteit Amsterdam and participants provided informed consent.

**Gawlik M | Not published | [PGC Psychchip] | gawli**

Patients were recruited at the Department of Psychiatry, Psychosomatics and Psychotherapy, University of Würzburg, Germany. Diagnosis according to DSM-IV (Diagnostic and Statistical Manual of Mental Disorders-fourth edition) was made by the best estimate lifetime diagnosis method, based on all available information, including medical records, and the family history method.

**Fullerton J, Mitchell PB, Schofield PR, Green MJ, Weickert CS, Weickert TW, The Australian Schizophrenia Research Bank | Not published | [PGC Psychchip] | neuc1**

The NeuRA collection comprised BD cases from three cohorts ascertained in Australia: the bipolar high risk study[97] (*n*=97), the Imaging Genetics in Psychosis Study (IGP; *n*=47)[98] and a clinic sample (*n*=109) recruited using the Sydney Bipolar Disorders Clinic[99]. The clinic sample used the same ascertainment procedures as described for the bip_bmau_eur sample. The bipolar high-risk study is a collaborative study with 4 US and one Australian groups, with young participants aged 12-30. The IGP sample was recruited from outpatient services of the Southeastern Sydney-Illawarra Area Health Service (SESIAHS), the Sydney Bipolar Disorders Clinic and the Australian Schizophrenia Research Bank. Healthy controls were sourced from the high risk, IGP and the Cognitive and Affective Symptoms of Schizophrenia Intervention (CASSI) trial[100] studies, and were recruited from the community, had no personal lifetime history of a DSM-IV Axis-I diagnosis as determined by psychiatric interview, and no history of psychotic disorders among first-degree biological relatives. Additional controls were recruited as part of the strategy to develop an Australian Schizophrenia Research Biobank for studies into the genetics of this disease. The ascertainment of these controls has been previously described[101].

**Landén M, Hillert J, Alfredsson L | Not published | [PGC Psychchip] | swed1**

The cases in the swed1 sample were recruited using the same ascertainment methods described for the bip_swa2_eur sample. Population-based healthy controls, randomly selected from the Swedish national population register, were collected as part of two case-control studies of multiple sclerosis: GEMS (Genes and Environment in Multiple Sclerosis) and EIMS (Epidemiological Investigation of Multiple Sclerosis)[102].

**Di Florio A, McQuillin A, McIntosh A, Breen G | Not published | [PGC Psychchip] | ukwa1**

McQuillin A: BD cases were recruited using the same protocol as the bip_uclo_eur described above. A subset (*n*=448) of the control subjects were random UK blood donors obtained from the ECACC DNA Panels (https://www.phe-culturecollections.org.uk/products/dna/hrcdna/hrcdna.jsp). The remaining control subjects (*n*=814) had been screened for an absence of mental illness in using the same protocol as the bip_uclo_eur described above.

Di Florio A: Cases were recruited across the United Kingdom in the same manner as described for the bip_wtcc_eur and bip_icuk_eur samples.

McIntosh AM: BD cases were recruited from the clinical case loads of treating psychiatrists from Edinburgh and across the central belt of Scotland. Controls were identified from non-genetic family members and from the extended networks of the participants themselves. All participants were of European ancestry, and diagnosis was confirmed using an established battery developed for ICCCBD. Breen G: Controls were drawn from blood donors to the UK Motor Neuron Disease Association DNA Biobank[103]

**Perlis, R; Sklar, P; Smoller, J, Nievergelt C, Kelsoe J | Not published | [PGC Psychchip] | usaw5**

Kelsoe, J: The Pharmacogenomics of Bipolar Disorder (**PGBD**) study was a prospective assessment of lithium response in BDI patients. The goal was to identify genes for lithium response. Subjects were recruited from clinics at 11 international sites and followed for up to 2.5 years. Diagnosis was obtained by DIGS interview and medical records reviewed by blind experienced clinicians. As the comparison was between lithium responders and non-responders, no controls were collected. All subjects provided written informed consent.

Perlis R: Cases of bipolar disorder were individuals treated with lithium drawn from the Partners Healthcare electronic health record database, (which spans two large academic medical centers, Massachusetts General Hospital and Brigham and Women's Hospital in addition to community and specialty outpatient clinics)[104]. Any patients aged 18 years or older with at least one lithium prescription between 2006 and 2013 based on e-prescribing data were included. The Partners Institutional Review Board approved all aspects of this study. Individuals with a diagnosis of schizophrenia based on ICD9 codes were excluded.

Smoller J: Cases and controls were recruited in the same manner as described above for "usaw4".

======== **PGC3 Samples** ========

**Ferentinos P, Dikeos D, Patrinos G | Not published | Greece (Attikon General Hospital) | greek**

All adult patients with a DSM-IV-TR/DSM-5 diagnosis of bipolar disorder hospitalized at the inpatient unit or followed-up at the specialized 'Affective disorders and Suicide' outpatient clinic of the 2nd Department of Psychiatry, National and Kapodistrian University of Athens, Attikon General Hospital, Athens, Greece from 2012 to 2017 were recruited for the current study. Patients were referred to the specialized 'Affective disorders and Suicide' outpatient clinic either from the inpatient unit after hospitalization or from the community. Diagnosis was established and demographic (age, gender, family status, profession, employment status, education) and relevant clinical features (e.g. age at onset, polarity of first and most recent episode, number of lifetime depressive and manic/hypomanic episodes, number of hospitalizations, lifetime suicidality, lifetime psychosis) were extracted through a M.I.N.I.-5.0.0-based semi-structured diagnostic interview, which was administered during patients' initial clinical assessment and regularly updated ever since, interviews of primary caregivers and inspection of medical records. Lifetime presence of any DSM-IV-TR axis I psychiatric comorbidities (dysthymia, panic disorder, agoraphobia, social phobia, generalized anxiety disorder, obsessive-compulsive disorder, post-traumatic stress disorder, alcohol and substance abuse and dependence, anorexia nervosa, bulimia nervosa) was similarly extracted. Family history of major psychiatric disorders and suicidality in first- and second-degree relatives was recorded with a specific questionnaire based on the Family Interview for Genetic Studies. Medical comorbidities were recorded with the Cumulative Illness Rating Scale, completed based on interview with patient and primary caregivers, inspection of patient's medical records and laboratory exams (basic or specific, if considered necessary). Presence of selected medical diseases was specifically recorded.

Control (unaffected) participants were a convenient sample drawn from the same geographic area as case participants, either within health care facilities or as community volunteers. All of them went through a brief clinical interview including items on psychiatric and medical history, psychiatric family history, past and current medical or psychiatric therapies, and a brief mental state examination. Only participants found to be free of lifetime major mental disorders (MDD,

BD, schizophrenia, or other psychotic disorders) and with no family history of major mental disorder in their first-degree relatives were recruited as controls.

All cases and controls were native Greek speakers. All participants provided written informed consent before being included in the study and the study protocol was approved by the Research Ethics Committee of Attikon General Hospital.

**Andreassen, OA | Not published | Norway (TOP) | norgs**

The NORGS bipolar disorder cases and controls were ascertained in the same way as the bip_top7_eur (TOP7) samples described above and recruited from hospitals across Norway.

**Andreassen, OA | Not published | Norway (TOP) | noroe**

The NOROE bipolar disorder cases and controls were ascertained in the same way as the bip_top7_eur (TOP7) samples described above and recruited from hospitals across Norway.

**Reininghaus EZ | Not published| Austria (Medical University of Graz) | graza**

Univ. Prof. DDr. Eva Reininghaus, Priv.Doz. DDr. Susanne Bengesser, Priv.Doz. Dr. Nina Dalkner, Dr. Frederike Fellendorf and further team members of the special outpatient's department for bipolar affective disorders at the Department of Psychiatry and Psychotherapeutic Medicine, Medical University of Graz, Austria: Cases with bipolar affective disorder (type I and II) and healthy controls were recruited at the Department of Psychiatry and Psychotherapeutic Medicine at the Medical University of Graz (MUG), Austria. Study protocols were approved by the ethics committee of the Medical University of Graz. Patients and healthy controls gave written informed consent, and the study was conducted according to the declaration of Helsinki. All patients received a clinical interview by a psychiatrist or psychologist and a diagnosis according to DSM-IV with the SCID-I (Structured clinical interview). Healthy controls did not have a history of a psychiatric disorder. Furthermore, healthy controls did not have any first- or second-degree relatives with a psychiatric disorder. The PGC-Graz sample ($n$= 244; 114 males, 130 females) includes 167 cases with bipolar disorder and 77 healthy controls genotyped with Omniexpress 1.2 by Illumina.

**Grigoroiu-Serbanescu M | 31791676; 26806518 | Romania (BOMA-Romania) | bmrom**

This sample includes the BOMA-Romania sample and additional cases from the ConLiGen-Romania sample. For the BOMA-Romania sample, unrelated BP-I patients were recruited from consecutive admissions in the Obregia Psychiatric Hospital of Bucharest, Romania. All participants provided written informed consent following a detailed explanation of the study aims and procedures. The study was performed in accordance with the Code of Ethics of the World Medical Association (Declaration of Helsinki). All participants were of Romanian descent according to self-reported ancestry. Genealogical information about parents and all four grandparents was obtained through direct interview of the subjects.

The patients were investigated with the Diagnostic Interview for Genetic Studies (DIGS)[77] and the Family Interview for Genetic Studies (FIGS)[57] The diagnosis of BP-I was assigned according to DSM-IV criteria on the basis of both the DIGS and medical records. Patients were included in the sample if they had at least two documented hospitalized illness episodes (one manic/mixed and one depressive or two manic episodes) and no residual mood incongruent psychotic symptoms during remissions. This information was also confirmed by first degree relatives for 64% of the cases. The illness age-of-onset was defined as the age at which the proband first met DSM-IV criteria for a manic, mixed, or major depressive episode. Family history of psychiatric illness was obtained with FIGS administered both to the patients and to all available relatives.

Cases in the ConLiGen-Romania study were ascertained in the same manner as for BOMA-Romania. Cases were required to have taken lithium for at least two years and lithium treatment response was evaluated with the Alda scale[105].

Population-based controls were evaluated using the DIGS and FIGS to screen for a lifetime history of major affective disorders, schizoaffective disorders, SCZ and other psychoses, obsessive-compulsive disorder, eating disorders, and alcohol or drug addiction. Unaffected individuals were included as controls in the present study.

#### ======== PGC4 Samples ========

**Grigoroiu-Serbanescu M | PMID : 31791676| Romania (BOMA-Romania) | rom4**

Cases were recruited from consecutive admissions to the Obregia Clinical Psychiatric Hospital, Bucharest, Romania. Patients were administered the DIGS[77] and FIGS[57] interviews. Information was also obtained from medical records and close relatives. The diagnosis of BP-I was assigned according to DSM-IV-TR criteria using the best estimate procedure. All patients had at least two hospitalized illness episodes. Population-based controls were evaluated using the DIGS to exclude a lifetime history of major affective disorders, schizophrenia, schizoaffective disorders, and other psychoses, obsessive-compulsive disorder, eating disorders, and alcohol or drug addiction.

**McQuillin A | PMID: 37643680 | UCL (University College London), London, UK | amq1**

Case and controls were collected using the protocol described above for bip_uclo_eur.

**Squassina A, | PMID: 21961650 | Italy | ital1**

Patients with bipolar I or bipolar II disorder were recruited at the outpatient unit (Lithium Clinic) of the Clinical Psychopharmacology Centre at the Department of Biomedical Science, Section of Neuroscience & Clinical Pharmacology, University of Cagliari, University Hospital Agency of Cagliari, Italy. Clinical assessments followed a strict procedure. After providing informed consent, participants were interviewed using one of the structured or semistructured interviews SADS-L. Clinical diagnosis was confirmed by DSM-IV criteria. We also used available medical records, narrative summaries of all interviews, and details such as baseline assessments, clinical course, response to treatment, treatment adherence, psychiatric and medical comorbidities, history of suicidal behavior, and symptom profiles in OPCRIT format.[55]

For uniform evaluation of treatment response, we used all available information including data from clinical records, diagnostic interviews, and prospective follow-up assessed by NIMH Life-Chart Method[106]. We used the Alda scale to assess lithium response[105].

**Manchia M, Carpiniello B, Squassina A | PMID: 35566641 | Italy | ital2**

The case samples were recruited among patients attending the outpatient clinic of the community mental health center of the Unit of Clinical Psychiatry within the University Hospital of Cagliari, Italy. Patients were enrolled in the genetic study if they met the following inclusion criteria: diagnosis of either Bipolar I or Bipolar II disorder according to DSM 5[107] criteria validated through the Italian version of the SCID-5-CV (Structured Clinical Interview for DSM-5 Clinical Version); being in euthymic phase.

All patients provided a written consent form regarding the use of their biological and clinical data for research purposes. Blood samples were gathered at the beginning of the study along with the relevant demographic and biometric data. All the clinical documents are stored in an anonymized database, accessible only by authorized personnel.

The recruited subjects were phenotypically characterized with the use of the following standardized tests:

- Brief Assessment of Cognition in Affective Disorders (BACA)
- Brief Assessment of Cognition in Schizophrenia to assess baseline cognitive capacities
- Hamilton Depression Rating Scale (HDRS)
- Young Mania Rating Scale (YMRS)
- Hamilton Anxiety Rating Scale (HAM-A)
- Barratt Impulsivity scale (BIS)
- Clinical Global Impression Scale – Severity (CGI-S)
- Alda score for Lithium response (clinical response defined as a score >7)
- OPCRIT

**Tondo L, Squassina A | PMID: 20348464 | Italy | ital3**

Our sample population encompasses a cohort of patients followed at the Mood Disorder Lucio Bini Center in Cagliari (Italy), a specialized outpatient clinic for the diagnosis, treatment and research of affective disorders. Since the founding of this outpatient clinic in 1977, all demographic and clinical information about patients have been recorded systematically by means of semi-structured initial and follow-up interviews, a life chart, extensive clinical evaluation and repeated assessments with standard rating scales for mood such as the Hamilton Depression Rating Scale (HDRS)[108], and Young Mania Rating Scale[109], typically every 4–6 weeks. Diagnoses were updated to meet the Diagnostic and Statistical Manual of Mental Disorders (DSM)-5 criteria[107] after the year 2013. Written informed consent was obtained for collection and analysis of patient data to be presented anonymously in aggregate form, in accordance with the requirements of Italian law and following review by a local ethical committee. Required data were entered into a computerized database in coded form to protect subject identity.

Patients were included in the study if they had at least 12 months of treatment with lithium and if they had a diagnosis of bipolar disorder or major depressive disorder according to DSM-5. The clinical response to lithium treatment was characterized using the "Retrospective Criteria of Long-Term Treatment Response in Research Subjects with Bipolar Disorder" scale, also known as Alda Scale[105].

**Kircher T, Dannlowski U | PMID 30267149| Germany | FOR 2107**

FOR 2107 is a longitudinal cohort study aiming to integrate clinical and neurobiological associations of genetic and environmental risk factors and their interaction involved in the **etiology**, onset and course of affective disorders[110]. Participants of the present study were part of the bi-center "Marburg Münster Affective Cohort Study" (MACS) and were recruited from in- and out-patient departments of the universities of Marburg and Münster, Germany, local psychiatric hospitals (Vitos Marburg, Gießen, Herborn, and Haina, LWL Münster, Germany), and using postings in local newspapers and flyers.

Lifetime BD was assessed using a semi-structured interview according to DSM-IV-TR (Diagnostic and Statistical Manual of Mental Disorders)[111] applied by trained staff and according to detailed SOPs. Controls did not report any lifetime history of psychiatric diagnosis or treatment as determined in line with DSM-IV-TR. All raters underwent standardized training, and interviews were regularly supervised and videotaped.

All procedures were approved by the local Ethics Committees of Marburg (AZ:07/14) and Münster (AZ:2014-422-b-S), Germany, according to the Declaration of Helsinki. Participants gave written informed consent prior to study participation and received financial compensation.

The study is funded by the German Research Foundation (Research Unit FOR 2107). Principal investigators are Tilo Kircher (KI588/14-1, KI588/14-2, KI588/20-1, KI588/22-1), Udo Dannlowski (DA 1151/5-1, DA 1151/5-2, DA1151/6-1), Axel Krug (KR3822/5-1, KR3822/7-2), Igor Nenadic (NE2254/1-2,NE2254/3-1,NE2254/4-1), Carsten Konrad (KO4291/3-1), Marcella Rietschel (RI 908/11-1, RI 908/11-2), Markus Nöthen (NO 246/10-1, NO 246/10-2), Stephanie Witt (WI 3439/3-1, WI 3439/3-2). We are deeply indebted to all study participants and staff. A list of acknowledgments can be found here: www.for2107.de/acknowledgements. Tilo Kircher received unrestricted educational grants from Servier, Janssen, Recordati, Aristo, Otsuka, neuraxpharm. Further information on the FOR 2107 data used for this article can be found here: www.for2107.de. Qualified researchers can request access to FOR 2107 data through contacting the principal investigators.

**Alda M | Not published | Nova Scotia, Canada | hal3**

The case samples were recruited from patients longitudinally followed at a specialty mood disorders clinic in Halifax (Canada). Cases were interviewed in a blind fashion with the Schedule of Affective Disorders and Schizophrenia-Lifetime version (SADS-L)[63] by pairs of clinician researchers (psychiatrists and/or nurses). The interviews together with medical records were subsequently reviewed in a blind fashion by a panel of senior clinical researchers. Consensus diagnoses were made according to DSM-IV[64] and Research Diagnostic Criteria[60] Protocols and procedures were approved by the local Ethics Committees and written informed consent was obtained from all patients before participation in the study.

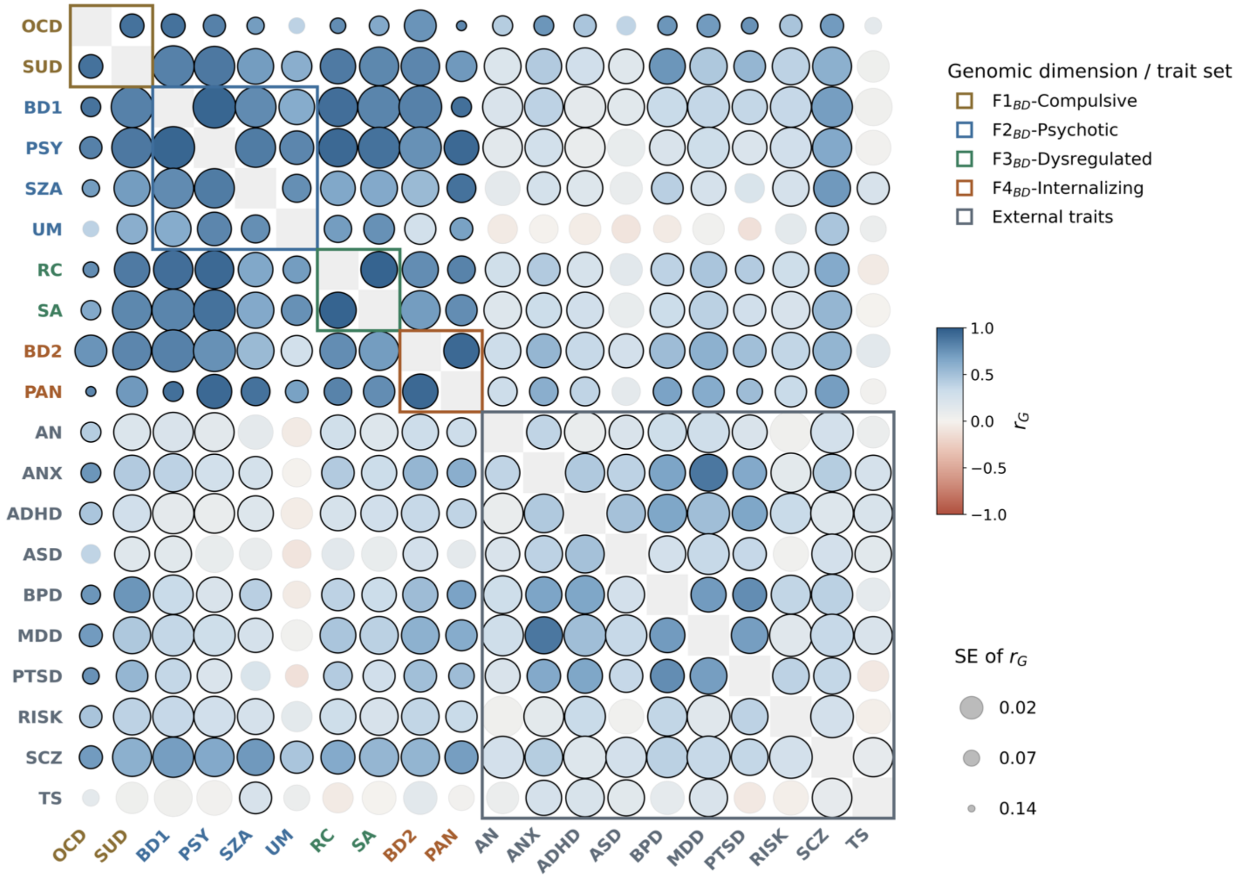

**Supplementary Fig.1**. **Genetic correlations between 10 bipolar disorder subphenotypes and 10 external traits**. Heatmap of pairwise bivariate LDSC genetic correlations ($r_G$) between the 10 bipolar disorder subphenotypes and 10 external psychiatric and behavioral traits **(Supplementary Table 9).** Color encodes $r_G$ (blue, positive; red, negative; from −1 to +1) and circle size encodes precision (larger circle = smaller standard error of $r_G$). The 10 BD subphenotypes are ordered and outlined by their assigned genomic dimension (F1$_{BD}$-Compulsive, F2$_{BD}$-Psychotic, F3$_{BD}$-Dysregulated, F4$_{BD}$-Internalizing), as in **Fig. 1b,** and **Fig.3**; the 10 external traits (AN, ANX, ADHD, ASD, BPD, MDD, PTSD, RISK, SCZ, TS) form a fifth block. The diagonal is fixed at 1. External summary statistics derive from the largest, most recently published GWAS. Cells outlined in black are Bonferroni-significant ($P < 2.63 \times 10^{-4}$); the remaining (faint gray outline) did not survive this stringent correction.

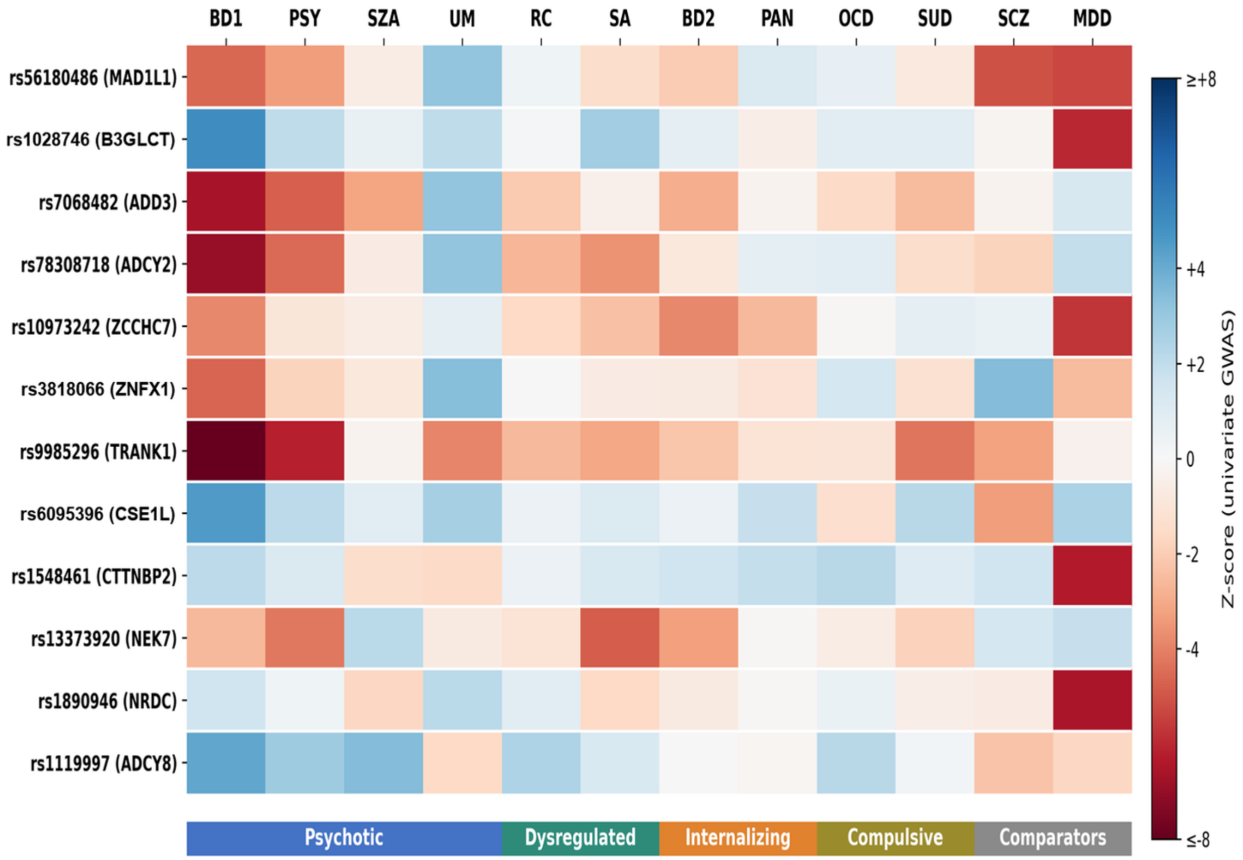

**Supplementary Fig.2. $Q_{SNP}$ heterogeneity loci: per-trait *z*-score heatmap**. Heatmap of $Q_{SNP}$ test *z*-scores across 10 bipolar subphenotypes and two external comparators (SCZ, MDD) for the **12** $Q_{SNP}$-heterogeneous loci identified by Genomic SEM common-factor analysis ($Q_{SNP}$ $P < 5 \times 10^{-8}$). Rows are loci labeled by lead SNP rsID and nearest gene; columns are traits grouped by latent factor domain (colored header bars: $F1_{BD}$-Compulsive, $F2_{BD}$-Psychotic, $F3_{BD}$-Dysregulated, $F4_{BD}$-Internalizing; see **Fig. 3**). Trait columns are grouped by factor color for convenience; underlying univariate *z*-scores are at the subphenotype level. Color scale reflects *z*-score magnitude and direction: blue = positive, red = negative, white = zero.

#### $F1_{BD}$-Compulsive

**OCD**

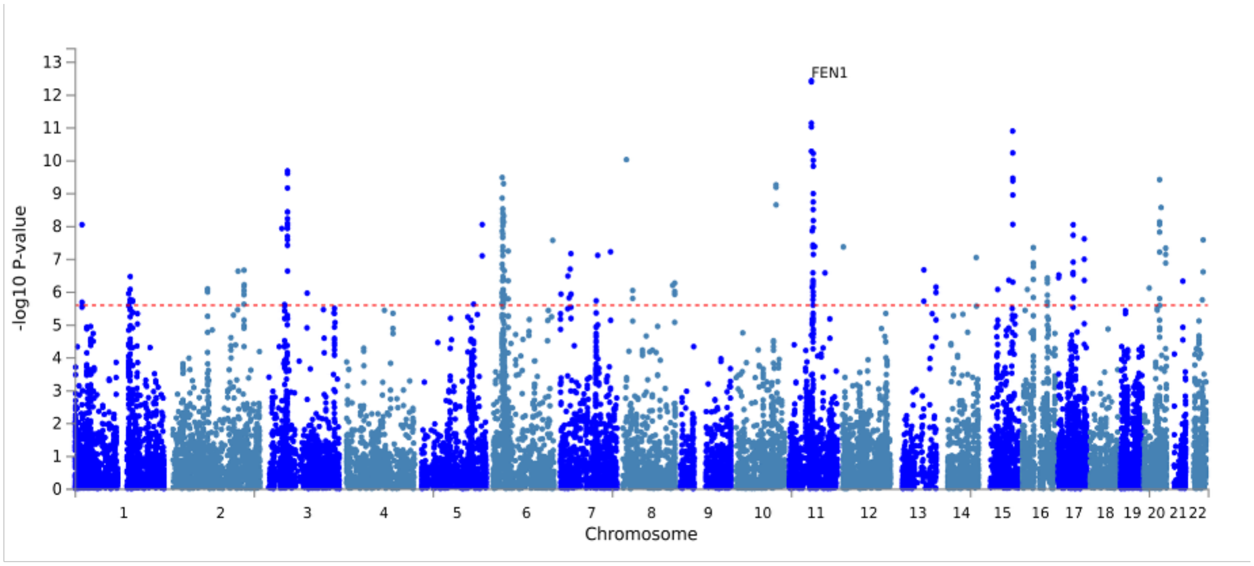

**SUD**

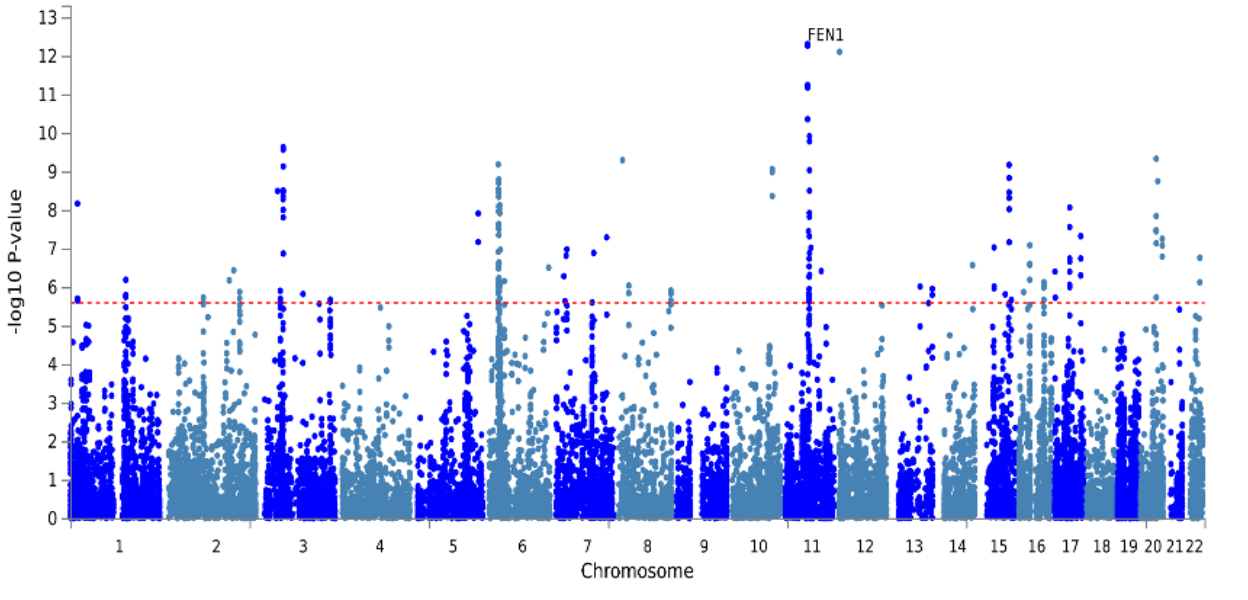

#### F2BD-Psychotic

**BD1**

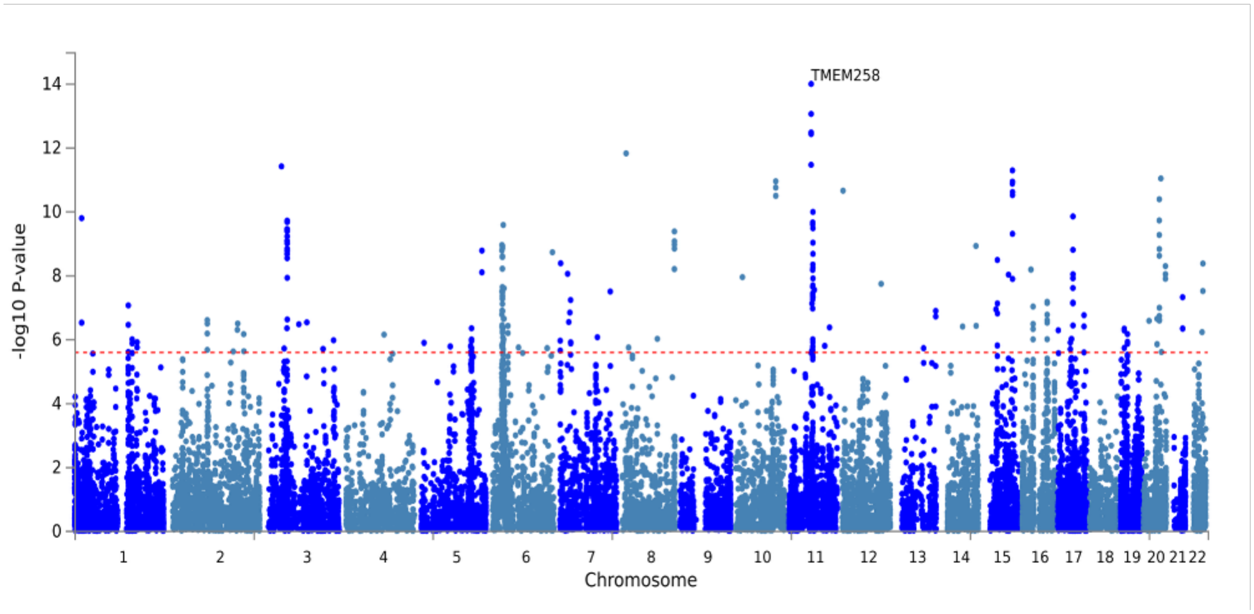

**PSY**

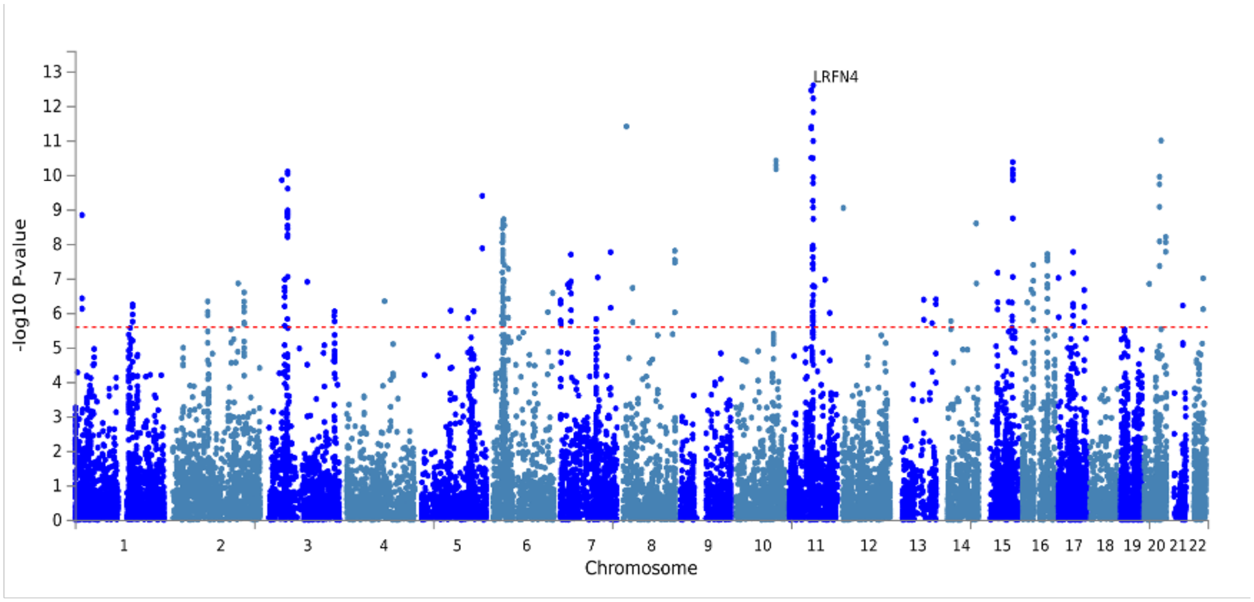

**SZA**

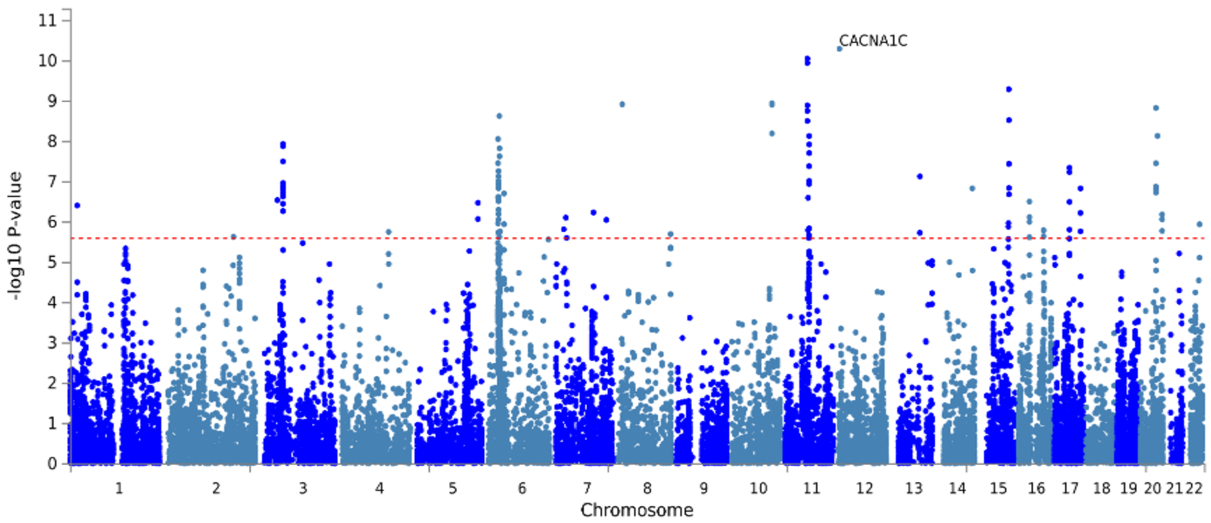

CACNA1C
-log10 P-value
0 1 2 3 4 5 6 7 8 9 10 11
1 2 3 4 5 6 7 8 9 10 11 12 13 14 15 16 17 18 19 20 21 22
Chromosome

**UM**

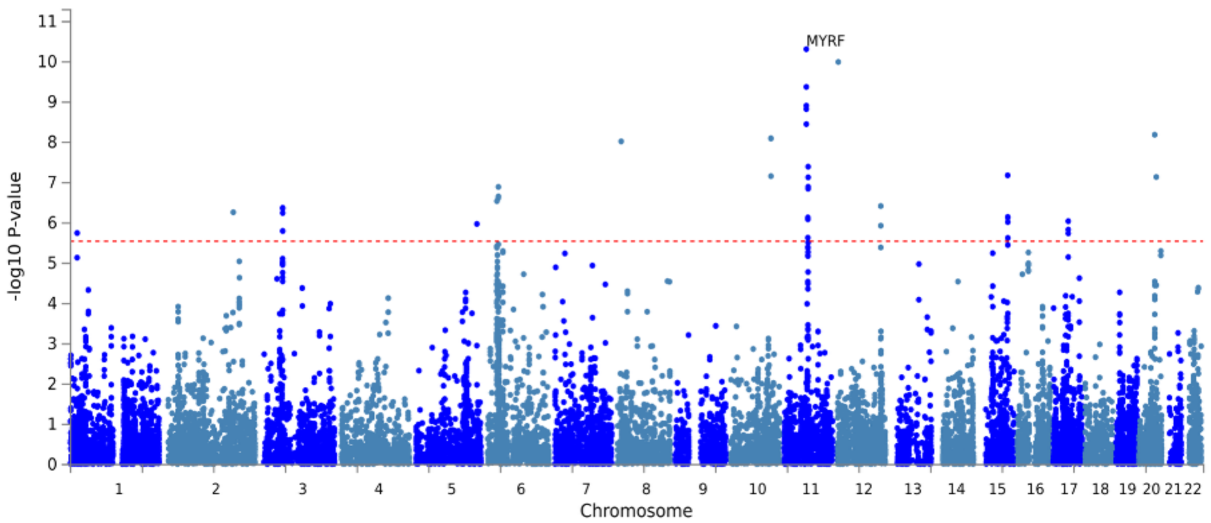

MYRF
-log10 P-value
0 1 2 3 4 5 6 7 8 9 10 11
1 2 3 4 5 6 7 8 9 10 11 12 13 14 15 16 17 18 19 20 21 22
Chromosome

#### F3$_{BD}$-Dysregulated

**RC**

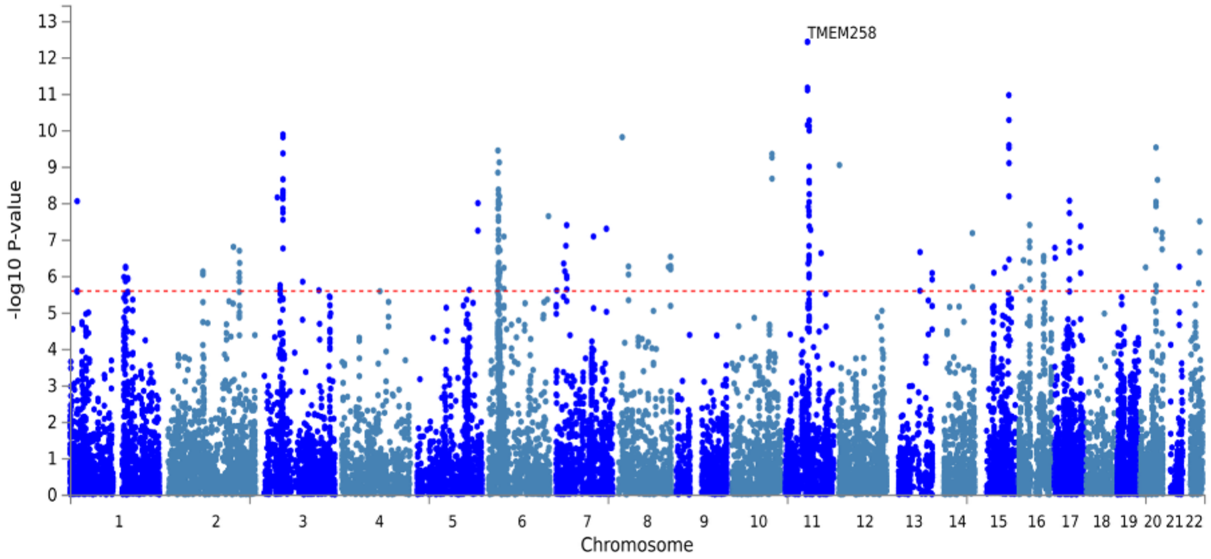

**SA**

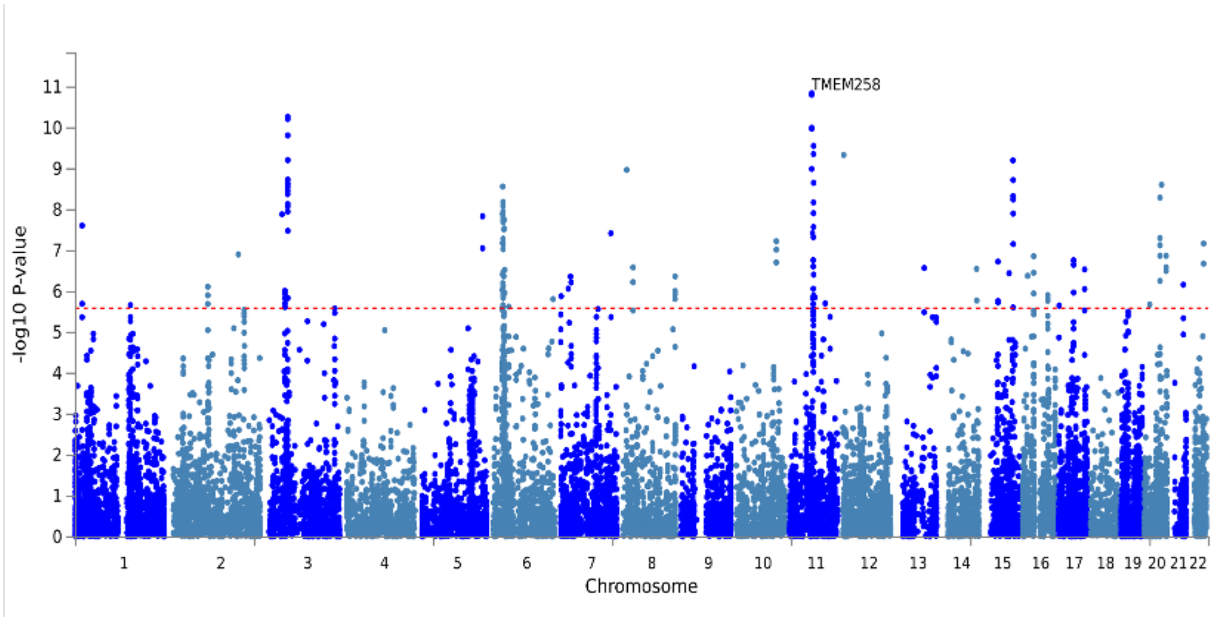

#### F4$_{BD}$-Internalizing

**PAN**

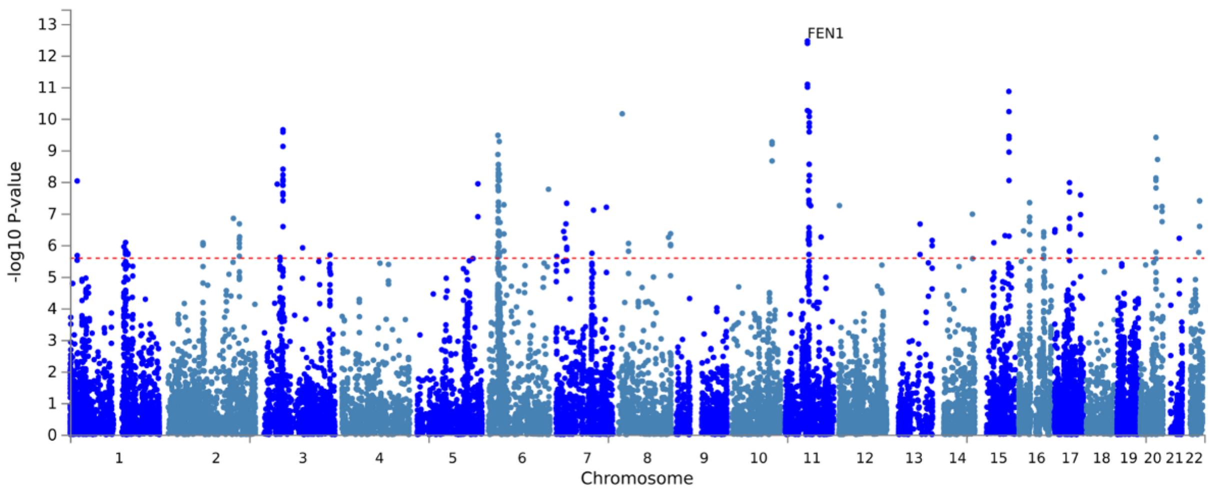

**BD2**

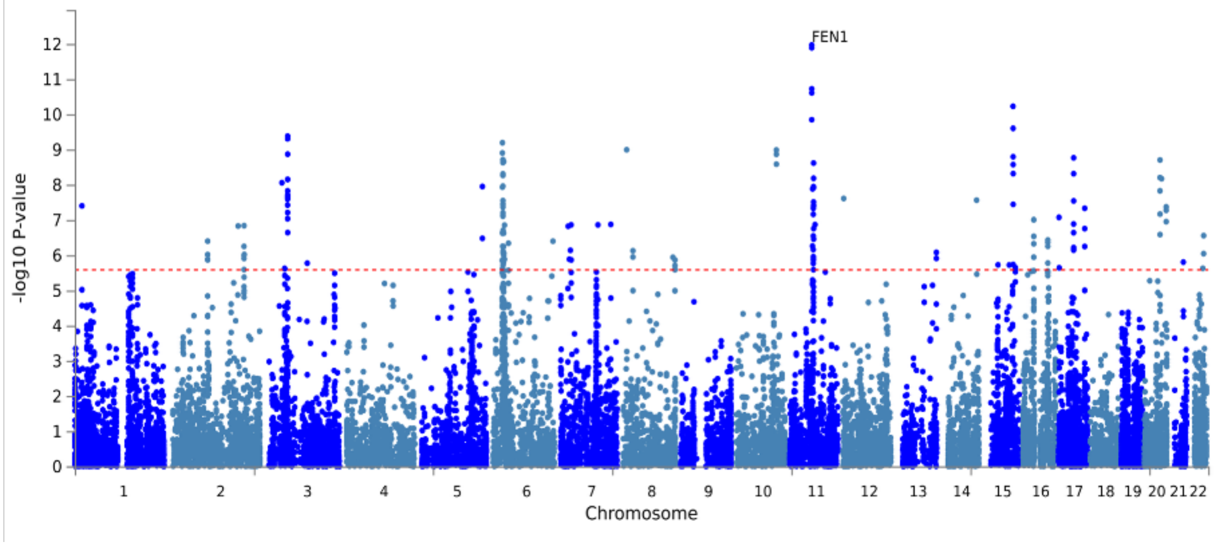

**Supplementary Fig.3**. **Gene-based Manhattan plots grouped by factor domain.** MAGMA gene-based association -log$_{10}$(*P*) values plotted by chromosomal position. Each panel corresponds to one of the 10 bipolar subphenotypes; panels are grouped by factor domain (F1$_{BD}$-Compulsive: OCD, SUD; F2$_{BD}$-Psychotic: BD1, PSY, SZA, UM; F3$_{BD}$-Dysregulated: RC, SA; F4$_{BD}$-Internalizing: PAN, BD2). Horizontal dashed line marks Bonferroni-corrected significance threshold ($P_{BONFERRONI} < 2.61\times10^{-6}$). The strongest gene-level hit per panel is labeled.

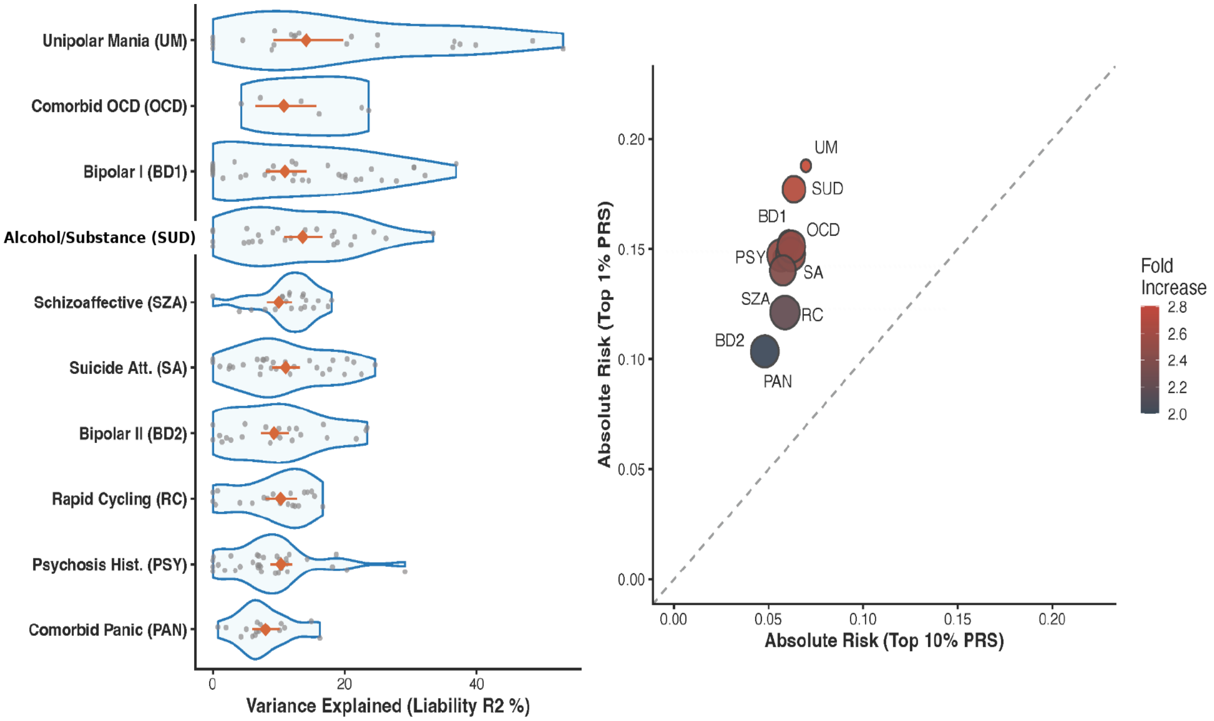

**Supplementary Fig.4. PRS performance heterogeneity and absolute risk stratification.** Variance-explained heterogeneity (liability $R^2$) across cohorts per subphenotype (violin plots, left side), with overlapping confidence intervals (CI 95%) indicating distributional overlap across the 10 BD subphenotypes. Effective sample sizes for the PRS target meta-analyses, computed as Neff = 4·N_case·N_control / (N_case + N_control) from the per-subphenotype case and control counts in **Supplementary Table 31**, range from 1,984 (OCD) to 38,153 (BD1) across the 10 subphenotypes. PRS absolute risk stratification (right side): top 1% vs. top 10% risk per subphenotype. Scatter plot of top 1% absolute risk (y-axis) vs. top 10% absolute risk (x-axis) for each of the 10 bipolar subphenotypes. All points are above the diagonal line indicating elevated risk in the extreme tail of the PRS distribution. Color denotes the fold increase in risk (top 1% relative to baseline). Circle area is proportional to the precision of the estimate (inverse standard error of the liability-scale $R^2$; **Supplementary Table 31**), so larger points denote more precisely estimated subphenotypes.

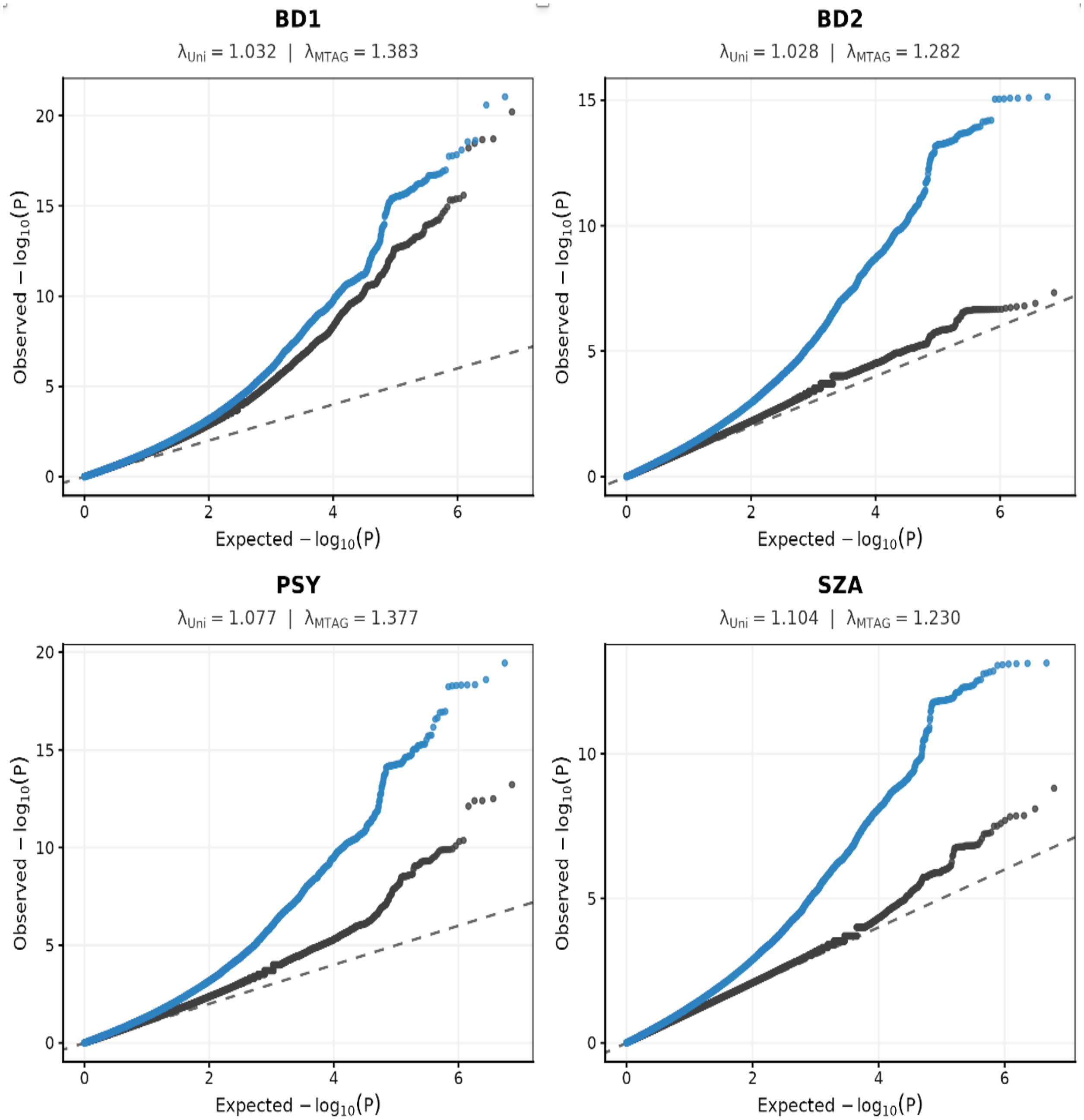
BD1
$\lambda_{Uni} = 1.032$ | $\lambda_{MTAG} = 1.383$
BD2
$\lambda_{Uni} = 1.028$ | $\lambda_{MTAG} = 1.282$
PSY
$\lambda_{Uni} = 1.077$ | $\lambda_{MTAG} = 1.377$
SZA
$\lambda_{Uni} = 1.104$ | $\lambda_{MTAG} = 1.230$
Observed $-\log_{10}(P)$
Expected $-\log_{10}(P)$

1726

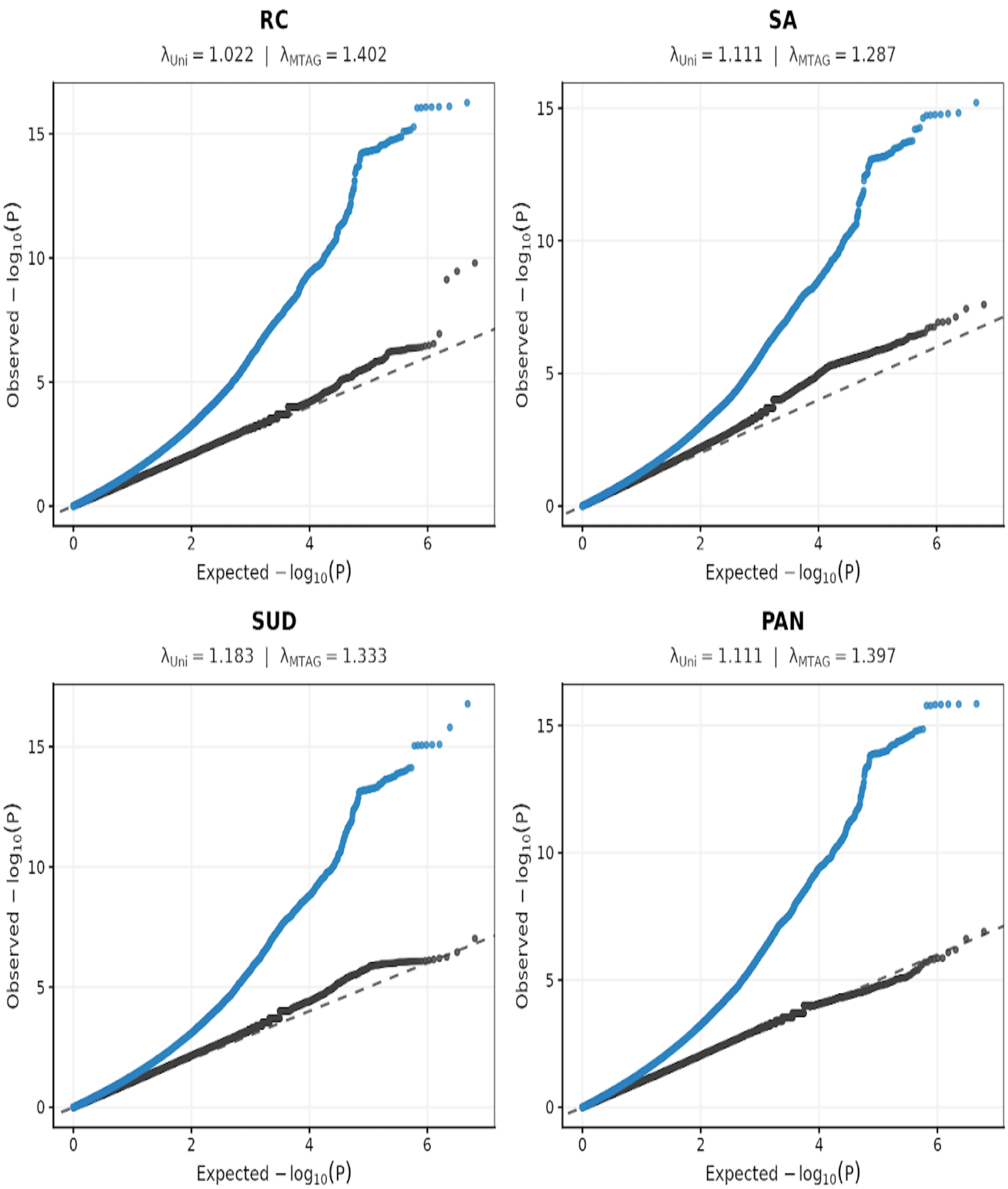
RC
λUni = 1.022 | λMTAG = 1.402
SA
λUni = 1.111 | λMTAG = 1.287
SUD
λUni = 1.183 | λMTAG = 1.333
PAN
λUni = 1.111 | λMTAG = 1.397
Observed − log10(P)
Expected − log10(P)
0
2
4
6
5
10
15

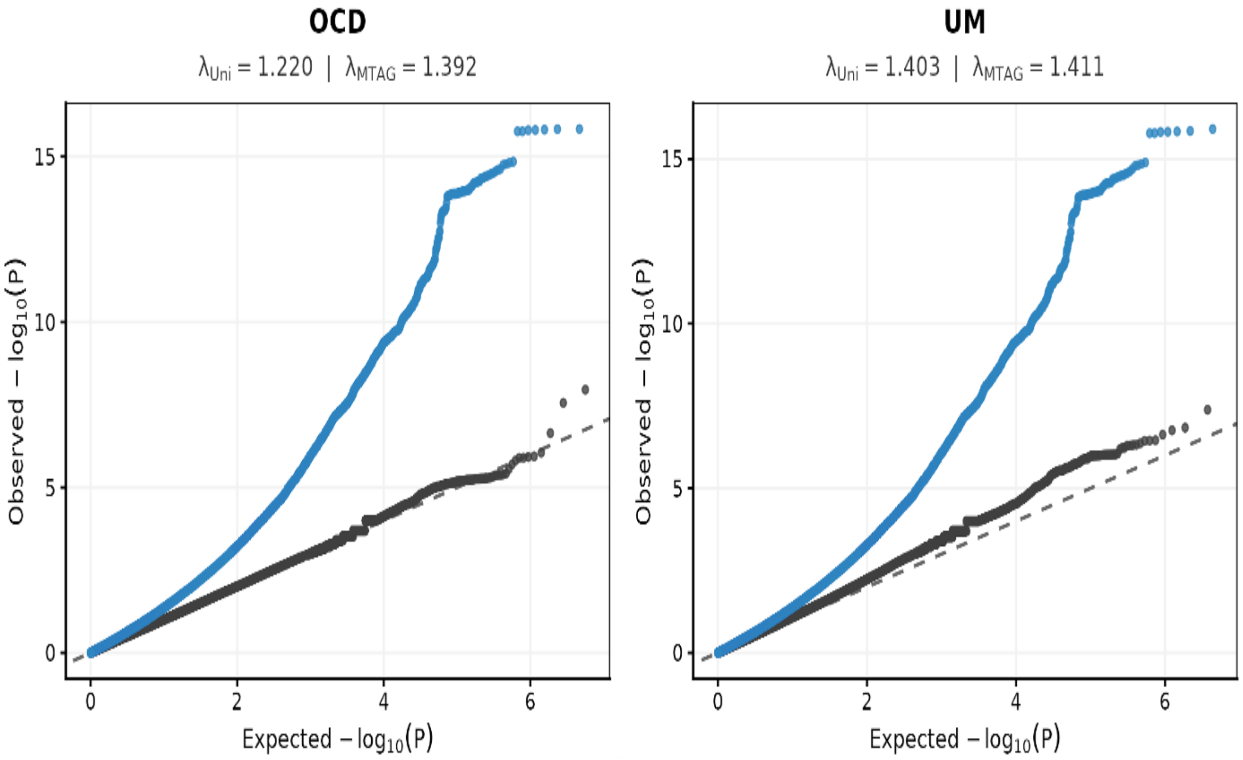

**Supplementary Fig.5. Quantile–quantile (Q–Q) plots comparing univariate GWAS and MTAG-enhanced association statistics.** The full meta-analytic sample (226,032 individuals; 38,022 cases, 188,010 controls) is the union of unique individuals across all subphenotypes; sample sizes vary across analyses. Per-subphenotype effective sample sizes (Neff = 4· N_case· N_control / (N_case + N_control)) for the contributing univariate GWAS are listed in **Supplementary Table 3** and range from 1,866 (OCD) to 95,808 (BD1) across the 10 subphenotypes. On QQ-plots, observed versus expected$-\log_{10}(P)$ for the baseline univariate GWAS (black) and the MTAG model (blue) for each of the 10 bipolar subphenotypes; each MTAG analysis augmented the subphenotype GWAS with additional BD SNP effects from the largest and most recent BD GWAS (O'Connell et al. 2025; European ancestry only, no self-report data). The earlier and steeper departure of the blue curves above the null diagonal reflects the gain in discovery power conferred by MTAG, and the corresponding rise in the genomic inflation factor (λ_GC) is an expected consequence of the increased effective sample size rather than confounding. Some curves fall marginally below the null diagonal across the bulk of the distribution, reflecting the mild conservativeness (deflation) of association testing at small effective sample sizes; such deflation is conservative with respect to discovery and does not affect the genome-wide-significant associations. Test statistics were otherwise well controlled: across the univariate GWAS, LDSC intercepts ranged from 1.002 to 1.047 (**Supplementary Table 2**), indicating that the bulk of the null distribution tracks the diagonal and that inflation is overwhelmingly polygenic in origin; the modest intercept elevation in the two smallest datasets (OCD and UM) likely reflects contraction of the mean-$\chi^2$ denominator in low-powered analyses rather than uncontrolled stratification[112]. Since the LDSC intercept is not an appropriate inflation diagnostic for MTAG output, MTAG validity was instead indexed by MaxFDR, which ranged from $1.7\times10^{-4}$ to $5.7\times10^{-4}$ across the 10 analyses[9], confirming that the additional associations reflect genuine power gain rather than spurious signal.

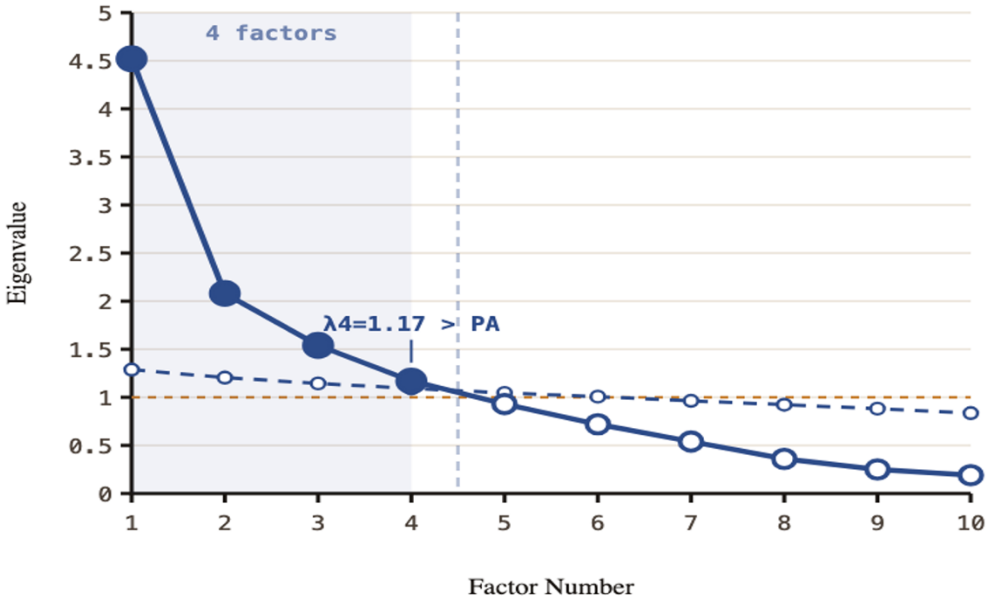

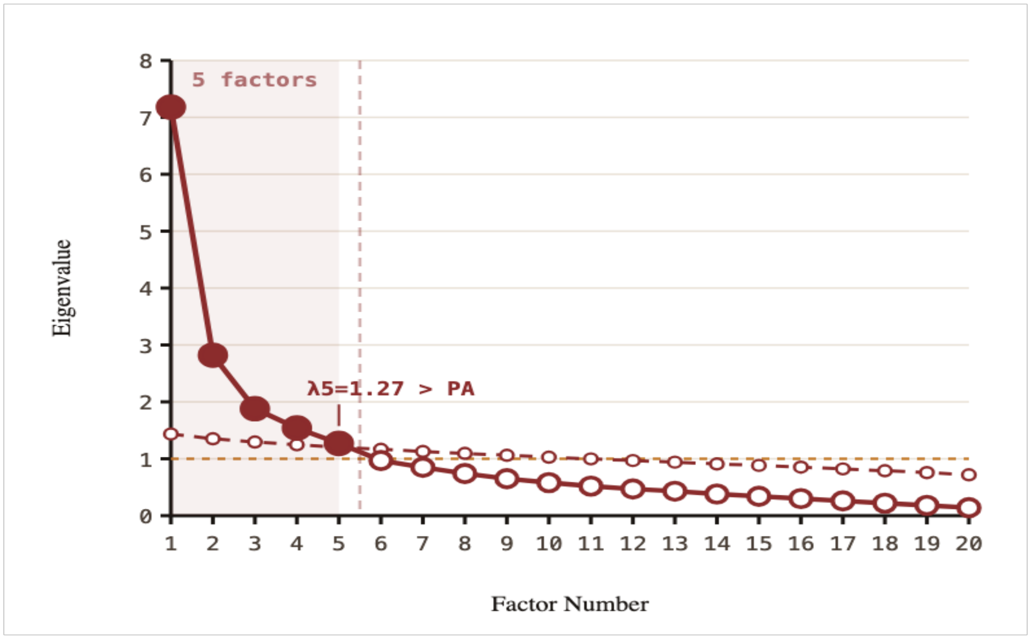

**Supplementary Fig.6**. **Scree plot and parallel analysis for EFA factor retention.** Eigenvalues from the exploratory factor analysis (EFA) of the 10-trait bipolar (blue) genetic covariance matrix, using 10,000 permutations (parallel analysis). The intersection points supported retention of four or five factors as indicated. Results for the 20-trait cross-disorder model are shown red.

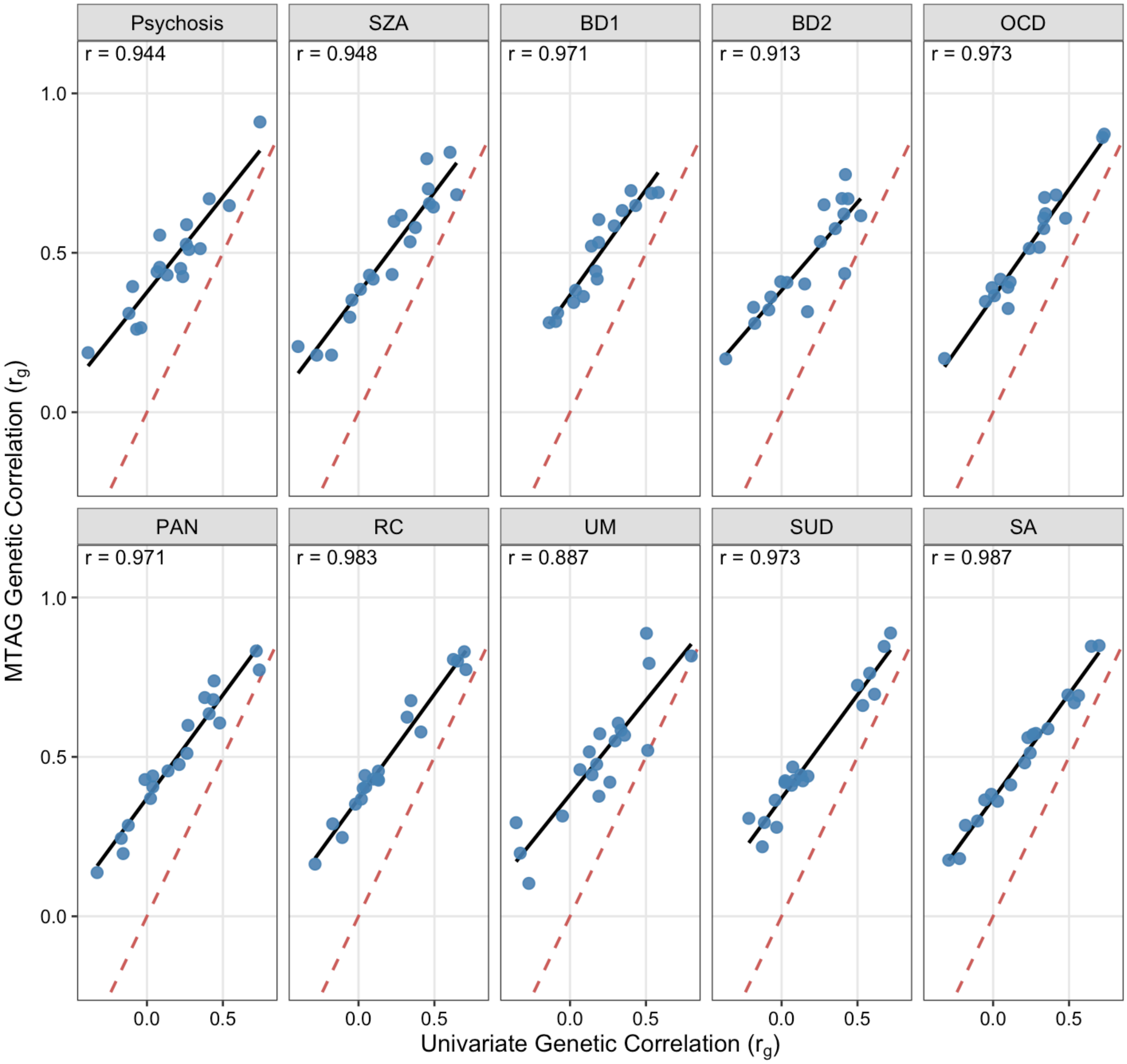

**Supplementary Fig.7. Univariate versus MTAG-enhanced rG comparisons across 10 bipolar disorder subphenotypes.** For each subphenotype, the scatter plot compares its pre-MTAG univariate GWAS-derived genetic correlations (x-axis) with its MTAG-enhanced genetic correlations (y-axis) across 19 external psychiatric, cognitive, personality, and sleep traits: (1) ADHD, (2) ANX, (3) ASD, (4) BPD, (5) Insomnia, (6) Intelligence, (7) MDD, (8) Matrix reasoning, (9) Memory, (10) AN, (11) PTSD, (12) SCZ, (13) Risk behavior [RISK], (14) Reaction Time, (15) Symbol Digit, (16) Trail-Making B (TMT-B), (17) Tower task, (18) Verbal Numerical Reasoning, and (19) TS. Each point is one external trait; the Pearson correlation (*r*) shown in each panel is computed across all 19 traits. The high concordance across the 10 subphenotypes indicates that MTAG enhancement preserves each subphenotype's trait-specific genetic signal. The red dashed line is the identity line (slope = 1); the black line is the observed regression fit.

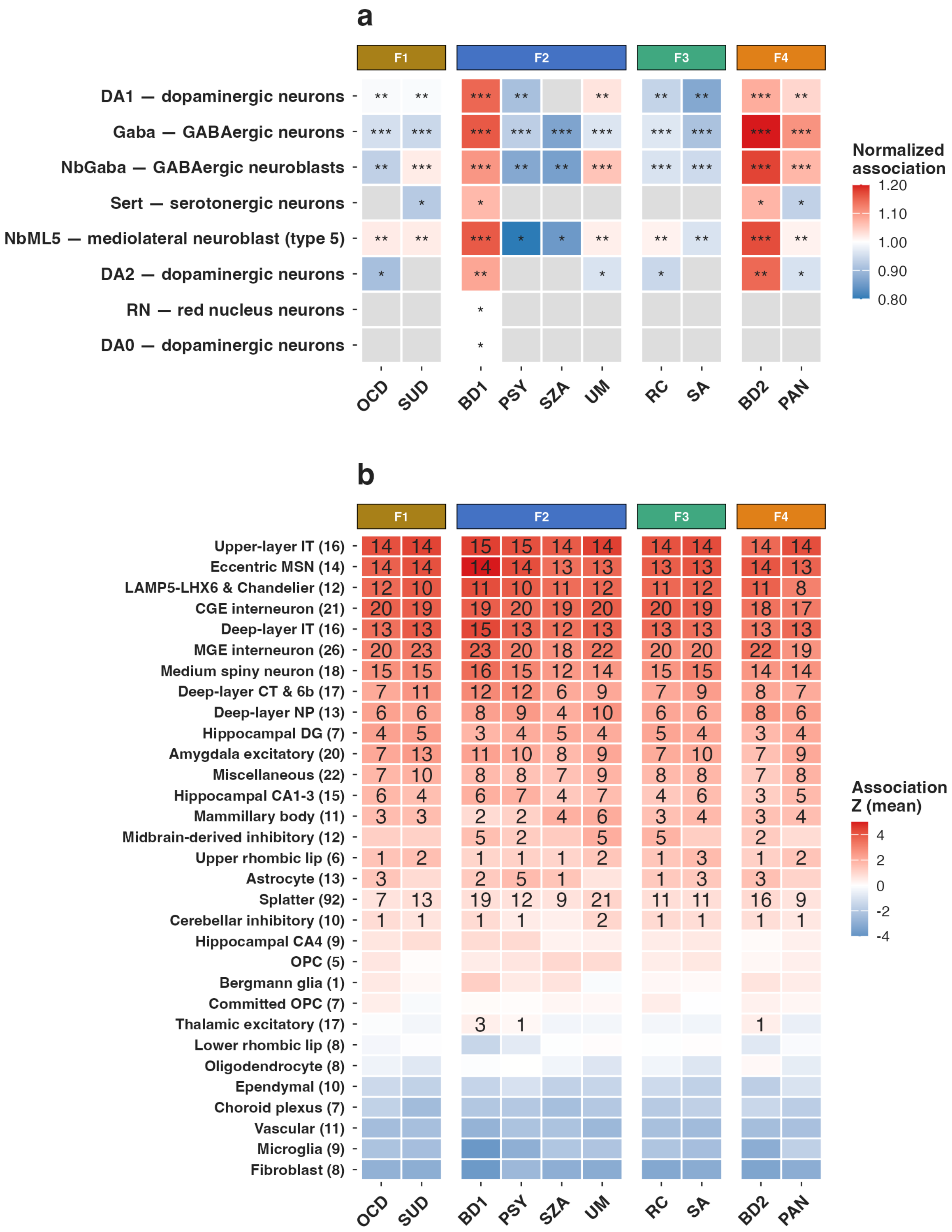

**Supplementary Fig. 8. Cell-type significance detail across 10 bipolar disorder subphenotypes. a.** Normalized association for 8 midbrain neuronal subclasses across the 10 BD subphenotypes, with Benjamini-Hochberg FDR significance marked (*FDR < 0.05, **FDR <0.01,

***FDR < 0.001). Color is red = above the cell's pan-BD baseline, blue = below. **b.** Mean association *z* for each of 31 superclusters (rows), ordered top to bottom by mean *z*. In-cell numbers give the count of constituent clusters reaching FDR < 0.05 in each subphenotype (blank = none). Subphenotype columns are grouped by factor for convenience as in **Fig. 3** (F1$_{BD}$-Compulsive, F2$_{BD}$-Psychotic, F3$_{BD}$-Dysregulated, F4$_{BD}$-Internalizing) (**Supplementary Table 28f**).
